# An *APOE*\*4-Informed Genomic Atlas of the X Chromosome in Alzheimer’s Disease

**DOI:** 10.64898/2026.05.05.26352461

**Authors:** Noah Cook, Youjie Zeng, Chenyu Yang, Zhiwen Jiang, Ting-Chen Wang, Yann Le Guen, Karly Cody, Matthew Johnson, Rui Zhang, Victoria C. Merritt, Richard L. Hauger, the VA Million Veteran Program, FinnGen, Mary Ellen Koran, Elizabeth C. Mormino, Brian Gordon, Alex DeCasien, Shea Andrews, Logan Dumitrescu, Derek Archer, Timothy J. Hohman, Cyril Pottier, Carlos Cruchaga, Richard Sherva, Mark Logue, Valerio Napolioni, Michael D. Greicius, Michael E. Belloy

## Abstract

The genetic contributions of the X chromosome to Alzheimer’s disease (AD) remain poorly understood yet are expected to importantly shape sex differences in AD. We therefore performed large-scale X-chromosome-wide association studies (N=1,240,451), evaluating differential risk due to sex, *APOE*\*4, and escape from X-chromosome inactivation, finding most X-linked loci appear relevant to female-biased AD etiology. In evaluating genetic pleiotropy with hormonal, lipid, and brain imaging traits, we discovered X-linked AD loci converged on white matter traits, particularly in the anterior corona radiata and splenium of the corpus callosum. Through brain-centric functional genomics analyses, we then nominated candidate causal genes, including 5 that appeared highly robust. Notably, we found the escape gene *RBBP7* decreases AD risk in *APOE*\*4 carriers likely through higher expression in excitatory neurons to counter tau-related neurodegeneration. Altogether, we provide an atlas of sex and *APOE*\*4-informed candidate X-linked AD risk loci, genes, and mechanisms that will guide future studies.

## Introduction

Genome-wide association studies (GWAS) have transformed our understanding of Alzheimer’s disease (AD), revealing over 80 genetic loci and enabling the prioritization of candidate genes leading to new biological insights and therapeutic targets^1,2^. However, studies including the X chromosome have lagged behind autosome-focused GWAS^3,4^, despite the X chromosome comprising 5% of the genome, being enriched for brain and immune-relevant genes^5,6^, and contributing to sex differences in brain aging and AD^7,8^. Together with converging evidence that sex differences are integral to AD, including higher prevalence in women^9–11^ and differences in genetic risk^12,13^, clinicopathological profiles^14–17^, and treatment response^18^, deep knowledge of X-linked genetic variation in AD is crucially needed.

The relative scarcity of X chromosome-wide genetic association studies (XWAS) reflects multiple technical and analytical hurdles^3,4^. Males carry a single copy of the X chromosome (hemizygous) whereas females carry two copies but undergo random X-chromosome inactivation (rXCI), a process in which one X chromosome is randomly silenced to balance gene expression with males^19^. However, rXCI is incomplete and heterogeneous with approximately 30% of genes escaping X-chromosome inactivation (eXCI), which can result in double gene dosages in women that in turn can shape sex biases^20,21^. Ultimately, compared to the autosomes, different genetic modelling strategies are required. Additionally, male hemizygosity reduces power by 50%, while in women, uncertainty of the active genotype due to rXCI reduces power by 75%^19^, such that XWAS need substantially larger sample sizes than autosomal GWAS. Despite these barriers, methodological advances and expanding genetic datasets have led to successful XWAS in several complex traits, including cardiovascular disease^22,23^, kidney disease^24^, Parkinson’s disease^25^, and brain imaging measures^6,26^. Two recent large-scale XWAS by Belloy *et al.* and Le Borgne *et al.* identified several X-linked loci for AD^27,28^, yet few reached X-wide significance criteria and only one locus in Belloy *et al.*, *SLC9A7*, passed a stricter genome-wide significance threshold despite a sample size of N=1,152,284. These findings highlight persistent power challenges for X chromosome analyses and suggest that important sub-threshold signals remain to be elucidated. Crucially, sex-stratified and eXCI insights remained incomplete and the role of the *APOE*\*4 allele–the strongest genetic risk factor for late-onset AD–was not assessed, despite extensive evidence that *APOE*\*4 confers higher AD risk and tau pathology in women relative to men^29–32^. Recent *APOE*\*4-stratified autosomal studies have also successfully identified modifiers of *APOE*-related AD risk^33–36^, underscoring the need for analogous analyses on the X chromosome.

To address these gaps, we increased sample sizes for AD XWAS and combined rXCI and eXCI modeling with sex- and *APOE*\*4-stratified analyses (**Fig.1**). To mitigate power challenges, we identified subthreshold AD risk loci that displayed shared pleiotropic signals with recent XWASs of AD-relevant traits, with a focus on brain neuroimaging traits. Using this comprehensive atlas of potential risk loci, we mapped brain areas where X-linked genetic risk converges and may thus contribute to AD etiology. Finally, we performed *in silico* functional genomics follow-up studies which included leveraging large brain transcriptomics datasets to prioritize the most promising loci and their candidate causal genes.

**Figure 1.**
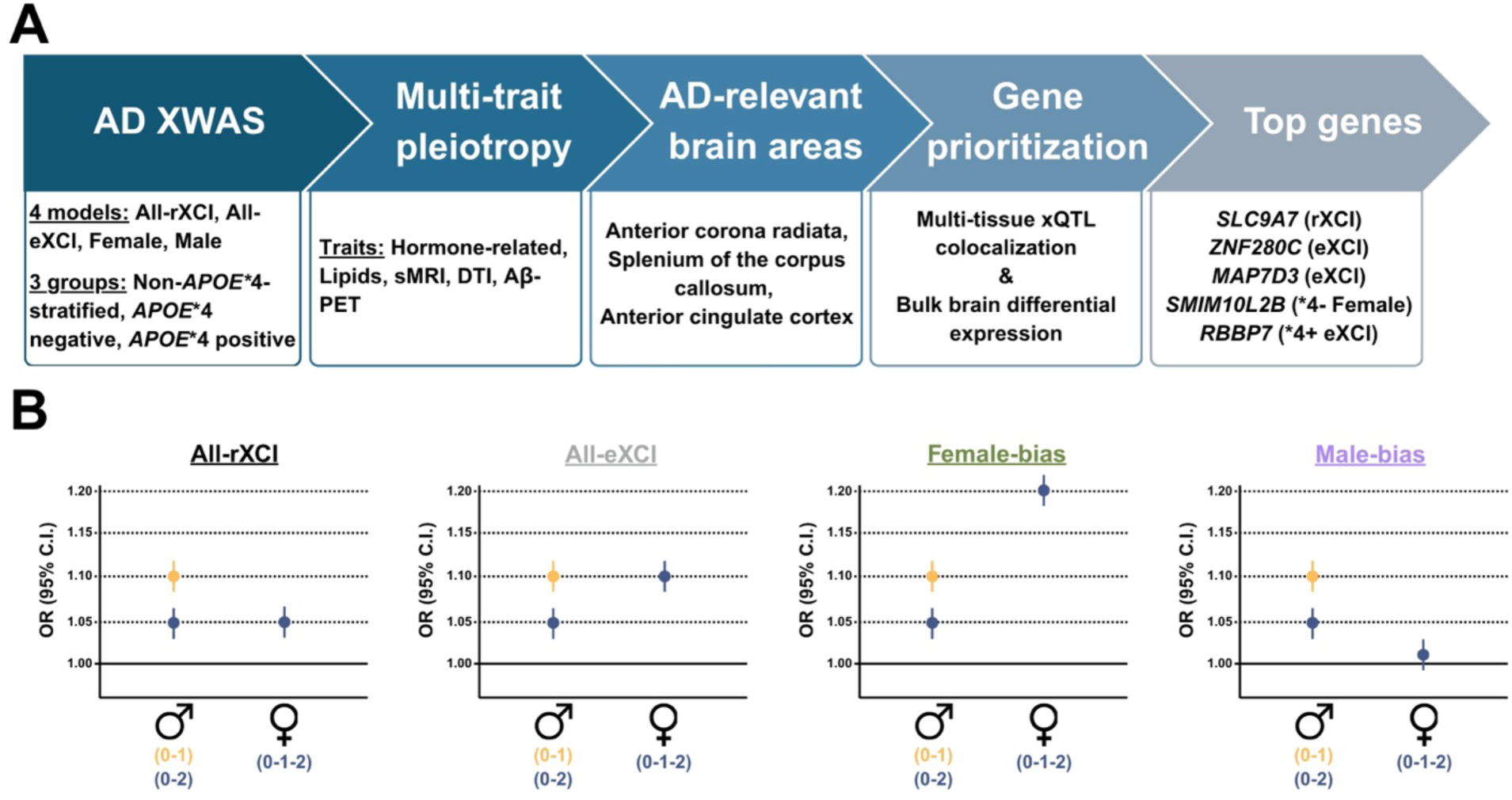
Study overview and framework for X chromosome genetic association analyses in Alzheimer’s disease. **A)** Schematic of the study design. Alzheimer’s disease (AD) X-chromosome-wide association analyses (XWAS) were performed under four models or sex strata (All-rXCI, All-eXCI, female-specific, and male-specific) and across three *APOE*\*4 strata. Associated loci were evaluated for multi-trait pleiotropy, enrichment in AD-relevant brain regions, and functional genomics support using multi-tissue xQTL colocalization and bulk brain differential expression for gene prioritization. **B)** Conceptual illustration of expected XWAS effect-size patterns under alternative X-chromosome inactivation (XCI) assumptions and sex-biased analyses. Under the rXCI model (full dosage compensation), females are assumed to have one X chromosome copy randomly inactivated, such that genotype dosages 0-1-2 relate to a 0-0.5-1 probability of association, effectively reducing variant effect sizes by 50%. Male hemizygous genotype dosages are thus encoded as 0-2 to reduce the effect estimate by 50% and be equivalent with women. Under the eXCI model, females are assumed to not undergo XCI (full escape), such that variant effect estimates are not diluted and align with male effect sizes for genotype dosages encoded as 0-1. Female- and male-biased XWAS aimed to capture sex-effect heterogeneity beyond eXCI and rXCI differences.

## Results

### X-chromosome-wide association studies of Alzheimer’s disease

We performed 12 XWAS of AD across European ancestry subjects (**Fig.1**), reflecting combinations of 4 models or sex strata (All-rXCI, All-eXCI, Females, Males) with 3 *APOE*\*4 strata (non-*APOE*\*4-stratified, *APOE*\*4-, *APOE*\*4+). XWAS meta-analyses were conducted across clinically diagnosed subjects from the Alzheimer’s Disease Genetics Consortium (ADGC, **Table-S1**) and Alzheimer’s Disease Sequencing Project (ADSP; **Table-S2**)^37,38^, and health-registry and family-proxy-defined cases and controls from the UK Biobank (UKB), FinnGen, and Million Veterans Program (MVP), with total samples sizes including up to 143,465 AD cases (82,386 proxy-cases) and 1,096,986 controls (**Fig.2A**, for detailed demographic cf. **Table-S3,** for phenotype definitions cf. Methods). These samples largely overlapped those in Belloy *et al.*, but eXCI and *APOE*\*4-stratified analyses were novel, and sex-stratified analyses newly included samples from FinnGen^27^. Across all XWAS, quantile-quantile plots showed no evidence of inflation (**Fig.S1**).

**Figure 2.**
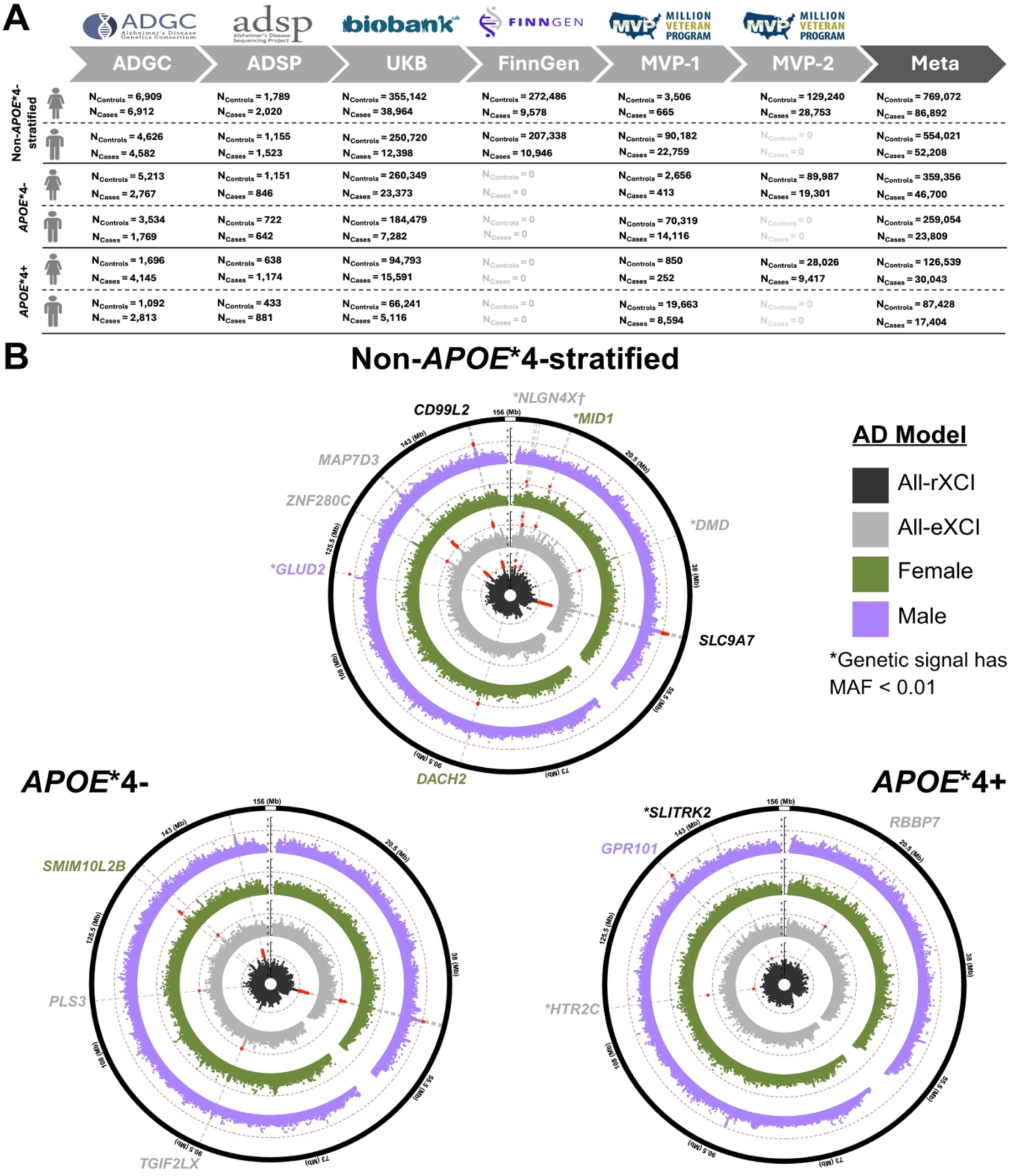
Sex- and *APOE*\*4-stratified X-chromosome-wide association results for Alzheimer’s disease. **A)** Summary of case-control sample sizes across cohorts for sex and *APOE*\*4 stratified XWAS (detailed demographics are in Table-S3). **B)** Circos plots summarize XWAS results across respective *APOE*\*4 strata. Concentric tracks represent the rXCI model (inner, black), eXCI model (second, grey), female-specific analysis (third, green), and male-specific analysis (outer, purple). Red points denote independent signals reaching X-wide significance P<1e-5. Gene labels indicate nearest protein-coding gene or a functionally prioritized gene; gene name color reflects the model with the most significant association for the signal. *† NLGN4X locus contains 3 independent X-wide significant signals in the eXCI (2) and female (1) analyses. The most significant signal arose from the eXCI model, and the locus was classified accordingly*.

Given the known power limitations of XWAS, we wielded an explorative approach using the conventional, suggestive significance cut-off (P<1e-5) for detection of X-wide significant signals without further multiple-comparison correction, leveraging subsequent genomics analyses to prioritize the most promising loci (cf. later results). All significant signals were evaluated for independence, while signals observed in sex or *APOE*\*4 strata were evaluated respectively for cross-sex or *APOE*\*4 effect heterogeneity, retaining only those with at least FDR-P_Het_<0.05 (cf. Methods). Using this approach, we identified 18 independent association signals across 16 loci (**Table-1, Table-S4, Fig.2B**; locus zoom plots in **Fig.S2**), of which non-*APOE*\*4-stratified signals near *NLGN4X* (eXCI), *MID1* (female), *SLC9A7* (rXCI), *ZNF280C* (eXCI), *MAP7D3* (eXCI), and *CD99L2* (rXCI, previously labelled *MTM1*) replicated those in Belloy *et al.*^27^, while those near *NLGN4X* (eXCI)*, DMD* (eXCI), and *DACH2* (female) overlapped loci reported in Le Borgne *et al.* but represented independent signals (R2<0.01 across index variants)^28^, and one signal near *GLUD2* (male) represented a novel locus. The *SLC9A7* locus remained the only locus to reach genome-wide significance (P<5e-8)^27^. Stratification by *APOE*\*4 carrier status further revealed 3 novel *APOE*\*4- signals near *TGIF2LX* (eXCI), *PLS3* (eXCI), and *SMIM10L2B* (female), and 4 novel *APOE*\*4+ signals near *RBBP7* (eXCI), *HTR2C* (eXCI), *GPR101* (male), and *SLITRK2* (rXCI). Out of all 16 risk loci, the majority appeared potentially relevant to female AD etiology, including 8 eXCI loci, which may relate to candidate genes with double dosage in women relative to men, and 3 female-biased loci with distinctly larger effect sizes relative to men. To assess signal robustness, forest plots were generated and cross-cohort effect heterogeneity evaluated for each of the 18 independent lead variants, indicating effect sizes were generally consistent across cohorts (**Fig.S2**). We further compared effect estimates across full meta-analyses, meta-analyses excluding proxy phenotypes, and proxy-only analyses, and found they were largely consistent. Across these checks, the only notable deviation was observed at the *DMD* locus, which showed an attenuated effect size in proxy-only analyses (**Fig.S3**). Importantly, none of the discovered signals appeared to be specifically driven by the inclusion of proxy-phenotype XWAS.

**Table 1.**
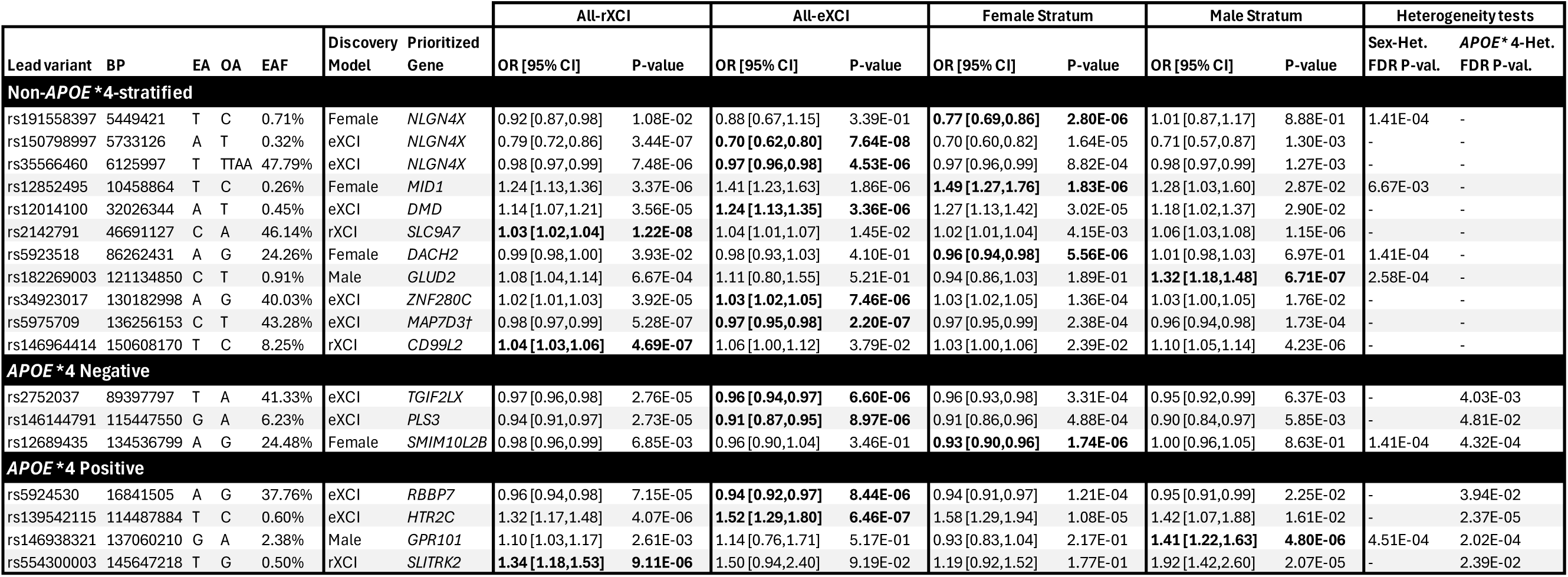
Sex- and *APOE*\*4-stratified X-chromosome-wide association study of Alzheimer’s disease: Associated lead variants. Listed lead variants passed XWAS significance thresholds (P<1e-5) for a respective model or sex and *APOE*\*4 stratum; Sex and *APOE*\*4 stratified findings also passed FDR-P<0.05 on respective cross-stratum heterogeneity tests. Bolded cells represent the most significant model or stratum. The ‘Prioritized Gene’ column represents either the nearest protein-coding gene, or gene that was functionally prioritized in downstream analysis. Variants are annotated using dbSNP153. An expanded version of this table including sample sizes for respective associations is provided in Table-S4. † For the *MAP7D3* locus, the reported variant corresponds to the third most significant XWAS signal at the locus, as it demonstrated colocalization with external traits and xQTL signals. The two more significant variants at this locus (LD R^2^ ≈ 0.4; cf. Fig.S2.10) did not show evidence of colocalization with external traits or molecular phenotypes. ***Abbreviations:*** *BP, base pair position (hg38); EA, effect allele; OA, other allele; EAF, effect allele frequency; OR, odds ratio; CI, confidence interval*.

|  |  |  |  |  |  |  | All-rXCI |  | All-eXCI |  | Female Stratum |  | Male Stratum |  | Heterogeneity tests |  |
| --- | --- | --- | --- | --- | --- | --- | --- | --- | --- | --- | --- | --- | --- | --- | --- | --- |
| Lead variant | BP | EA | OA | EAF | Discovery Model | Prioritized Gene | OR [95% CI] | P-value | OR [95% CI] | P-value | OR [95% CI] | P-value | OR [95% CI] | P-value | Sex-Het. FDR P-val. | *APOE* *4-Het. FDR P-val. |
| **Non-*APOE*\*4-stratified** |  |  |  |  |  |  |  |  |  |  |  |  |  |  |  |  |
| rs191558397 | 5449421 | T | C | 0.71% | Female | *NLGN4X* | 0.92 [0.87,0.98] | 1.08E-02 | 0.88 [0.67,1.15] | 3.39E-01 | **0.77 [0.69,0.86]** | **2.80E-06** | 1.01 [0.87,1.17] | 8.88E-01 | 1.41E-04 | - |
| rs150798997 | 5733126 | A | T | 0.32% | eXCI | *NLGN4X* | 0.79 [0.72,0.86] | 3.44E-07 | **0.70 [0.62,0.80]** | **7.64E-08** | 0.70 [0.60,0.82] | 1.64E-05 | 0.71 [0.57,0.87] | 1.30E-03 | - | - |
| rs35566460 | 6125997 | T | TTAA | 47.79% | eXCI | *NLGN4X* | 0.98 [0.97,0.99] | 7.48E-06 | **0.97 [0.96,0.98]** | **4.53E-06** | 0.97 [0.96,0.99] | 8.82E-04 | 0.98 [0.97,0.99] | 1.27E-03 | - | - |
| rs12852495 | 10458864 | T | C | 0.26% | Female | *MID1* | 1.24 [1.13,1.36] | 3.37E-06 | 1.41 [1.23,1.63] | 1.86E-06 | **1.49 [1.27,1.76]** | **1.83E-06** | 1.28 [1.03,1.60] | 2.87E-02 | 6.67E-03 | - |
| rs12014100 | 32026344 | A | T | 0.45% | eXCI | *DMD* | 1.14 [1.07,1.21] | 3.56E-05 | **1.24 [1.13,1.35]** | **3.36E-06** | 1.27 [1.13,1.42] | 3.02E-05 | 1.18 [1.02,1.37] | 2.90E-02 | - | - |
| rs2142791 | 46691127 | C | A | 46.14% | rXCI | *SLC9A7* | **1.03 [1.02,1.04]** | **1.22E-08** | 1.04 [1.01,1.07] | 1.45E-02 | 1.02 [1.01,1.04] | 4.15E-03 | 1.06 [1.03,1.08] | 1.15E-06 | - | - |
| rs5923518 | 86262431 | A | G | 24.26% | Female | *DACH2* | 0.99 [0.98,1.00] | 3.93E-02 | 0.98 [0.93,1.03] | 4.10E-01 | **0.96 [0.94,0.98]** | **5.56E-06** | 1.01 [0.98,1.03] | 6.97E-01 | 1.41E-04 | - |
| rs182269003 | 121134850 | C | T | 0.91% | Male | *GLUD2* | 1.08 [1.04,1.14] | 6.67E-04 | 1.11 [0.80,1.55] | 5.21E-01 | 0.94 [0.86,1.03] | 1.89E-01 | **1.32 [1.18,1.48]** | **6.71E-07** | 2.58E-04 | - |
| rs34923017 | 130182998 | A | G | 40.03% | eXCI | *ZNF280C* | 1.02 [1.01,1.03] | 3.92E-05 | **1.03 [1.02,1.05]** | **7.46E-06** | 1.03 [1.02,1.05] | 1.36E-04 | 1.03 [1.00,1.05] | 1.76E-02 | - | - |
| rs5975709 | 136256153 | C | T | 43.28% | eXCI | *MAP7D3†* | 0.98 [0.97,0.99] | 5.28E-07 | **0.97 [0.95,0.98]** | **2.20E-07** | 0.97 [0.95,0.99] | 2.38E-04 | 0.96 [0.94,0.98] | 1.73E-04 | - | - |
| rs146964414 | 150608170 | T | C | 8.25% | rXCI | *CD99L2* | **1.04 [1.03,1.06]** | **4.69E-07** | 1.06 [1.00,1.12] | 3.79E-02 | 1.03 [1.00,1.06] | 2.39E-02 | 1.10 [1.05,1.14] | 4.23E-06 | - | - |
| ***APOE*\*4 Negative** |  |  |  |  |  |  |  |  |  |  |  |  |  |  |  |  |
| rs2752037 | 89397797 | T | A | 41.33% | eXCI | *TGIF2LX* | 0.97 [0.96,0.98] | 2.76E-05 | **0.96 [0.94,0.97]** | **6.60E-06** | 0.96 [0.93,0.98] | 3.31E-04 | 0.95 [0.92,0.99] | 6.37E-03 | - | 4.03E-03 |
| rs146144791 | 115447550 | G | A | 6.23% | eXCI | *PLS3* | 0.94 [0.91,0.97] | 2.73E-05 | **0.91 [0.87,0.95]** | **8.97E-06** | 0.91 [0.86,0.96] | 4.88E-04 | 0.90 [0.84,0.97] | 5.85E-03 | - | 4.81E-02 |
| rs12689435 | 134536799 | A | G | 24.48% | Female | *SMIM10L2B* | 0.98 [0.96,0.99] | 6.85E-03 | 0.96 [0.90,1.04] | 3.46E-01 | **0.93 [0.90,0.96]** | **1.74E-06** | 1.00 [0.96,1.05] | 8.63E-01 | 1.41E-04 | 4.32E-04 |
| ***APOE*\*4 Positive** |  |  |  |  |  |  |  |  |  |  |  |  |  |  |  |  |
| rs5924530 | 16841505 | A | G | 37.76% | eXCI | *RBBP7* | 0.96 [0.94,0.98] | 7.15E-05 | **0.94 [0.92,0.97]** | **8.44E-06** | 0.94 [0.91,0.97] | 1.21E-04 | 0.95 [0.91,0.99] | 2.25E-02 | - | 3.94E-02 |
| rs139542115 | 114487884 | T | C | 0.60% | eXCI | *HTR2C* | 1.32 [1.17,1.48] | 4.07E-06 | **1.52 [1.29,1.80]** | **6.46E-07** | 1.58 [1.29,1.94] | 1.08E-05 | 1.42 [1.07,1.88] | 1.61E-02 | - | 2.37E-05 |
| rs146938321 | 137060210 | G | A | 2.38% | Male | *GPR101* | 1.10 [1.03,1.17] | 2.61E-03 | 1.14 [0.76,1.71] | 5.17E-01 | 0.93 [0.83,1.04] | 2.17E-01 | **1.41 [1.22,1.63]** | **4.80E-06** | 4.51E-04 | 2.02E-04 |
| rs554300003 | 145647218 | T | G | 0.50% | rXCI | *SLITRK2* | **1.34 [1.18,1.53]** | **9.11E-06** | 1.50 [0.94,2.40] | 9.19E-02 | 1.19 [0.92,1.52] | 1.77E-01 | 1.92 [1.42,2.60] | 2.07E-05 | - | 2.39E-02 |

### Pleiotropy analyses reveal shared genetic architecture with AD-relevant traits

We next sought to determine to what extent X-linked AD genetic signals were shared, i.e. pleiotropic, with hormone-related or AD-relevant phenotypes, including lipid traits, an AD-relevant set of gray matter structural magnetic resonance imaging (sMRI) and white matter diffusion tensor imaging (DTI) brain phenotypes, and amyloid status from brain PET (**Tables-S5-7**, cf. Methods). These analyses aimed to provide potential mechanistic insights and, most importantly, corroborate the putative biological relevance of X chromosome AD risk loci identified using explorative significance thresholds. To identify pleiotropic signals likely sharing a causal variant, we used genetic colocalization approaches (ABF and SuSiE) on signals associated with at least P<1e-3 (heuristic threshold) in both AD and the paired trait (**Fig.3A**). In total, we identified 63 AD-trait pairs demonstrating evidence of colocalization (posterior probability [PP4]>0.6), spanning 21 independent loci (**Fig.3B, Table-S8**; *APOE*\*4 or sex-stratified signals were filtered to those displaying at least P_het_<0.05 cross-stratum heterogeneity). Importantly, 8 of these loci also passed X-wide significance criteria (**Fig.2B**), including *SLC9A7*, *GLUD2*, *ZNF280C*, and *MAP7D3* in non-*APOE*\*4-stratified analyses, *SMIM10L2B* in *APOE*\*4- subjects, and *RBBP7*, *HT2RC*, and *GPR101* in *APOE*\*4+ subjects, lending further support to their potential AD relevance. Among these, *SMIM10L2B* (*APOE*\*4-, female) and *RBBP7* (*APOE*\*4+, eXCI) displayed the most abundant pleiotropy with AD-relevant traits (**Fig.3B**).

**Figure 3.**
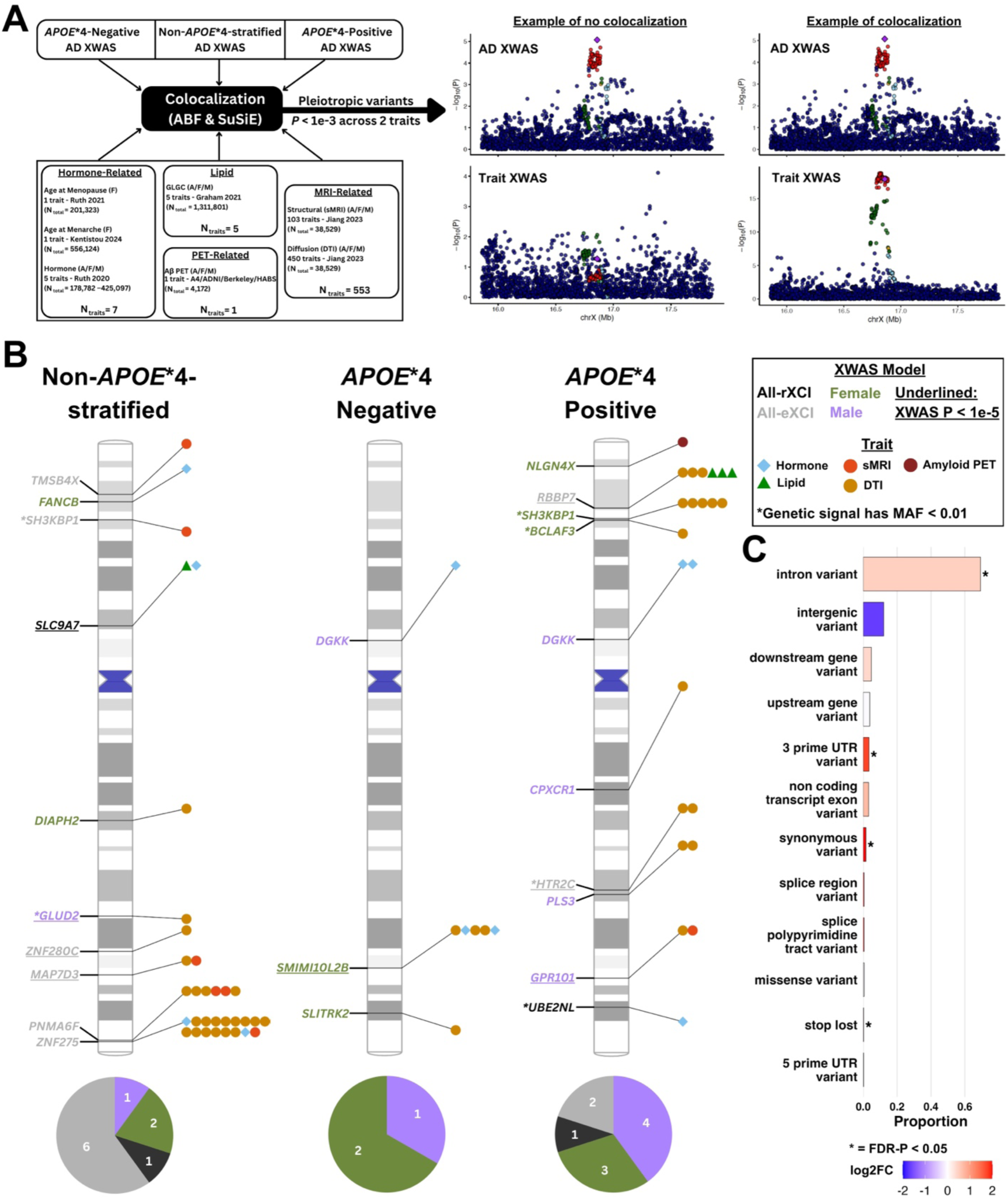
X chromosome Alzheimer’s disease loci display extensive pleiotropy with Alzheimer’s disease relevant traits. **A)** Overview of the multi-trait pleiotropy framework. Left, AD-relevant trait categories evaluated for genetic colocalization with AD XWAS signals. Right, representative regional association plots illustrating absence (left) and presence (right) of colocalization between AD XWAS and trait XWAS signals. **B)** Pleiotropic loci across *APOE*\*4 strata. Phenograms depict X-chromosome positions of loci with evidence of pleiotropy (colocalization PP4>0.6). Gene labels denote the nearest protein-coding gene or a functionally prioritized gene, with text color indicating the AD model or sex with the most significant association for the respective signal. Gene labels marked with an asterisk indicate lead variants with minor allele frequency (MAF) < 0.01. Symbols to the right of each locus indicate the trait(s) showing pleiotropy. Pie charts summarize the distribution of XWAS models or sex-biased associations among pleiotropic loci within each *APOE*\*4 stratum. **C)** Functional consequences of prioritized pleiotropic variants. Bars indicate the proportion of variants annotated to each consequence class, with color representing log2 fold enrichment relative to background. Pleiotropic variants show enrichment for intronic, 3ʹ UTR, synonymous, and stop-lost consequences. Asterisks denote FDR-P < 0.05.

Across the 21 pleiotropic AD loci, genetic overlap was observed with all investigated trait groups, including 6 AD loci overlapping sex-hormone traits such as age-at-menopause, age-at-menarche, testosterone, and sex hormone binding globulin levels, 2 overlapping metabolic traits including HDL cholesterol and triglyceride levels, 6 overlapping gray matter sMRI traits, and 14 overlapping DTI white matter microstructure traits. Similar to X-wide significant loci (**Fig.2B**), the pleiotropic loci were more frequently observed for eXCI and female-specific AD signals (N=14) compared with male-specific AD signals (N=5), suggesting the majority could have relevance to female AD risk mechanisms (**Fig.3B**). Similarly, more signals were found in *APOE*\*4+ (N=10) than *APOE*\*4- subjects (N=3), which may point to an increased number of X-linked genetic interactions with *APOE*\*4, although it should be considered the *APOE*\*4+ group had the smallest sample sizes so more of the identified signals may include false positives (cf. **Tables-S3-4** for demographics). While chance colocalizations could be a concern when performing extensive trait comparisons, 11 out of all 21 pleiotropic loci (and 6 out of 10 *APOE*\*4+ loci) had at least 2 different traits colocalizing with AD, supporting their robustness (**Fig.2**). Importantly, pleiotropic loci were consistent when comparing AD XWAS meta-analyses including and excluding proxy-phenotype samples, indicating their AD associations were not driven by the inclusion of proxy-XWAS (**Fig.S3**). To further confirm the pleiotropic loci did not simply represent chance overlap of non-meaningful signals, but in fact related to functional signals, we evaluated enrichment of variant consequences using Ensembl Variant Effect Predictor (VEP) annotations, comparing prioritized variants (lead pleiotropic variants and those in high linkage disequilibrium; cf. Methods) with the background of all analyzed X chromosome variants (**Fig.3C**, **Tables-S9-10**). Prioritized variants were enriched for intronic annotations, which comprised 69.2% of prioritized variants compared to 49.3% in the background distribution (FDR-P=2.7e-16), while significant enrichment was also observed for less abundant annotations including 3ʹ untranslated region (UTR), synonymous, and stop-lost variants. This observation is notable as intronic and exonic SNPs tend to be depleted on the X chromosome relative to the autosomes^4^. Together, these enrichment patterns support the likely functional relevance of the identified pleiotropic loci.

### X-linked AD loci show extensive overlap with brain white matter microstructure

Given the prominent pleiotropy between AD-associated loci and neuroimaging traits, we next closely examined the implicated brain regions. Across sMRI traits, 7 brain regions showed evidence of colocalization with AD-associated loci (**Fig.4A**), with each region implicated only once by a pleiotropic locus (**Table-S11**). Three of the brain regions were ventricles, which may relate to a brain atrophy-AD link, while two others, the left middle temporal cortex and right isthmus of cingulate gyrus, are also implicated in early AD pathology and atrophy. Compared to sMRI, pleiotropy with DTI traits revealed greater convergence with AD-associated loci. In total, 10 white matter regions demonstrated evidence of colocalization, with 6 overlapping more than one independent AD locus (**Fig.4A, Table-S11**). Notably, the anterior corona radiata (ACR) exhibited the strongest convergence with genetic colocalization at 5 independent AD loci (**Fig.4B**). The splenium of the corpus callosum (SCC) also demonstrated substantial overlap, with 4 independent AD loci colocalizing (**Fig.4B**). Interestingly, all SCC-associated signals originated from *APOE*\*4+ AD XWAS models (and included the newly identified X-wide significant *RBBP7* eXCI locus), suggesting that X-linked genetic effects influencing this white matter tract may be particularly relevant in the context of *APOE*\*4-related AD risk (cf. Discussion).

**Figure 4.**
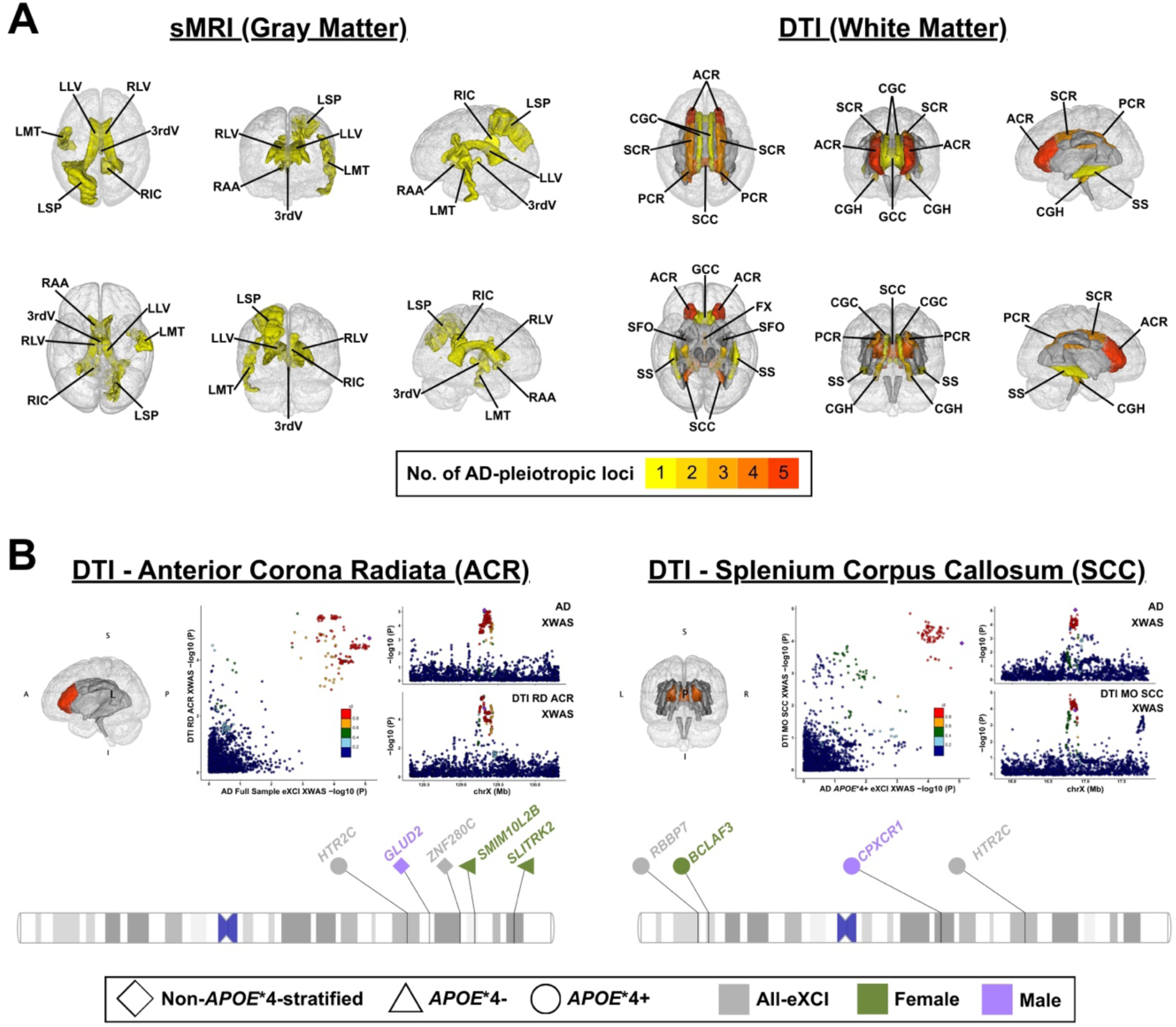
Alzheimer’s disease X-chromosome pleiotropic loci converge in white matter regions. **A)** Brain imaging maps showing the number of independent AD signals that colocalize with gray matter (sMRI, left) and white matter (DTI, right) traits in respective areas. Colors indicate the number of independent AD signals colocalizing with each region. For each set of brain masks, views are shown clockwise from top left: superior, anterior, left, right, posterior, and inferior. **B)** DTI white matter regions with the highest degree of AD genetic overlap. Left, anterior corona radiata (ACR), which colocalized with five independent AD XWAS signals across multiple models and *APOE*\*4 strata. Right, splenium of the corpus callosum (SCC), which colocalized with four independent AD XWAS signals, all within the *APOE*\*4+ stratum. For each region, the brain mask (top left), an example regional association plot illustrating AD–DTI signal overlap (top right), and a phenogram summarizing implicated loci (bottom) are shown. Phenogram symbols denote *APOE*\*4 strata and colors indicate XWAS models. ***Abbreviations:*** *3rdV, 3^rd^ Ventricle; ACR, anterior corona radiata; CGC, cingulum (cingulate gyrus); CGH, cingulum (hippocampus); FX, fornix; GCC, genu of corpus callosum; LLV, left lateral ventricle; LMT, left medial-temporal lobe; LSP, left superior parietal lobe; No., number; PCR, posterior corona radiata; RAA, right accumbens area; RIC, right isthmus cingulate; RLV, right lateral ventricle; SCC, splenium corpus callosum; SCR, superior corona radiata; SFO, superior fronto-occipital fasciculus; SS, sagittal stratum*.

### Functional genomics analyses prioritize candidate genes at X-linked AD loci

For all AD loci nominated through XWAS (**Fig.2**) and pleiotropy (**Fig.3**), we sought to identify candidate causal genes and those loci with the overall most robust genomic support (**Fig.5**). First, we performed genetic colocalization analyses with multi-tissue quantitative trait locus (QTL) datasets that have mapped variant associations with gene expression (eQTL), transcript splicing (sQTL), and protein levels (pQTL), collectively referred to as xQTL (**Table-S12**). Notably, these included novel brain cell type-specific eQTL data through reprocessing of a recent, large-scale (N=424) single-nucleus RNA-seq dataset from the dorsolateral prefrontal cortex (DLPFC; cf. Methods). In total, for 8 out of 26 evaluated loci we were able to prioritize 30 genes showing evidence of colocalization (PP4>0.6), with *SLC9A7*, *ZNF280C*, *MAP7D3*, and *RBBP7* being most numerously prioritized across QTL datasets (**Fig.5**, **Fig.S4**, **Table-S13)**. For the remaining 18 loci, we nominated candidate causal genes by mapping intronic signals (N=9) to the gene in which they reside and intergenic signals (N=9) to the nearest protein-coding gene (distance in base-pair positions to transcription start site: median=87,240, min=26,215, max=606,045; **Table-S14**). While less robust than evidence of genetic colocalization, this is established to likely implicate the causal gene when the association signal is proximal to the gene transcription start site^39^. The resultant set of 48 candidate genes was subsequently evaluated for differential expression of genes (DEG) between AD cases and controls using bulk transcriptomic datasets from 8 AD-relevant brain regions to further provide orthogonal support for their potential AD relevance (**Table-S15**; DEG analyses for respective genes were sex-matched to their related AD XWAS signals, cf. Methods). We then built support level scores for each gene by integrating whether their underlying genetic signals were associated through (i) AD XWAS (P<1e-5) and/or (ii) multi-trait pleiotropy, and the extent to which they were supported by (iii) xQTL colocalization, and (iv) DEG (cf. Methods). A comprehensive overview of findings is presented in **Table-S16**; however, we prioritized the 20 loci with stronger genetic support (AD XWAS P<1e-5 or at least 2 pleiotropic trait colocalizations) and present their top candidate genes in **Fig.5A**. Interestingly, among the 20 candidate genes, significant DEG findings were most abundant in the anterior cingulate cortex (ACC), with 15 showing FDR-P<0.05 and 11 remaining significant after Bonferroni correction for the number of evaluated brain regions (**Fig.5A**). To determine whether this was not a chance observation, we compared the DEG effect sizes of prioritized genes to a null distribution derived from all X chromosome genes tested in the corresponding brain area (cf. Methods). This confirmed the prioritized genes displayed significant DEG enrichment in the ACC (*P*=0.03; **Fig.5B**). Notably, the ACC is connected to frontal cortical networks through the ACR, a white matter tract that exhibited strong convergence with AD loci in the pleiotropy analyses (**Fig.4**), further corroborating that these brain areas may be implicated in X-linked AD susceptibility.

**Figure 5.**
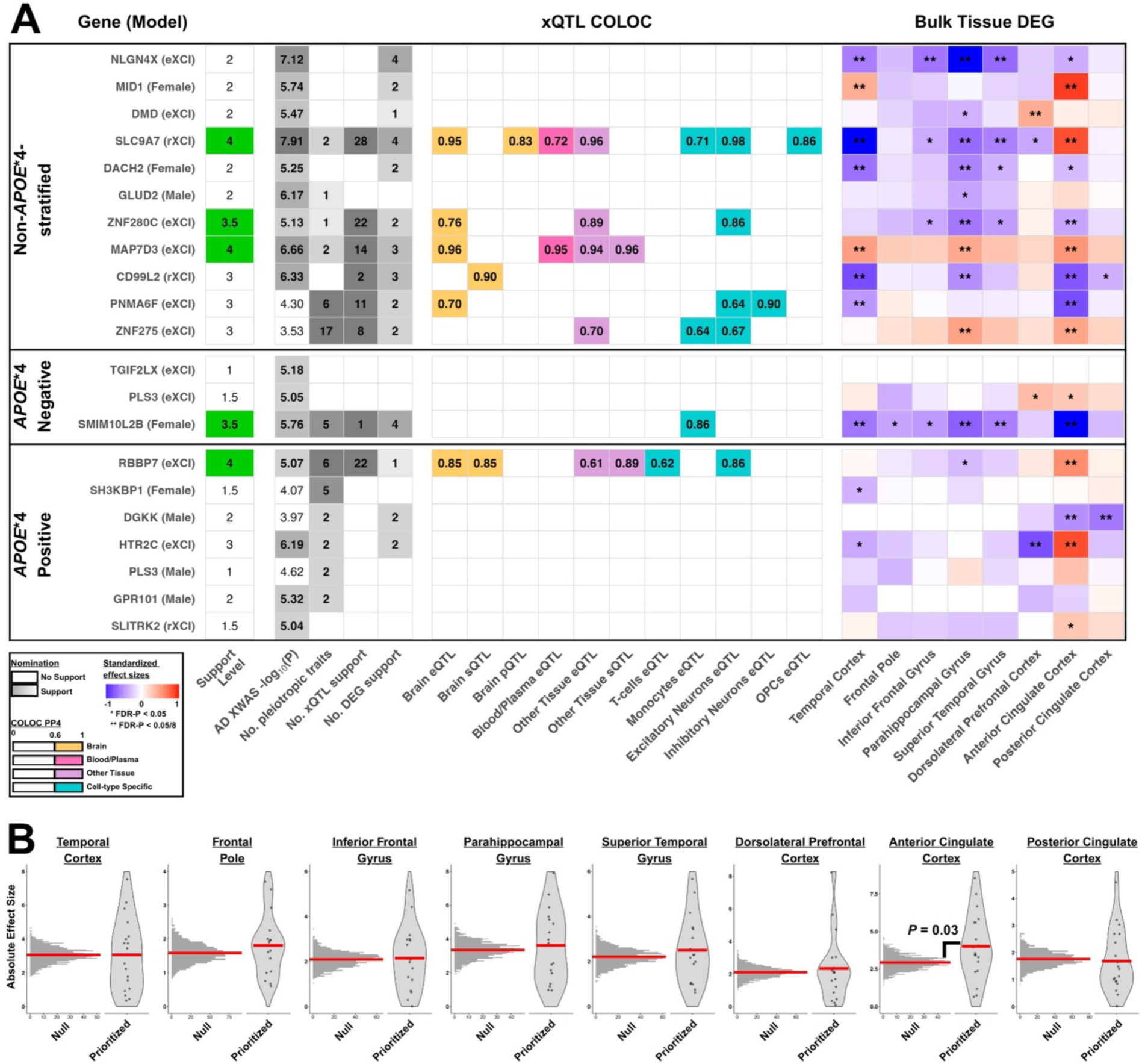
Gene prioritization and enrichment of brain area-specific case-control differential expression. **A)** The gene prioritization matrix summarizes multi-layer support for the top prioritized gene at respective AD XWAS signals across *APOE*\*4 strata; 20 loci with stronger genetic support are shown, while results for all loci and nominated genes are available in Table-S16. The support level column indicates the number of independent evidence sources with higher values denoting greater support (cf. Methods). Additional columns summarize AD XWAS signal significance (−log_10_P), number of pleiotropic traits, number of xQTL colocalizations (PP4≥0.6), and number of brain regions (out of eight) showing differential expression of genes (DEG) across AD cases and controls. Middle columns display xQTL colocalization posterior probabilities (PP4), with colored cells indicating evidence of colocalization in respective tissues or cell-types. Right columns show bulk DEG across eight brain regions; red indicates higher expression in AD cases and blue indicates lower expression. Asterisks denote DEG significance (*FDR-P < 0.05; **FDR-P < 0.05/8). **B)** DEG patterns of prioritized genes are shown across eight brain regions. In each panel, the histogram (left) represents the null distribution of absolute DEG effect sizes for randomly sampled X-chromosome genes, and the violin plot (right) summarizes the distribution of absolute DEG effects for prioritized genes. Significant enrichment of DEG was observed in the anterior cingulate cortex (ACC, P=0.03).

### Top X-linked AD loci and genes

Among the 20 prioritized genes at their respective loci, *SLC9A7*, *ZNF280C*, *MAP7D3*, *SMIM10L2B*, and *RBBP7* obtained the highest support levels (scores >3). They were supported across all evidence layers marking them as the 5 most robust genes with potential relevance to AD (**Fig.5A**). Notably, the genetic signals underlying these genes were all implicated to causally regulate the gene’s expression or abundance levels in brain bulk tissues or excitatory neurons (**Fig.5A**), further supporting their relevance to AD and the brain imaging traits associated at these loci. Among them, *SMIM10L2B* and *RBBP7* related to novel X-linked loci. *RBBP7* was of particular interest since the underlying genetic signal indicated an eXCI mechanism that confers protection against AD specifically in *APOE*\*4 carriers (**Fig.6**). Importantly, multiple genes at this locus, including *RBBP7*, have extensive prior evidence of undergoing eXCI (**Fig.6**, **Table-S17**), corroborating the validity of the AD XWAS association model. The AD signal was pleiotropic with several lipid traits and DTI metrics, including in the SCC, and regulates *RBBP7* expression in excitatory neurons from DLPFC. Further inspection of brain DLPFC single-cell eQTL data confirmed the AD-protective index variant was associated with increased *RBBP7* expression solely in excitatory neurons and not in any other cell-type, while independent single-cell ATAC-seq data derived from multiple brain areas additionally confirmed the AD index variant, rs5924530, fell within a chromatin accessibility peak for excitatory neurons (**Fig.S5**). These findings corroborate the gene’s putative causal mechanism likely manifests in excitatory neurons. For the remaining 4 top genes, detailed result summaries equivalent to **Fig.6** are provided in **Fig.S6-9**, while mechanistic interpretations of the findings are presented in the discussion.

**Figure 6.**
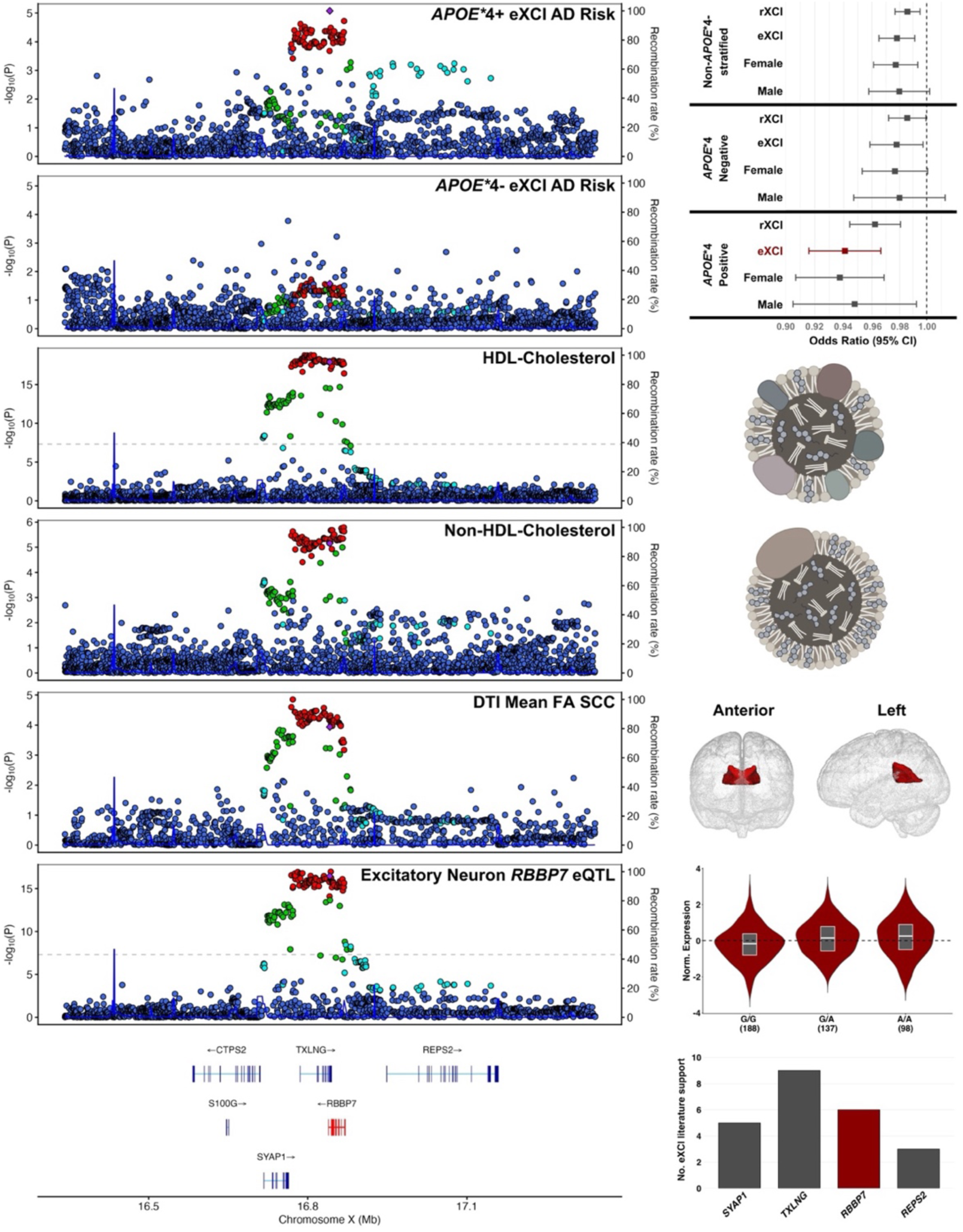
Escape gene *RBBP7* is associated with reduced Alzheimer’s disease risk in *APOE*\*4 carriers. **Left.** Stacked locus zoom plots highlight from top to bottom the *APOE*\*4+ biased eXCI AD association signal at the *RBBP7* locus, followed by its pleiotropy with peripheral lipid and brain white matter traits, and QTL colocalization evidence implicating the signal may causally regulate *RBBP7* expression levels in excitatory neurons (supporting data are in Fig.S5). Bottom depicts gene tracks at the locus. **Right.** Panels contain supporting or visual information matching locus zoom plots on the left. From top to bottom, forest plot displays effect sizes of the AD index variant across all models and strata, schematics indicate lipid traits or implicated brain area, violin plots show the AD protective allele is associated with increased *RBBP7* expression in excitatory neurons, and the bottom histogram indicates the number of literature-supported reports of eXCI for genes at this locus, corroborating the AD eXCI model validity at this locus.

## Discussion

We performed the most comprehensive, to date, investigation of the X-linked genetic architecture of AD and for the first time mapped X-linked loci associated with AD in an *APOE*\*4-dependent manner. Recognizing that XWAS face significant power challenges, we explored genetic signals below conventional significance criteria, using pleiotropy and functional genomics analyses to corroborate them, and built an atlas of 26 potential X-linked AD loci and their candidate genes, as well as the most likely genetic model, sex, and *APOE*\*4 stratum that drives their association. While these loci and genes are unlikely to all be causal to AD, this resource will be important in guiding future experimental studies and enable anchoring of novel findings to human genetics support established here. Importantly, by composing genomic support scores for nominated genes, we guide gene prioritization and propose *RBBP7, SMIM10L2B, SLC9A7, ZNF280C*, and *MAP7D3* as the most robust candidate AD genes. Additionally, we found that several AD loci and candidate genes converged on the anterior corona radiata, splenium of the corpus callosum, and anterior cingulate cortex, implicating these brain regions as potentially important to X-linked AD etiology.

Our X-linked AD loci contained many novel associations but also overlapped all loci previously reported by Belloy *et al*. (as expected) as well as several reported by *Le Borgne* et al.^27,28^. Sensitivity analyses confirmed their associations were not driven by the inclusion of proxy XWAS, supporting the robustness of our findings. Notably, many AD genetic signals were conferred by eXCI models or female strata. While eXCI signals do not inherently reflect larger variant effect sizes in women, the anticipated double gene dosage in women relative to men is posed to contribute to female-biased disease mechanisms^19^. Our findings thus align with converging reports that more sex-biased genetic loci and genes relate to female-biased AD risk mechanisms and are also consistent with the anticipated larger female-biased role of the X chromosome in AD risk and resilience^7,8,12,13,17^. Another striking observation was the convergence of AD pleiotropy with white matter metrics, which was most extensive in the anterior corona radiata (ACR) and splenium of the corpus callosum (SCC), and the prioritization of the anterior cingulate cortex (ACC) through case-control DEG enrichment. The ACR and SCC are major white matter tracts respectively involved in frontal cortical connectivity. The ACR is a complex white matter region with connections to several inter- and intra-hemispheric white matter tracts, whereas the SCC has projections to several AD-relevant parietal and temporal brain regions, supporting their potential relevance to AD^40–43^. The genetic loci that converged onto the ACR and SCC represented a mix of eXCI, male, and female-biased AD associations, suggesting these regions might contribute to sex-dimorphic AD pathobiology. Notably, the 4 loci that converged on the SCC all displayed *APOE*\*4+ biased effects. Increasing evidence has implicated the relevance of white matter changes in AD pathology and cognitive decline and has importantly shown these changes can be *APOE*\*4 and sex-dependent^40,41,44–47^, including specific observations of sex-by-*APOE*\*4 interactions in splenium and ACR-related tracts^40^. Importantly, *APOE*\*4 has been shown to disrupt lipid metabolism in oligodendrocytes and impair myelin maintenance, potentially rendering highly myelinated tracts vulnerable to degeneration^47,48^. A recent study further linked increased glucose metabolism in the corona radiata to cognitive decline and specifically suggested this may represent a compensatory mechanism^49^. Notably, the ACR forms a major white matter pathway connecting the ACC with prefrontal and subcortical regions involved in cognitive control, attention, and emotional regulation^50,51^. Early network hyperactivity in the ACC has been reported in both humans and mice and has been proposed as either an early driver of AD or a compensatory mechanism^52–54^. Our case-control DEG findings for ACC included several genes (*MID1*, *SLC9A7*, *RBBP7*, *HTR2C*) showing AD associations that markedly differed from those observed in other brain areas (**Fig.5A**), further corroborating the ACC may contribute to AD in a distinct manner. Altogether, our findings emphasize the need for additional *APOE*\*4 and sex-informed research into the role of white matter changes and the ACR, SCC, and ACC regions in AD. While our findings are anchored to X-linked genetic loci, we are not aware of equivalent studies on the autosomes, such that future work will need to confirm whether these observations are uniquely X-linked or also extend to autosomal risk loci.

Among the 5 top X-linked candidate AD genes, *RBBP7, MAP7D3, and ZNF280C* were found under an eXCI model, while *SLC9A7* was found under an rXCI model, which aligns directly with prior reports for these genes (**Table-S17**). Notably, *RBBP7,* associated in *APOE*\*4 carriers, and *MAP7D3*, associated independent of *APOE*\*4 status, both appeared to be directly relevant to tau pathology. Prior work found that reduced *RBBP7* (*Retinoblastoma-binding protein 7*) expression in bulk tissue and neurons from the human medial-temporal gyrus is associated with AD and tau pathology and provided experimental evidence that loss of *RBBP7* promotes tau acetylation and downstream neurodegeneration, whereas restoration of *RBBP7* activity suppresses tau-mediated neurotoxicity^55^. This aligns with the AD index variant at this locus being protective against AD risk while increasing *RBBP7* expression levels specifically in excitatory neurons derived from DLPFC. Prior studies have reported that females display higher tau pathology than men, especially female *APOE*\*4 carriers, and may be more resilient to tau burden during earlier stages of the disease^14,16,17,30,32^. *RBBP7* may tie into this, since this escape gene would be expected to have double dosage in women relative to men which in turn could contribute to female-biased protection against tau burden. We also observed that DEG support for *RBBP7* was most notable in the ACC and indicated *RBBP7* levels are higher in AD cases versus controls, which appears opposite to the genetic effects of the AD index variant and prior experimental findings. However, this could reflect compensatory mechanisms or the fact that the genetic effect on bulk tissue or excitatory neuron *RBBP7* expression was only observed in DLPFC. It should also be considered that we found evidence of the AD index variant affecting splicing of *RBBP7* in bulk brain tissue (**Fig.S4**), which may represent another mechanism linking it to AD risk. Further, *MAP7D3* (microtubule-associated protein 7 Domain Containing 3) has been reported to share its binding site with tau on microtubules^56^, meaning that altered *MAP7D3* levels could directly impact tau dynamics. The AD index variant at this locus was protective against AD risk while increasing *MAP7D3* expression levels in bulk DLPFC (**Fig.S6**), whereas DEG findings indicated *MAP7D3* levels were not significantly associated with AD case-control status in DLPFC but showed higher levels in AD cases versus controls in the temporal cortex, parahippocampal gyrus, and ACC. Beyond relevance to tau pathology, *SLC9A7* and *SMIM10L2B* have been linked to amyloid. *SLC9A7* was the main gene reported in Belloy *et al*. where a detailed description specified a mechanism by which *SLC9A7* could regulate pH homeostasis in Golgi secretory compartments and impact downstream effects on Aβ^27^. Extending on Belloy *et al*., we confirm that the AD-risk variant at this locus was associated with increased expression of *SLC9A7* not just in brain DLPFC bulk tissue, but more specifically in DLPFC excitatory neurons and OPCs. One hypothesis derived from prior experimental studies is that higher *SLC9A7* levels increase AD risk^27^, which aligns with the observed genetic effect on *SLC9A7* levels in DLPFC and its DEG association with AD status in the ACC, although opposite associations were found for DEG in other brain regions. Another relevant, novel insight was provided from pleiotropy analyses, which suggested that the *SLC9A7* locus in fact appeared to comprise two distinct signals in partial LD, with the main rXCI signal colocalizing with HDL-cholesterol and *SLC9A7* expression, while a second eXCI signal colocalizes with testosterone levels in women (**Fig.S7**), suggesting that multiple regulatory mechanisms at this locus may contribute to AD susceptibility. Moving on, the candidate gene *SMIM10L2B* was nominated at an *APOE*\*4- female-specific XWAS signal. This locus was notably pleiotropic with bioavailable and total testosterone levels which could relate to its sex-biased effect in AD. Colocalization analyses further identified the AD signal related to regulation of *SMIM10L2B* expression in monocytes (**Fig.S8**). Higher levels of *SMIM10L2B, a.k.a. small integral membrane protein 10–like protein 2B* (*SIL2B*), in macrosomes secreted by microglia was recently implicated to inhibit Aβ fibrillation and it was proposed that *SIL2B* overexpression in macrosomes holds therapeutic potential for AD^57^. Consistently, in bulk brain transcriptomic analyses, *SMIM10L2B* expression appeared to be downregulated in AD cases versus controls across 6 different brain regions. In contrast, the AD index variant at this locus was protective against AD while decreasing *SMIM10L2B* expression levels in monocytes, although no genetic impact was inferred for microglial *SMIM10L2B* expression nor is it clear whether this would directly relate to *SMIM10L2B* levels in secreted macrosomes. Finally, for *ZNF280C* (*Zinc Finger Protein 280C*; **Fig.S9**), we did not infer a clear mechanistic link to AD based on prior literature, although its role as a transcription factor may yet tie it to AD-relevant genes. While for the 5 candidate genes there tended to be differences in association patterns across genetic effects and postmortem gene expression across brain regions, it should be considered these effect estimates could often not be compared in the same tissue context and may further be decoupled due to late-stage disease-associated changes, reactive processes, or compensatory mechanisms in postmortem samples. The DEG analyses thus mainly provide orthogonal support to corroborate the genetically prioritized risk genes. Altogether, for our 5 top candidate genes, gaps remain to understand their exact mechanistic links to AD, but their strong genomic support and AD-relevant biological precedent strongly encourage experimental follow-up.

The large number of pleiotropic loci we observed is consistent with expectations of wide-spread pleiotropy at GWAS loci for complex traits^58,59^. Importantly, pleiotropy does not imply causality and many variant effect estimates at our AD-trait pair loci did not indicate straightforward causal relationships nor alignment of sex biases (**Table-S8**). Interpretation of effect estimates at pleiotropic loci is challenging because of differences in study recruitment designs and biases, potential disconnection between effects on earlier life-course and intermediate traits versus late-life neurodegenerative disease risk, uncertain phenotypic impact (e.g. white matter DTI traits) and, in case of the X chromosome, potential differences in trait sex dimorphism and association power (e.g. male XWAS usually have more power than female XWAS). Rather, the insights gathered here enable hypothesis generation of how implicated loci may impact AD risk or alternatively reflect non-causally shared biology. Per illustration, our findings and prior work prioritize a neuronal mechanism through which *RBBP7* could affect tau-related AD pathology^55^. However, this locus also displayed pleiotropy with peripheral cholesterol and triglyceride levels, as well as regional brain white matter microstructure. Lipid metabolism is an important aspect of AD pathobiology, suggesting that other risk mechanisms may relate *RBBP7* to AD risk. Even if such alternative mechanisms would not causally contribute to AD etiology, changes in lipid metabolism and white matter integrity with aging could potentially alter *RBBP7* availability to indirectly impact AD risk. Another possibility is that this locus also regulates expression of *TXLNG*, which has been linked to insulin resistance^60^, such that other causal genes and mechanisms may exist. The pleiotropic findings thus provide a more comprehensive picture to elucidate potential causal mechanisms at the *RBBP7* locus. Another notable showcase of insights enabled through pleiotropy was observed for the *DGKK* locus. Although *DGKK* did not reach the highest genomic support level, it stood out for displaying the strongest *APOE*\*4 heterogeneity (P_Het_=1.1e-7), with an AD risk-increasing effect in *APOE\**4- men and risk-decreasing effect in *APOE*\*4+ men, while demonstrating genetic overlap with testosterone levels and age-at-menarche (**Fig.S10**). Interactions with sex hormone biology could explain the male-biased effect at this locus while a recent report additionally suggests testosterone levels may be associated with cognition in an *APOE*\*4 dependent manner^61^. Further investigation of this locus may thus shed light on how testosterone biology could affect *APOE*\*4 and sex-dependent risk for AD^62^.

Several limitations should be considered when interpreting our findings. First, our XWAS focused on subjects of European ancestry, and such findings may not be generalizable across populations. Future cross-ancestry XWAS will be important. Additionally, some of the sex or *APOE*\*4 biased heterogeneous effect estimates may partially reflect sex-specific or *APOE*\*4-specific differences in prevalence or underlying environmental factors rather than true biological heterogeneity. Further, most AD cases were determined through proxy or health-registry phenotypes, which may reduce specificity to AD versus general dementia. Together with the fact that XWAS face power challenges and only one of our associated loci passed genome-wide significance, further studies will be needed to corroborate our genetic discoveries. As noted earlier, one of our core aims was to provide a resource of human genetic insights on X chromosome to steer future studies, but it is expected that several loci will not actually be causal to AD. Further experimental validation is warranted.

In summary, we provide the most comprehensive investigation of the X chromosome’s contributions to AD. The implicated candidate genetic loci, causal genes, pleiotropic insights, and AD-relevant brain areas, will directly guide future follow-up studies to elucidate X-linked mechanisms in AD. Importantly, our findings support the crucial need for more sex and *APOE*-aware research to advance our understanding of AD and identify novel therapeutic targets.

## Online Methods

### Ethics declaration

Participants or their caregivers provided written informed consent in the original studies. The current study protocol was granted an exemption by the Washington University Institutional Review Board because the analyses were carried out on “de-identified, off-the-shelf” data; therefore, additional informed consent was not required. The FinnGen ethics statement is available in the supplement (Nr HUS/990/2017).

### AD XWAS samples, phenotypes, and quality control

Detailed information is provided in the **Supplemental Methods**. Case-control, family-based, and longitudinal AD genetic cohorts from the Alzheimer’s Disease Genetics Consortium (ADGC) and Alzheimer’s Disease Sequencing Project (ADSP; release 3) were obtained through public data repositories and included genotype data generated using SNP microarrays and whole-genome sequencing (**Tables-S1-2**)^37,38^. These cohorts contributed clinically diagnosed AD cases including a subset of pathology-confirmed diagnoses (∼40%; **Table-S3**)^63–65^. Analyses in UK Biobank (UKB), FinnGen (R12), and the Million Veteran Program (MVP) were based on SNP microarray data processed through cohort-specific pipelines^66–69^. To maximize statistical power in FinnGen, we used the broad AD phenotype definition without age restrictions for controls, consistent with prior autosomal AD association studies^70^. UKB contributed registry-confirmed AD cases as well as proxy Alzheimer’s disease and dementia (ADD) phenotypes based on parental history; FinnGen contributed registry-confirmed AD cases; and MVP contributed both registry-confirmed ADD cases (MVP-1) and proxy ADD phenotypes (MVP-2).

Genetic data from the ADGC and ADSP underwent extensive quality control (QC) and were imputed to the TOPMed reference panel (cf. Belloy *et al*.^27^). X-chromosome-specific QC procedures were applied following previously described approaches (cf. **Supplemental Methods**). Genetic data from UK Biobank, FinnGen, and the Million Veteran Program were processed according to each cohort’s established genotype QC and analysis pipelines^66–69^. For association analyses, we included non-Hispanic White individuals of European ancestry with XX or XY karyotypes concordant with self-reported sex and ages >60 years (>18 years and median=63 in FinnGen; **Table-S3**). Variants were filtered using cohort-specific minor allele frequency (MAF) thresholds, corresponding on average to a MAF of approximately 0.05% (cf. Belloy *et al*.^27^).

### AD XWAS approach and analyses

XWAS were restricted to non-pseudoautosomal regions since pseudoautosomal regions had insufficient variant coverage across ADGC SNP microarrays. XWAS evaluated case-control logistic regression models for AD risk, adjusting for age, sex (when applicable), technical covariates, and genetic principal components capturing population structure. Mixed-model association methods were used to account for relatedness in cohorts where appropriate, including ADGC, ADSP, and UKB (LMM-BOLT v2.4)^71^, as well as FinnGen (Regenie)^67^. Cohort-specific association results were subsequently combined across respective models or strata (cf. below) using fixed-effects inverse-variance weighted meta-analysis implemented with GWAMA. To increase specificity for clinically diagnosed AD (rather than ADD), meta-analysis results were restricted to variants with association results available in ADGC cohorts. The *APOE*\*4 stratified analyses grouped *APOE*\*2/2, *APOE*\*2/3, and *APOE*\*3/3 carriers as *APOE*\*4- subjects and *APOE*\*3/4, *APOE*\*3/4, and *APOE*\*4/4 carriers as *APOE*\*4+ subjects while further adjusting regression models for *APOE*\*2 and *APOE*\*4 dosage (as applicable) within the respective strata. *APOE*\*4 stratified XWAS were not available from FinnGen. Additional details on association models are provided in the **Supplemental Methods**.

Genotype encoding and sex-aware modelling strategies are visualized in **Fig.1B** and further detailed here. When modelling random X chromosome inactivation (rXCI) in joint male and female analyses, it was assumed that in females a single allele copy corresponds to a 50% probability of being active due to rXCI^72^. Therefore, to render effect estimates on the same scale, genotypes were encoded as 0/2 in males (XY) and 0/1/2 in females (XX). In the UKB and MVP cohorts, a large proportion of cases correspond to proxy phenotypes based on parental history of ADD. This proxy design has extensively been shown to capture autosomal AD genetic associations, and we previously successfully adapted it for AD XWAS^27,70,73^. Our approach is detailed in Belloy *et al*. and allows effect estimates and standard errors to be calibrated consistent with genotype encoding 0/2 in males (XY) and 0/1/2 in females (XX)^27^. Next, to model escape from X chromosome inactivation (eXCI), it was assumed that in females a single allele copy corresponds to a 100% probability of being active (full escape), such that males should be encoded as 0/1 to render effect estimates on the same scale. Since it was not possible to directly implement this eXCI encoding in joint male and female analyses across the different biobanks and analytical strategies we employed, we leveraged an alternative approach to eXCI modelling. Specifically, male-derived XWAS summary statistics can be changed directly from 0/2 to 0/1 encoding by doubling beta coefficient and standard errors (or vice versa by halving them). In line with the visual illustration in **Fig.1B**, we assumed that male X-linked effect estimates on the 0/1 scale will line up with female X-linked effect estimates if there is eXCI at a respective locus, but not if there is rXCI. Leveraging this framework, our eXCI XWAS were performed through random effects meta-analyses of male 0/1-encoded XWAS with female XWAS, which leads to increased association significance for loci displaying evidence of eXCI versus decreased significance for loci displaying evidence of rXCI. Finally, the male-stratified and female-stratified were respectively conducted using 0/2 and 0/1/2 genotype encoding, but for the purpose of cross-sex eXCI meta-analyses and reporting of effect estimates, male XWAS findings were converted to the 0/1 genotype scale. Male AD XWAS results were available across cohorts, except MVP-2 which uses paternal AD phenotypes as a proxy for male-biased AD risk. Since males cannot inherent X chromosome from their fathers and the fraction of female subjects in MVP is very small (<4%, **Table-S3**), we did not pursue these analyses.

### AD XWAS locus and signal selection

To identify independent association signals, variants from each of the 12 AD XWAS were first filtered to those reaching X-wide significance (P<1e-5). Within each XWAS, these variants were LD-clumped at R2<0.01 (1Mb window) using PLINK2^74^ and LD data from the ADGC cohort. To determine whether sex or *APOE*\*4 stratified signals displayed evidence of sex- or *APOE*\*4-biased effect estimates, cross-stratum heterogeneity was assessed using Z-tests, calculated as Z = (Beta_1_ – Beta_2_)/√ (SE_1_^2^ + SE_2_^2^) with two-sided P-values obtained from the standard normal distribution using pnorm in R. Variants demonstrating FDR-P<0.05 on these heterogeneity tests were classified as sex- or *APOE*\*4-biased signals. When multiple lead variants in LD (R2>0.01) were identified across different XWAS, these were clumped into a single representative signal which was labelled with the most significant model or sex group. Each independent lead variant was then assigned to the top gene prioritized through functional genomics analyses (cf. below) or, if not available, the nearest protein-coding gene using GENCODE v47.

### Multi-trait X-chromosome pleiotropy analyses

Genetic pleiotropy was assessed between all AD XWAS and a set of sex hormone-related and AD-relevant phenotypes from European ancestry XWAS, which included both sex-stratified and non-stratified results. A comprehensive overview of the 566 evaluated traits and their public identifiers is provided in **Table-S5**. Brain imaging XWAS for sMRI and DTI traits were available from Jiang *et al*. and were derived from 38,529 subjects from UKB^6^. Briefly, sMRI data were processed with Advanced Normalization Tools to derive regional brain volume and cortical thickness, using Desikan-Killiany-Tourville labelling, while surface area values were directly available from UKB and were derived using the Desikan-Killiany parcellation scheme with FreeSurfer. DTI metrics included AD (axial diffusivity; the eigenvalue of the principal direction), FA (fractional anisotropy; related to directionality), MD (mean diffusivity; magnitude of absolute directionality), MO (mode of anisotropy; third moment of a tensor), and RD (radial diffusivity; average of the eigenvalues of secondary diffusion directions). For each white matter tract, these metrics were averaged across voxels, and additionally an average across all metrics was computed, providing 6 traits per white matter tract. Additionally, for each white matter tract and DTI metric, functional principal components analysis was used to extract the top 5 PCs for that respective tract and metric, adding 25 additional traits per white matter tract (for a total of 30 traits per region). All available sMRI and DTI brain regions were then restricted to a set of AD-relevant brain regions to enable targeted analyses (**Tables-S6-7**). In gray matter regions, we prioritized analyses to primarily temporal, frontal, and parietal areas^75–77^. Similarly, we prioritized our DTI analyses to AD-relevant white matter areas^40–43,78–81^, including the anterior corona radiata, cingulum, fornix, genu/splenium of the corpus callosum, posterior corona radiata, superior corona radiata, superior fronto-occipital fasciculus, and sagittal stratum. Further, Amyloid PET data with matching genetic samples for 4,172 subjects were processed from the A4, ADNI, Berkeley, and HABS cohorts. Both amyloid positivity status and scalar amyloid values for an AD-relevant brain composite were derived and used to generate novel amyloid XWAS (cf. **Supplementary Methods** for additional details). XWAS of blood lipid levels were available from Graham *et al*. and were derived from 1,311,801 subjects^82^. Finally, sex hormone related trait XWAS were available for age-at-menopause^83^, age-at-menarche^84^, age-at-voice breaking^85^, total and bioavailable testosterone levels, and SHBG levels^86^, with sample sizes ranging ∼150,000-550,000^83–86^.

To identify potential pleiotropic loci, each of the 12 AD XWAS was intersected with each of the 566 evaluated traits to identify variants associated with at least P<1e-3 in both paired traits. When multiple variants met this criterion within a region, LD clumping was performed using PLINK2^74^ to identify representative lead variants per locus. For each representative variant, genetic colocalization (COLOC) was conducted within a ±1Mb window using both ‘coloc.abf’ with default per-variant priors, which assumes a single independent signal at the locus, and ‘coloc.susie’, which incorporates SuSiE (Sum of Single Effects) regression to resolve loci with multiple independent signals (coloc R package v.4.2.1)^87,88^. Signals were prioritized if they demonstrated evidence of a shared causal variant (posterior probability for hypothesis 4, PP4>0.6). Post hoc, pleiotropic AD signals that were observed in a respective sex or *APOE*\*4 stratum were evaluated for cross-stratum effect heterogeneity using the same approach as described above in “XWAS locus and signal selection”. Signals demonstrating P<0.05 on these heterogeneity tests were classified as sex- or *APOE*\*4-biased signals while others were excluded and not reported. Additional details are provided in the **Supplemental Methods**.

### Variant annotation and functional consequence enrichment

The Ensembl Variant Effect Predictor (VEP; release 115)^89^ was used to annotate consequences of all variants investigated in AD XWAS. Because many variants were assigned multiple consequence terms, each variant was collapsed to a single annotation corresponding to the most severe VEP-defined consequence. For functional consequence enrichment analyses, the background set comprised all variants that passed QC and were part of AD XWAS while prioritized variants were defined as the lead variant at each pleiotropic AD locus together with variants in high LD (R2>0.8) and associated with at least P<1e-3. Enrichment of VEP consequence categories among prioritized variants relative to the background was assessed using one-sided hypergeometric tests. Proportions of variants per consequence category were compared between sets and enrichment was summarized as log₂ fold change of prioritized versus background proportions. P-values were adjusted using Benjamini-Hochberg FDR correction.

### Visualization of MRI trait colocalization across brain regions

Brain MRI traits showing evidence of genetic colocalization with AD XWAS signals were summarized by counting the number of unique AD signals colocalized at a given brain region. Frequency maps were visualized using brain masks rendered in Mango (Multi-image Analysis GUI)^90^.

### Candidate gene nomination – QTL colocalization and gene proximity

Consistent with prior AD GWAS and XWAS^13,27,70^, we sought to identify candidate causal genes at X-linked AD loci using genetic colocalization with QTL datasets containing *cis* genetic association signals for a wide range of molecular traits across diverse tissues and cell types. These analyses were focused on neurologically relevant tissues such as brain, plasma/blood, brain cell-type specific data, and peripheral immune cells (monocytes, macrophages, and T-cells), as well as other tissues from across the body available through GTEx v8^91^ (**Table-S12**). The molecular traits, collectively referred to as “xQTL”, included gene expression (eQTL), protein abundance (pQTL), and transcript splicing (sQTL). Brain cell-type specific X chromosome eQTL data were newly generated using pseudobulk expression data from DLPFC tissue of the ROSMAP cohort, available from Fujita *et al*^92^. Matching whole genome sequencing data (N=423 subjects) was subjected to X chromosome specific quality control^27^. Pseudobulk gene expression data for seven cell types–excitatory neurons, inhibitory neurons, astrocytes, microglia, oligodendrocytes, oligodendrocyte progenitor cells (OPCs), and endothelial cells–were obtained and processed with low-expression filtering, TMM normalization, voom transformation (logCPM), ComBat batch correction, and quantile normalization. X-chromosome *cis* eQTLs (2Mb window) were mapped using PLINK2 (--xchr-model 2), adjusting for technical and biological covariates following the original study^92^. COLOC analyses evaluated both ‘coloc.abf’ with default per-variant priors and ‘coloc.susie’ (coloc R package v.4.2.1)^87,88^. Colocalization analyses were restricted to variants located within ±1 Mb of the lead associated AD variant. Genes with PP4>0.6 marked successful colocalization. Additional details on COLOC analyses are provided in the **Supplemental Methods**. For loci where genetic colocalization did not identify any genes, we nominated candidate causal genes by mapping intronic signals to the gene in which they reside and intragenic signals to the nearest protein-coding gene (**Table-S14**)^39^.

### Bulk brain tissue differential expression analyses

To further corroborate whether prioritized AD candidate genes were relevant to AD risk, we interrogated them in bulk brain RNA-sequencing datasets from three independent cohorts that have been centrally processed by AMP-AD to evaluate differential expression of genes (DEG) across AD cases and controls (cf. data availability statement). The phenotype definitions in these analyses integrated both clinical and neuropathological evidence, defining AD cases as those with cognitive impairment, Braak pathology >3, and CERAD scores >2, while controls were defined as those without cognitive impairment, Braak pathology ≤3, and CERAD scores ≤2. The DEG analyses included eight brain regions spanning temporal cortex (N_AD_=80, N_CN_=68) in Mayo Clinic^93^, frontal pole (N_AD_=146, N_CN_=69), inferior frontal gyrus (N_AD_=132, N_CN_=69), parahippocampal gyrus (N_AD_=155, N_CN_=71), and superior temporal gyrus (N_AD_=156, N_CN_=64) in Mount Sinai Brain Bank^94^, and dorsolateral prefrontal cortex (N_AD_=294, N_CN_=143), anterior cingulate cortex (N_AD_=171, N_CN_=94), and posterior cingulate cortex (N_AD_=151, N_CN_=99) in ROSMAP^95^. All DEG analyses included non-sex-stratified as well as sex-stratified evaluations. We extracted DEG results for all 48 genes we prioritized and matched the DEG contrasts to the XWAS model and sex from which the gene was prioritized. Specifically, genes derived from the rXCI AD XWAS models were evaluated using DEG between all AD cases and controls within each cohort. In contrast, genes prioritized from female-specific or eXCI AD XWAS models were evaluated using female-specific case-control contrasts, while genes prioritized from male-specific AD models were evaluated using male case-control contrasts. We then accounted for multiple testing using false discovery rate (FDR)-adjusted p-values within each brain region followed by a Bonferroni correction across brain regions defined as FDR-adjusted P<0.00625 (0.05/8 tissues).

### Locus and gene prioritization

At all candidate AD loci identified through X-wide significance, pleiotropy, or both, candidate causal genes were nominated through QTL colocalization or gene body proximity (cf. above “Candidate gene nomination”) and subsequently evaluated for evidence of DEG across 8 different brain regions. For each gene, we then composed an evidence level score, ranging 0.5 to 4, to reflect the robustness of the underlying genetic signal and functional genomics support. Per-gene scores were summed following these criteria: (i) genes related to an AD genetic signal that passed X-wide significance received 1 point, (ii) genes related to an AD genetic signal with only one pleiotropic trait colocalization received 0.5 points while those with more than 1 colocalization received 1 point (reflecting reduced concern of chance colocalizations), (iii) genes with one QTL colocalization received 0.5 points while those with more than 1 colocalization received 1 point (reflecting more robust gene support), and (iv) genes that passed at most DEG FDR-P<0.05 received 0.5 points versus those that further passed FDR-P<0.05/8 received 1 point.

While scores were calculated for all candidate genes at all associated loci, we focused reporting and subsequent analyses to the top genes across 20 (out of 26) loci that displayed stronger genetic support, reflected as having AD XWAS P<1e-5 or at least 2 pleiotropic trait colocalizations.

### Brain region-specific enrichment of differentially expressed genes

To determine whether prioritized candidate genes were associated with AD risk in specific brain regions more than expected by chance, we tested whether the magnitude of differential expression of genes (DEG) was enriched among prioritized genes relative to the background distribution of X-chromosome genes within each brain region. For each of the eight brain regions examined in DEG analyses, we calculated the mean absolute differential expression effect size (using T-values) for the prioritized gene set. This observed mean was then compared to a null distribution generated through 1,000 random simulations in which the same number of genes was randomly sampled from all X-chromosome genes tested in the corresponding dataset. The T-value was calculated for each simulation to estimate the expected population mean under the null. A Z-test was subsequently used to compare the observed mean of the prioritized genes to the simulated population mean.

### *RBBP7* Single-cell ATAC-seq validation

For the *RBBP7* locus, we assessed overlap of the AD index variant with regulatory peaks from single-cell ATAC-seq data in matching cell-types from human brain tissues available from Corces *et al*.^96^

### Prior evidence of X-chromosome inactivation escape

Prior evidence for eXCI was assessed for prioritized genes through a literature review of 12 published studies that evaluated XCI escape status across human tissues and cell types. For each gene, reported evidence of eXCI was extracted from these studies and summarized to quantify the number of studies supporting escape. The complete list of referenced studies and their corresponding public identifiers, along with a summary eXCI support across prioritized genes is provided in **Table-S17**.

## Data Availability

All summary statistics will be available in NIAGADS and GWAS Catalogue upon publication.

## Data availability

Data used in the XWAS analyses are available upon application to:

- dbGaP (https://www.ncbi.nlm.nih.gov/gap/)
- NIAGADS (https://www.niagads.org/)
- LONI (https://ida.loni.usc.edu/)
- AMP-AD knowledge portal / Synapse (https://www.synapse.org/)
- Rush (https://www.radc.rush.edu/)
- NACC (https://naccdata.org/)
- UKB (https://www.ukbiobank.ac.uk/)
- FinnGen (https://www.finngen.fi/en)
- MVP (https://www.mvp.va.gov/)

The specific data repository and identifier for ADGC and ADSP data are indicated in **Tables-S1-2** of the supplement. Full, stratified XWAS summary statistics will be available in NIAGADS and GWAS Catalogue upon publication.

Data used in the multi-trait pleiotropy analyses are publicly available:

- Age at menopause (https://www.reprogen.org/data_download.html)
- Age at menarche (https://www.repository.cam.ac.uk/items/8c5f7afb-5fa2-45ea-b52d-4e643bc2a5b7)
- Hormone exposure traits (https://www.ukbiobank.ac.uk/)
- Global lipids genetics consortium (https://csg.sph.umich.edu/willer/public/glgc-lipids2021/results/chrx_summary_stats/)
- Jiang et al. MRI phenotypes (https://zenodo.org/records/12676622)

Data used in the xQTL colocalization analyses are publicly available:

- Wingo et al. DLPFC pQTL and eQTL (https://www.synapse.org/Synapse:syn51150434/wiki/621280)
- Fujita et al. brain single cell eQTL data (https://www.synapse.org/Synapse:syn52335807)
- eQTL Catalogue database (https://www.ebi.ac.uk/eqtl/Data_access/)

Data used in the bulk brain differential expression analyses are available upon application to:

- AD Knowledge Portal (https://www.synapse.org/Synapse:syn26720676)

A table overview of all traits used in the multi-trait pleiotropy analyses, and their public identifiers are indicated in **Table-S5** of the supplement. All QTL resources and their public identifiers are indicated in **Table-S12** of the supplement. A public repository of all locus compare plots, with colocalization (PP4 ≥ 0.6) from the multi-trait pleiotropy and xQTL colocalization analyses, can be accessed at **10.5281/zenodo.18807726**.

## Code availability

Codes for this work are available https://github.com/Belloy-Lab/AD_XWAS_Pleiotropy. We used publicly available software for all analyses in this study. This included PLINK2, BOLT-LMM (v.2.5), Regenie, and GWAMA (v.1.2.6), to perform XWAS, and LDSC (v.2.0.0) to format summary statistics. Variant annotation was performed using VEP release 114. coloc (v.4.2.1) R package was used to perform colocalization. CMplot (v.4.5.1) was used to generate circus plots. locuszoomr (v.0.3.8), locuscomparer (v.1.0.0), and ggplot2 (v.3.5.2) were used to create locus zoom and locus compare plots. Ritchie Lab Visualization was used to generate Phenograms. Brain imaging masks were generated using Mango (v.4.1). Other figures utilized cowplot (v.1.2.0), scales (v.1.4.0), patchwork (v.1.3.2), ggrepel (v.0.9.6), ggtext (v.0.1.2), and ggpubr (v.0.6.1) R packages.

## Acknowledgements

We thank all study participants and their families as well as many involved institutions and their staff. This work was supported by grants from the National Institutes of Aging (R00AG075238, M.E.B.; K01-073584, D.B.A.), Cure Alzheimer’s Fund (M.E.B.), and Alzheimer’s Association (AARG-24-1027303). This work was supported by access to equipment made possible by the Hope Center for Neurological Disorders, the Neurogenomics and Informatics Center (NGI: https://neurogenomics.wustl.edu/) and the Departments of Neurology and Psychiatry at Washington University School of Medicine. The ADSP Phenotype Harmonization Consortium (ADSP-PHC) is funded by NIA (U24 AG074855, U01 AG068057 and R01 AG059716). This research is based on data from the Million Veteran Program, Office of Research and Development, Veterans Health Administration, and was supported by MVP000 as well as award I01BX004192 (MVP015) and I01BX005749 (MVP040). This publication does not represent the views of the Department of Veteran Affairs or the United States Government.

We would like to thank the following resources for allowing access to their data: ADGC, ADSP, UK Biobank, FinnGen, Million Veterans Program, AMP-AD. UK Biobank data were analyzed under Application Number 45420. Detailed acknowledgements for different genetic cohorts and biobanks are provided in the supplementary material.

## Author contributions

N.C. performed data acquisition, designed analyses, performed analyses, and wrote paper. C.Y., Y.Z., T.-C.W., Y.L., M.J., R.Z., and R.S. curated data and performed analyses. Z.J., K.C., M.E.K, B.M., L.D., T.J.H., C.P., and C.C. contributed data and analyses. M.L. and J.M.G were involved in data, funding, and resource acquisition. M.L., R.Z., V.C.M., M.S.P., R.L.H., B.G., A.D., S.A., and D.A. were involved in conceptualization and study design. V.N. and M.D.G. supervised analyses and supervised work. M.E.B. performed data acquisition and analyses, designed analyses, designed study, supervised analyses, supervised work, wrote paper, and obtained funding.

