## Supplemental Document for "An *APOE*\*4-Informed Genomic Atlas of the X Chromosome in Alzheimer’s Disease"

### Supplementary Methods

#### X-chromosome wide association studies

In the current study, we used data from a variety of cohorts and sequencing projects related to AD[1–22]. All available genetic/phenotypic data were jointly harmonized with the purpose of performing phenotype/covariate harmonization. Details are provided below.

##### ADGC & ADSP Phenotypic Data Quality Control and Processing

###### *Cohorts and Phenotype Ascertainment*

Details on phenotype ascertainment are described elsewhere[1–5,7]. Briefly, all individuals with a diagnosis of AD met National Institute of Neurological and Communicative Disorders and Stroke/Alzheimer's Disease and Related Disorders Association (NINCDS-ADRDA) criteria for definite, probable, or possible late-onset AD[6], or met Diagnosis and Statistical Manual of Mental Disorders IV-V (DSMIV-V) criteria[8,9], or had a clinical dementia rating (CDR® Dementia Staging Instrument[10]) > 0.5. Some cohorts verified AD diagnoses through neuropathology, using Braak staging[11], CERAD scoring[21], or National Institute on Aging Reagan (NIA-Reagan) 1997 criteria[12]. Cognitively normal subjects did not have AD according to the above clinical AD criteria, did not have a diagnosis of mild-cognitive impairment (MCI), and had a CDR of 0 and/or Mini-Mental State Examination (MMSE[13]) > 25. In MIRAGE, control status was evaluated through a Modified Telephone Interview of Cognitive Status score ≥ 86 (a telephone version of the MMSE)[14].
Further, the National Alzheimer's Coordinating Center (NACC), Rush University Religious Orders Study/Memory and Aging Project (ROSMAP), and Alzheimer's Disease Neuroimaging Initiative (ADNI), are longitudinal cohorts that provide detailed information regarding clinical status (control, MCI, demented) and presumed disease etiology at repeated examinations. Additionally, deceased subjects are assessed for neuropathology. Where possible, in NACC, a final diagnosis of MCI or possible/probable/definite AD was obtained using NIA Alzheimer's Association (NIA-AA) 2011 criteria[15,16]. In all three cohorts, AD diagnoses were verified by neuropathology as middle or high AD likelihood following NIA-Reagan 1997 criteria (moderate to frequent neuritic plaques and Braak stage III-VI)[12]. In concordance with the category "possible AD dementia with evidence of the AD pathophysiological process" from the NIA-AA 2011 criteria[15], we attributed possible AD diagnoses to subjects who met clinical criteria for non-AD dementia but also met AD neuropathological criteria. In concordance with the NIA-AA 2011/2012 framework[16,17], we also evaluated neuropathology in MCI subjects to verify presumed AD etiology. Controls were not re-evaluated based on neuropathology data. Subjects that reverted from dementia to control status during longitudinal follow-up were excluded. Additional cohort-specific details are listed below.

###### *NACC*

Genotyping waves 1 through 7 from the Alzheimer's Disease Centers (ADC1-7) and a subset of the ADSP projects include subjects ascertained and evaluated by the clinical and neuropathological cores of 32 NIA-funded ADCs. NACC coordinates the collection of these phenotypes, implements diagnoses (cognitively normal, cognitively impaired but not MCI, MCI, demented; and presumed disease etiology), and then provides all data to researchers under the form of the Minimum Data Set (MDS), Uniform Data Set (UDS)[18,19,22], and Neuropathology data set (NP)[20]. The MDS represents an older subset of the NACC data and only contains cross-sectional data, while the more recent UDS provides longitudinal phenotypes and covariates. Since 2015, the UDS was updated to incorporate the NIA-AA 2011 criteria for MCI and AD[16,23]. In the current study, we used the UDS and NP for which data was collected between September 2005 and March 2022, to determine phenotypes for subjects in ADC1-7, ADSP WES/WGS, and ADGC Exome arrays.

Subjects that had a diagnosis of Down syndrome, central nervous system neoplasm, bipolar disorder, schizophrenia, alcohol-induced dementia, or substance-abuse-induced dementia, were excluded. Subjects carrying mutations of dominantly inherited AD or frontotemporal lobar degeneration (FTLD) were also excluded. Subjects with a final diagnosis of MCI or dementia, for which the etiology was unknown, not due to AD, or only secondary due to AD (and without AD neuropathological information), were excluded. Subjects with a final diagnosis of "cognitively impaired but not MCI", but having no other neurological disorder, were kept as controls, considering that this more consistently matched control criteria in many of the other cohorts considered in this study.

##### *ROSMAP*

In ROSMAP, subjects were diagnosed at each visit: as possible/probable AD according to NINCDS-ADRDA criteria[6]; as MCI when judged to have cognitive impairment but not meeting dementia criteria according to the clinician; or as control when there was no cognitive impairment or the subject did not meet dementia criteria[24,25]. At time of death, a final clinical diagnosis was made by an expert neurologist, followed by a case conference consensus review (blinded to postmortem data)[26].

##### *ADNI*

In ADNI, subjects were diagnosed at regular visits: as possible/probable AD according to NINCDS-ADRDA criteria[6]; as MCI according to Petersen/Winblad criteria; or as control when not demented, not MCI, CDR = 0, and MMSE > 28. Neuropathology assessments followed the NACC NP framework.

##### *Phenotype Harmonization*

The available sample contained many subjects that were genotyped multiple times across different studies. This largely reflected efforts from the ADGC, ADSP, and AMP-AD, to perform next-generation sequencing (NGS) on existing cohort samples for the purpose of rare variant discovery and AD gene prioritization. In other instances, participants were recruited in different studies at different times. Therefore, to handle potential duplicate discordance and phenotype heterogeneity, we implemented a cross-sample phenotype harmonization procedure aiming to standardize pathology-verified diagnoses where possible, share unique missing information across all duplicate entries of a given subject, resolve longitudinal changes in diagnosis, and flag subjects with unresolvable duplicate discordance for exclusion. Duplicate samples were identified by determining genetic cryptic relatedness (cf. below), but for sample cross-referencing did not include known identical twins in LOAD and ROSMAP samples. First, duplicate samples were flagged as discordant if their age-at-death information differed by more than 2 years or if pathology measures (Braak or neuritic plaque density) differed. Across all cohorts, where possible, AD diagnoses were verified by neuropathology as middle or high AD likelihood following NIA-Reagan 1997 criteria (moderate to frequent neuritic plaques and Braak stage III-VI)[12]. Additionally, when only either neuritic plaque or Braak information was available and in line with NIA-Reagan 1997 middle or high AD likelihood criteria, and/or the cohort/project demographics provided a diagnosis of definite AD, the subject was considered to have pathology-verified AD status. Cognitively normal (CN) subjects with evidence of AD pathology were kept as CN. Further, if at least one entry across duplicate samples indicated a diagnosis of Down syndrome, central nervous system neoplasm, bipolar disorder, schizophrenia, alcohol-induced dementia, substance-abuse-induced dementia, neurological (not including Parkinson's disease), or systemic disease despite being cognitively normal, or carrying mutations of dominantly inherited AD or frontotemporal lobar degeneration (FTLD), then all duplicate samples were marked as such and flagged for exclusion. Extending on the above, all genetic samples were checked for the presence of known pathogenic mutations on *APP, PSEN1, PSEN2,* and *MAPT*, whereby carriers and their duplicate samples were flagged for exclusion.

Then, duplicate samples with differing age entries (i.e. longitudinal changes) were evaluated. Reversions from AD or dementia to MCI status, or from MCI to cognitively normal (CN) status, were permitted, but reversions from AD or non-AD dementia to CN status were flagged for exclusion. "Reversions" from AD to non-AD dementia status were permitted, unless pathology (cf. above) indicated the presence of AD pathology, thereby marking the subject as AD. Vice versa, "conversions" from non-AD dementia to AD status were permitted, unless pathology (cf. above) indicated no presence of AD pathology, thereby marking the subject as non-AD dementia. All other types of conversions were directly permitted. Then, duplicate samples for which the diagnoses at the oldest shared age entries differed, or for which diagnoses differed, but age was consistent (i.e. apparent cross-sectional discordances), were evaluated. Discordances between AD and non-AD dementia status were resolved based on pathology (cf. above) or flagged as discordant if no pathology data was available. Discordances between CN and AD status, or CN and non-AD dementia status, were resolved as respectively AD or non-AD dementia when those dementia diagnoses corresponded to a unique age-at-onset (of symptoms) without other available age information (i.e. indicating that a conversion likely occurred after the subject was lost to follow-up in the cohort that last observed a CN status), or, were flagged as discordant if duplicate entries shared the same age-at-examination and age-at-last-exam. Discordances between CN and MCI status, or MCI and AD status, or MCI and non-AD dementia status, were resolved as respectively MCI, AD, or non-AD dementia (i.e. keeping the most severe diagnosis).

Finally, once all clinical diagnostic and pathological data were unified across duplicate entries, pathological criteria were applied once more to obtain the final diagnoses. Where possible, AD diagnoses were verified by neuropathology as middle or high AD likelihood following NIA-Reagan 1997 criteria (moderate to frequent neuritic plaques and Braak stage III-VI)[12]. In concordance with the category "possible AD dementia with evidence of the AD pathophysiological process" from the NIA-AA 2011 criteria[15], we attributed possible AD diagnoses to subjects who met clinical criteria for non-AD dementia but also met AD neuropathological criteria. In concordance with the NIA-AA 2011/2012 framework[16,17], we also evaluated neuropathology in MCI subjects to verify presumed AD etiology and considered subjects as cases if AD pathology, following NIA-Reagan 1997 criteria (cf. above), was present (i.e. marking high likelihood of AD etiology). Controls were not re-evaluated based on neuropathology data.

Beyond cross-referencing clinical diagnostic and pathological data across subjects, other covariates were considered for cross-referencing or sharing in case of missingness across duplicate entries. These included age-at-onset of cognitive symptoms, age-at-examination providing clinical diagnosis, at-at-last exam, age-at-death, sex, race, ethnicity, *APOE* genotype provided from demographics, *APOE* genotype provided from whole-genome sequencing, and *APOE* genotype provided from whole-exome sequencing. Duplicate entries with discordant sex or race information were flagged for exclusion.

#### ADGC and ADSP Genetic Data Quality Control and Processing

##### *Ascertainment of Genetic Data*

Genotypes were available from high-density single-nucleotide polymorphism (SNP) genotyping microarrays (Illumina or Affymetrix) for ADGC or whole genome sequencing (WGS) for ADSP (**Tables-S1-2**). Genotype samples had their genetic variants lifted to hg38 using liftOver if not released in hg38 and annotated using dbSNP153 variant identifiers[27].

##### *ADGC Autosomal Quality Control*

Autosomal variants were extracted from the SNP array data and further processed in several stages. In each cohort/platform/array, variants were excluded based on genotyping rate (<95%), MAF<1%, and Hardy-Weinberg equilibrium in controls ($p<10^{-6}$) using PLINK v1.9[28]. As in our prior work[29], information derived from the gnomAD v.3.1 database[30] was used to filter out SNPs that met one of the following exclusion criteria: (i) located in a low complexity region, (ii) located within common structural variants (MAF > 1%), (iii) multiallelic SNPs with MAF > 1% for at least two alternate alleles, (iv) located within a common insertion/deletion, (v) having any flag different than PASS in gnomAD, (vi) having potential probe polymorphisms, and (vii) more than 10% MAF difference with gnomAD frequency in non-Finnish Europeans. The remaining SNPs were checked for consistency with the TOPMed panel, flipping of palindromic SNPs, and were imputed on the TOPMed Imputation server[31,32], which uses Minimac 4 for imputation. The following parameters were selected: reference panel TOPMed-r2 (2022), phasing with Eagle v2.4, r-square imputation score cut off 0.3.

##### *ADSP Autosomal Quality Control*

The ADSP WGS data (NG00067.v5) were joint called by the ADSP following the SNP/Indel Variant Calling Pipeline and data management tool used for the analysis of genome and exome sequencing for the Alzheimer's Disease Sequencing Project (VCPA)[33]. The current analyses of ADSP WGS were restricted to bi-allelic variants, to which we applied the Variant Quality Score Recalibration (VSQR) quality control filter ("PASS" variants; GATK v4.1)[34]. Variants with a genotyping rate less than 80%, deviating from Hardy Weinberg Equilibrium (HWE) in the full sample or in controls ($p<10^{-6}$), and a minor allele count less than 10, were excluded. Consistent with the methodology detailed in Belloy et al. 2022[35], we then applied several filters to remove artifactual variants: (i) variants that represented sequencing center or platform artifacts as identified by Fisher exact testing in controls ($p<10^{-5}$), (ii) variants reported in gnomAD v3.1[36] to have a "non-PASS", falling in a low complexity region, or showing more than 10% allele frequency deviation between our European ancestry control participants in ADSP and non-Finnish European participants in gnomAD, and (iii) duplicate discordance variants that show discrepancies across several 100 technical duplicates present in ADSP.

##### *Genetic Relationship Determination using King*

Across all cohorts, the relatedness of subjects (after QC indicated above) was evaluated through identity-by-descent (IBD) analysis (using directly genotyped non-palindromic SNPs shared across all genetic datasets with a call rate > 95% & minor allele frequency (MAF)>1%)[37]. This outcome was used for duplicate tracking across samples, which in turn was used to enable phenotype harmonization (cf. above).

##### *Ancestry Determination*

Individual ancestries were determined using SNPweights v.2.1 with populations from the 1000 Genomes Consortium as a reference[38,39]. By applying an ancestry percentage cut-off ≥ 75%, the samples were stratified into the five super populations, South-Asians (SAS), East-Asians (EAS), Amerindians (AMR), Africans (AFR) and Europeans (EUR) (Belloy et al., 2024[40] Figure-S1). When multiple samples were available for a single unique individual, the ancestry was inferred from the sample with the highest genetic coverage.

##### *Restriction to European ancestry for XWAS*

XWAS were focused on the European ancestry subsample. Two main reasons explain the more extreme population structure on the X-chromosome compared to autosomes: (i) the X-chromosome has a smaller effective population size and thus the rate of genetic drift of X-linked loci is amplified, (ii) local adaptation will lead to higher levels of differentiation between geographically isolated populations[41]. As such, to better control for population structure, we restricted our analyses to European ancestry participants.

##### *Relationship Determination and Principal Component Analysis using GENESIS*

For ADGC and ADSP data respectively, the relatedness of subjects and principal components capturing population substructure were determined using IBD and principal component analyses (PCA) as implemented through the R package GENESIS (R v3.6.0)[42]. Specifically, this approach first uses an R-implementation of KING-robust to determine kinship coefficients that take into account ancestry divergence. The derived pairwise kinship coefficients are then used to perform a PCA in related samples

(PC-AiR) providing accurate ancestry inference not confounded by family structure. The latter output is then used to estimate kinship coefficients using PC-Relate, which accounts for population structure (ancestry) among sample individuals through the use of ancestry representative principal components (PCs) to provide accurate relatedness estimates due only to recent family (pedigree) structure. For each respective data set, these analyses were performed on pruned SNPs (R2 < 0.5, call rate > 95%, MAF > 1%, and excluding palindromic SNPs) in non-Hispanic White European ancestry individuals.

##### *ADGC X chromosome Quality Control and TOPMed Imputation*

The X chromosome variants underwent a similar harmonization pipeline as the autosomes. We excluded multi-allelic SNPs, SNPs within structural variations, and potential probe polymorphism SNPs. Additionally, our analysis excluded the pseudoautosomal regions of the X chromosome and used only the European ancestry participants as derived above. Several steps were performed to avoid spurious findings: (i) variants with less than 95% genotyping rate and (ii) individuals with more than 5% genotype missingness were excluded. (iii) Reported sex was checked using PLINK1.9 --check-sex flag[28], with 0.4 max value for females and 0.94 min value for men, and all individuals with a discordant sex label were excluded. (iv) Heterozygous SNPs in males were set as missing in males, while (v) SNPs with differential missingness between AD cases and controls were removed (p<10-5 per cohort/platform/array). (vi) HWE was tested in female controls and SNPs with p<10-5 were removed (per cohort/platform/array). (vii) Any monomorphic SNPs that remained were removed. (viii) Differential missingness and differential MAF between males and females were both tested and SNPs with p<10-5, for either one of the tests, were excluded (per cohort/platform/array). Finally, as for the autosomes and based on gnomAD v3.1[36] information, we filtered variants (ix) located in a low complexity region, (x) located within common structural variants (MAF > 1%), (xi) multiallelic SNPs with MAF>1% for at least two alternate alleles, (xii) located within a common insertion/deletion, (xiii) having any flag different than PASS in gnomAD v.3.1, (xiv) having potential probe polymorphisms (xv) more than 10% MAF difference with gnomAD frequency in non-Finnish Europeans. The remaining SNPs were checked for consistency with the TOPMed panel, flipping of palindromic SNPs, and were imputed on the TOPMed Imputation server[31,32], which uses Minimac 4 for imputation. The following parameters were selected: reference panel TOPMed-r2 (2022), phasing with Eagle v2.4, r-square imputation score cut off 0.3.

##### *ADSP X chromosome Quality Control*

The X chromosome variants underwent a similar harmonization pipeline as the autosomes. Our analysis excluded the pseudoautosomal regions of the X chromosome and used only the European ancestry participants. The ADSP WGS data for X chromosome (NG00067.v5) were joint called by the ADSP following the SNP/Indel Variant Calling Pipeline and data management tool used for the analysis of genome and exome sequencing for the Alzheimer's Disease Sequencing Project (VCPA)[33]. The current analyses of ADSP WGS were restricted to bi-allelic variants, to which we applied the Variant Quality Score Recalibration (VSQR) quality control filter ("PASS" variants; GATK v4.1)[34]. Variants with a genotyping rate of less than 80% and a minor allele count of less than 2 were excluded. Consistent with the methodology detailed in Belloy et al. 2022[35], we then applied several filters to remove artifactual variants: (i) variants that represent sequencing center or platform artifacts as identified by Fisher exact testing in controls (p<10-5), (ii) variants reported in gnomAD v3.1 to have a "non-PASS", falling in a low complexity region, or showing more than 10% allele frequency deviation between our European ancestry control participants in ADSP and non-Finnish European participants in gnomAD, and (iii) duplicate discordance variants that show discrepancies across several 100 technical duplicates present in ADSP.

Several additional steps were performed to avoid spurious findings: (i) Heterozygous SNPs in males were set as missing in males, (ii) variants with a genotyping rate less than 80% in controls, cases, men, or women were excluded, (iii) variants with differential missingness between AD cases and controls were removed (p<10-10 for the full sample). (iv) HWE was tested in female and male controls using the Plink --hardy command that allows joint sex evaluation and variants with p<10-5 were removed (for the full sample). (v) Differential missingness between males and females was tested and SNPs with p<10-20 were excluded (for the full sample).

#### ADGC & ADSP Statistical Analyses

##### *Case-control XWAS*

All association analyses with AD risk were adjusted for sex (in non-stratified XWAS), array type, the first 5 genetic principal components (PC-AiRs), *APOE**4 dosage (0/1/2), and *APOE**2 dosage (0/1/2). Age adjustment in case-control analyses was not performed, given that the current AD genetic samples often showed younger ages for cases than controls due to the use of age-at-onset information (**Table-S3**), which violates the assumption for age adjustment (which is that older age is associated with increased AD incidence). In prior work, we showed that age adjustment in such scenarios leads to significantly decreased power for genetic association analyses[29]. Adjustment for *APOE* genotypes is relevant given the established interactions with sex[43], which may notably be relevant to the X chromosome and could lead to increased model noise if not accounted for. Additionally, the case-control clinical cohorts are enriched for *APOE**4 cases compared to population-based studies[43], which may further exacerbate any potential confounding effects.

Cohorts from ADGC were pooled into a mega-analysis. LMM-BOLT was used in both ADGC and ADSP[44], using autosomal data to derive genetic relationship matrices to allow the inclusion of related subjects. Resultant betas were converted to traditional odds ratios using the transformation approach as detailed in the LMM-BOLT manual. Across ADGC and ADSP, subjects were unrelated down to 1st degree.

##### *Age information*

For cases that only had age-at-death (AAD) available, the final ages used for regression analysis were subtracted by 10 years to approximate age-at-onset (AAO). This reflects expected mean delays between AAO and AAD for AD patients[45], and is consistent with the derived age covariate for AD cohorts provided by the Alzheimer's Disease Genetics Consortium (ADGC) on NIAGADS[46]. In cohorts that provide conversion information but not AAO, age-at-examination (AAE) was used and followed a prioritization of age-at-MCI-diagnosis > age-at-dementia-diagnosis (incident) > age-at-dementia diagnosis (prevalent). This was done to most closely approximate AAO. For the remaining control samples, age-at-last-examination (AAL) was used. After implementing these criteria, samples were filtered to have a minimal age of 60 years. Some samples were censored at ages 90+, for which we assumed the age was 90 (since there was no way to estimate the actual age).

#### UKB Phenotype Ascertainment

Detailed descriptions of all the variables and field provided by UKB are provided elsewhere[47].

In the first round of phenotype ascertainment, we derived health-registry-confirmed AD status and related age information for the individuals directly. Subjects were assumed to be controls if they had no other diagnosis inferred from health registry information relevant to dementia status. We specifically considered the following data fields and entries: *Diagnoses_main_ICD10* [G300,G301,G308,G309,F000,F001,F002,F009], *Diagnoses_secondary_ICD10* [G300,G301,G308, G309,F000,F001,F002,F009], *Date_of_first_in_patient_diagnosis_main_ICD10* [if date provided], *Date_of_first_in_patient_diagnosis_ICD10* [if date provided], *Source_of_alzheimers_disease_ report* [0,1,11,12,2,21,22 = self-report, hospital admission, death record], *Date_of_alzheimers_disease_ report* [if date provided], *Source_of_all_cause_dementia_report* [0,1,11,12,2,21,22 = self-report, hospital admission, death record], *Source_of_frontotemporal_dementia_report* [any entry], *Source_of_vascular_ dementia_report* [any entry], and *Date_of_all_cause_dementia_report* [if data provided]. The above fields were used to determine dementia status, allowing us to differentiate between late-onset AD individuals (LOAD), early-onset AD (EOAD), vascular dementia, frontotemporal dementia, and other all-cause dementia participants. For the health-registry AD phenotype, cases were restricted to all LOAD individuals. The above fields were further used to determine the earliest available age at which a dementia occurrence or report was made. Age information for controls was available from the variables: *Age_when_attended_assessment_centre* [oldest age entry retrieved] and *Age_at_death*.

We then identified the proxy ADD case and control status and related age by accessing the following fields and entries: *Illnesses_of_father*, *Illnesses_of_mother*, *Illnesses_of_sibblings*, *Fathers_age*, *Fathers_age_at_death*, *Mothers_age*, and *Mothers_age_at_death* (where it should be noted that age and sex info was not available for siblings). The youngest reported age was used for proxy ADD cases, while the oldest reported age was used for proxy controls. Proxy status was ignored if subjects were adopted. Both health-registry-confirmed AD status and proxy ADD status were then combined into a single phenotype (cf. Belloy et al., 2024[40] eTables6-7). Notably, all subjects were ages >60y in at least the subject or one parent.

#### UKB Genetic Data Quality Control and Processing

A Detailed description of all the UKB genetic data and processing is provided elsewhere46. Specifically, we accessed SNP array data imputed to the Haplotype Reference Consortium (HRC) and UK10K haplotype resource. We further filtered to subjects with consent, passing sex check QC, no heterozygosity outliers, having age information available, and belonging to a white ethnic background (field Ethnic_background [1001,1002,1003]). We then identified a homogenous ancestry cluster within this group using “aberrant” on the first 20 genetic PCs, as in Schwartzentruber et al. 2021[48].

#### UKB Statistical Analyses

All association analyses with the AD phenotype were adjusted for sex (in non-stratified XWAS), array type, assessment center, the first 20 genetic principal components provided by UKB, and APOE*4/2 dosage. LMM-BOLT was used (as was done for ADGC and ADSP)[44], using autosomal data to derive genetic relationship matrices to allow the inclusion of related subjects. Resultant betas were converted to traditional odds ratios using the transformation approach as detailed in the LMM-BOLT manual. Additionally, since the UKB XWAS leveraged the proxy phenotype, an additional correction factor was needed to rescale beta coefficients onto a regular case-control scale. This correction is detailed in Belloy et al., 2024[40] eTables6-7.

#### Finngen

The FinnGen study is a large-scale genomics initiative that has analyzed over 500,000 Finnish biobank samples and correlated genetic variation with health data to understand disease mechanisms and predispositions. The project is a collaboration between research organisations and biobanks within Finland and international industry partners.

Ascertainment of FinnGen phenotype and genotype data is described in detail elsewhere[49]. Summary statistics were available from version 10 (v10) of the publicly released set of genetic summary statistics made available by FinnGen here: https://www.finngen.fi/en/access_results. Documentation on Genetic data processing and statistical analyses is provided here: https://finngen.gitbook.io/documentation. FinnGen made us of Regenie to include related individuals in the genetic association analyses[50].

Information about phenotypes and endpoints is provided here: https://www.finngen.fi/en/researchers/clinical-endpoints, https://r10.risteys.finngen.fi/. We specifically leveraged the "Alzheimer's disease, wide definition" phenotype.

#### MVP Phenotype Ascertainment

Generation of phenotypes for MVP mirrors methods as described in Sherva et al. 2023[51], but was updated based on the more recent MVP 2022_1 data release. Briefly, MVP phenotype data were generated from VA electronic medical records. Alzheimer's disease and related dementia (ADRD) cases were identified on the basis of International Classification of Disease (ICD) 9 and 10 codes. Information on proxy cases (parental dementia) was obtained from the MVP Baseline Survey[52].

#### MVP Genetic Data Quality Control and Processing

Generation and quality control of the MVP genetic data is described in detail elsewhere[53]. Briefly, the genetic data were genotyped using the MVP 1.0 custom Axiom array[12], phasing was performed by SHAPEIT4 v 4.1.3, and imputation was performed with MINIMAC4 based on the TopMed imputation reference panel[31]. Variants were then filtered to imputation scores > 0.4 and allele frequencies >= 0.1% in the full dataset. The subset of European-ancestry MVP participants, as determined by the genetically informed Harmonized Ancestry and Race/Ethnicity (HARE) method[54], was then extracted for XWAS and variants were subsequently filtered to allele frequencies >= 0.05%.

#### MVP Statistical Analyses

As in Sherva et al. 2023[51], our ADRD case-control analysis was independent of the cohort used for the proxy analysis. The MVP case-control XWAS (MVP-1) included ICD-identified cases with onset after age 60. The controls were all over age 65 without any dementia or mild cognitive impairment ICD codes and without a history of AD medication usage. The MVP proxy XWAS (MVP-2) was performed separately using a set of controls without any report of parental dementia. Only survey data for Veteran participants over age 45 at last visit were included in proxy analyses (age for parents was not available). The proxy analyses were focused on maternal phenotypes only. This represents the majority of proxy samples since paternal phenotypes cannot be used for men and there was a relative paucity of women. In both the MVP-1 and MVP-2 cohorts, Plink was used to conduct case-control logistic regression analyses on unrelated subjects. For the proxy XWAS, a correction factor was needed to rescale beta coefficients onto a regular case-control scale. This correction is detailed in Belloy et al., 2024[40] eTables6-7.

### Multi-trait pleiotropy

For each AD–trait pair, variants were selected within ±1 Mb of representative AD XWAS variants as described in the **Methods**. Summary statistics from AD and external trait datasets were harmonized using genomic position and allele matching. Variants not present in both datasets were excluded, and effect alleles were aligned between studies. Variants displaying allele frequency differences greater than 10% between datasets were removed. For pleiotropy analyses, the index variant was defined as the lead AD XWAS variant at an independent locus that demonstrated nominal association in both AD and the corresponding external trait dataset ($P < 1 \times 10^{-3}$ in each study). When multiple variants satisfied this criterion within a region, LD clumping was used to define representative variants.

Genetic colocalization between AD and external traits was evaluated using the coloc R package[55,56] (v4.2.1) through the functions coloc.abf and coloc.susie. The coloc.abf implementation used the p-value–based approach without effect size or standard error inputs to avoid potential instability for variants with allele frequencies near 0.5. To evaluate loci potentially containing multiple independent signals, coloc.susie was applied using the SuSiE (Sum of Single Effects) Bayesian regression framework. The runsusie function was executed using effect size, variance, and minor allele frequency inputs, together with a linkage disequilibrium matrix derived from stage 1 ADGC XWAS data to identify credible sets of independent causal variants. If SuSiE failed to converge after seven days of runtime, the respective analysis was terminated and no results were reported. Because most external trait summary statistics were reported in the GRCh37 genome build, AD XWAS summary statistics were lifted from GRCh38 to GRCh37 prior to pleiotropy analyses. Following colocalization testing, resulting loci were converted back to GRCh38 coordinates for downstream analyses and reporting.

#### β-Amyloid PET XWAS

Amyloid PET data with matching genetic samples for 4,172 subjects aged 50 and older were processed from the A4, ADNI, Berkeley, and HABS cohorts. Both amyloid positivity status and scalar amyloid values for an AD-relevant brain composite were derived and used to generate novel amyloid XWAS. Amyloid positivity status was defined using the Gaussian mixture model (GMM) threshold of 50%. We performed sex-stratified and sex-interaction XWAS within each cohort using Plink1.9, covarying for age and the first five genetic ancestry PCs. The resulting summary statistics from the sex-stratified and interactional models were meta-analyzed separately using GWAMA[57]. Variants present in at least 80% of the cohort population were included for downstream analyses.

#### xQTL Colocalization

Colocalization between AD XWAS signals and molecular QTL datasets (xQTL) was performed using the same analytical framework as the multi-trait pleiotropy analyses explained above. Variants within ±1 Mb of lead XWAS variants were extracted and harmonized between AD and QTL datasets using genomic position alignment and allele matching, with effect allele frequencies required to agree within 10%. If QTL studies used smaller locus windows, analyses were inherently restricted to variants overlapping between AD XWAS and QTL datasets. All xQTL colocalization analyses were performed in the GRCh38 genome build. For QTL datasets originally reported in GRCh37, variant coordinates were lifted to GRCh38 prior to analysis.

### Acknowledgements

Data for this study were prepared, archived, and distributed by the National Institute on Aging Alzheimer's Disease Data Storage Site (NIAGADS) at the University of Pennsylvania (U24-AG041689), funded by the National Institute on Aging. The contents of this article do not represent the views of the National Institutes of Health, the U.S. Department of Veterans Affairs, or the United States Government.

#### Acknowledgements for the use of ADSP data

The Alzheimer's Disease Sequencing Project (ADSP) is comprised of two Alzheimer's Disease (AD) genetics consortia and three National Human Genome Research Institute (NHGRI) funded Large Scale Sequencing and Analysis Centers (LSAC). The two AD genetics consortia are the Alzheimer's Disease Genetics Consortium (ADGC) funded by NIA (U01 AG032984), and the Cohorts for Heart and Aging Research in Genomic Epidemiology (CHARGE) funded by NIA (R01 AG033193), the National Heart, Lung, and Blood Institute (NHLBI), other National Institute of Health (NIH) institutes and other foreign governmental and non-governmental organizations. The Discovery Phase analysis of sequence data is supported through UF1AG047133 (to Drs. Schellenberg, Farrer, Pericak-Vance, Mayeux, and Haines); U01AG049505 to Dr. Seshadri; U01AG049506 to Dr. Boerwinkle; U01AG049507 to Dr. Wijsman; and U01AG049508 to Dr. Goate and the Discovery Extension Phase analysis is supported through U01AG052411 to Dr. Goate, U01AG052410 to Dr. Pericak-Vance and U01 AG052409 to Drs. Seshadri and Fornage.
Sequencing for the Follow Up Study (FUS) is supported through U01AG057659 (to Drs. PericakVance, Mayeux, and Vardarajan) and U01AG062943 (to Drs. Pericak-Vance and Mayeux). Data generation and harmonization in the Follow-up Phase is supported by U54AG052427 (to Drs. Schellenberg and Wang). The FUS Phase analysis of sequence data is supported through U01AG058589 (to Drs. Destefano, Boerwinkle, De Jager, Fornage, Seshadri, and Wijsman), U01AG058654 (to Drs. Haines, Bush, Farrer, Martin, and Pericak-Vance), U01AG058635 (to Dr. Goate), RF1AG058066 (to Drs. Haines, Pericak-Vance, and Scott), RF1AG057519 (to Drs. Farrer and Jun), R01AG048927 (to Dr. Farrer), and RF1AG054074 (to Drs. Pericak-Vance and Beecham).
The ADGC cohorts include: Adult Changes in Thought (ACT) (UO1 AG006781, UO1 HG004610, UO1 HG006375, U01 HG008657), the Alzheimer's Disease Centers (ADC) ( P30 AG019610, P30 AG013846, P50 AG008702, P50 AG025688, P50 AG047266, P30 AG010133, P50 AG005146, P50 AG005134, P50 AG016574, P50 AG005138, P30 AG008051, P30 AG013854, P30 AG008017, P30 AG010161, P50 AG047366, P30 AG010129, P50 AG016573, P50 AG016570, P50 AG005131, P50 AG023501, P30 AG035982, P30 AG028383, P30 AG010124, P50 AG005133, P50 AG005142, P30 AG012300, P50 AG005136, P50 AG033514, P50 AG005681, and P50 AG047270), the Chicago Health and Aging Project (CHAP) (R01 AG11101, RC4 AG039085, K23 AG030944), Indianapolis Ibadan (R01 AG009956, P30 AG010133), the Memory and Aging Project (MAP) ( R01 AG17917), Mayo Clinic (MAYO) (R01 AG032990, U01 AG046139, R01 NS080820, RF1 AG051504, P50 AG016574), Mayo Parkinson's Disease controls (NS039764, NS071674, 5RC2HG005605), University of Miami (R01 AG027944, R01 AG028786, R01 AG019085, IIRG09133827, A2011048), the Multi-Institutional Research in Alzheimer's Genetic Epidemiology Study (MIRAGE) (R01 AG09029, R01 AG025259), the National Cell Repository for Alzheimer's Disease (NCRAD) (U24 AG21886), the National Institute on Aging Late Onset Alzheimer's Disease Family Study (NIA- LOAD) (R01 AG041797), the Religious Orders Study (ROS) (P30 AG10161, R01

AG15819), the Texas Alzheimer's Research and Care Consortium (TARCC) (funded by the Darrell K Royal Texas Alzheimer's Initiative), Vanderbilt University/Case Western Reserve University (VAN/CWRU) (R01 AG019757, R01 AG021547, R01 AG027944, R01 AG028786, P01 NS026630, and Alzheimer's Association), the Washington Heights-Inwood Columbia Aging Project (WHICAP) (RF1 AG054023), the University of Washington Families (VA Research Merit Grant, NIA: P50AG005136, R01AG041797, NINDS: R01NS069719), the Columbia University HispanicEstudio Familiar de Influencia Genetica de Alzheimer (EFIGA) (RF1 AG015473), the University of Toronto (UT) (funded by Wellcome Trust, Medical Research Council, Canadian Institutes of Health Research), and Genetic Differences (GD) (R01 AG007584). The CHARGE cohorts are supported in part by National Heart, Lung, and Blood Institute (NHLBI) infrastructure grant HL105756 (Psaty), RC2HL102419 (Boerwinkle) and the neurology working group is supported by the National Institute on Aging (NIA) R01 grant AG033193.

The CHARGE cohorts participating in the ADSP include the following: Austrian Stroke Prevention Study (ASPS), ASPS-Family study, and the Prospective Dementia Registry-Austria (ASPS/PRODEM-Aus), the Atherosclerosis Risk in Communities (ARIC) Study, the Cardiovascular Health Study (CHS), the Erasmus Rucphen Family Study (ERF), the Framingham Heart Study (FHS), and the Rotterdam Study (RS). ASPS is funded by the Austrian Science Fond (FWF) grant number P20545-P05 and P13180 and the Medical University of Graz. The ASPS-Fam is funded by the Austrian Science Fund (FWF) project I904),the EU Joint Programme - Neurodegenerative Disease Research (JPND) in frame of the BRIDGET project (Austria, Ministry of Science) and the Medical University of Graz and the Steiermärkische Krankenanstalten Gesellschaft. PRODEM-Austria is supported by the Austrian Research Promotion agency (FFG) (Project No. 827462) and by the Austrian National Bank (Anniversary Fund, project 15435. ARIC research is carried out as a collaborative study supported by NHLBI contracts (HHSN268201100005C, HHSN268201100006C, HHSN268201100007C, HHSN268201100008C, HHSN268201100009C, HHSN268201100010C, HHSN268201100011C, and HHSN268201100012C). Neurocognitive data in ARIC is collected by U01 2U01HL096812, 2U01HL096814, 2U01HL096899, 2U01HL096902, 2U01HL096917 from the NIH (NHLBI, NINDS, NIA and NIDCD), and with previous brain MRI examinations funded by R01-HL70825 from the NHLBI. CHS research was supported by contracts HHSN268201200036C, HHSN268200800007C, N01HC55222, N01HC85079, N01HC85080, N01HC85081, N01HC85082, N01HC85083, N01HC85086, and grants U01HL080295 and U01HL130114 from the NHLBI with additional contribution from the National Institute of Neurological Disorders and Stroke (NINDS). Additional support was provided by R01AG023629, R01AG15928, and R01AG20098 from the NIA. FHS research is supported by NHLBI contracts N01-HC-25195 and HHSN268201500001I. This study was also supported by additional grants from the NIA (R01s AG054076, AG049607 and AG033040 and NINDS (R01 NS017950). The ERF study as a part of EUROSPAN (European Special Populations Research Network) was supported by European Commission FP6 STRP grant number 018947 (LSHG-CT-2006-01947) and also received funding from the European Community's Seventh Framework Programme (FP7/2007-2013)/grant agreement HEALTH-F4- 2007-201413 by the European Commission under the programme "Quality of Life and Management of the Living Resources" of 5th Framework Programme (no. QLG2-CT-2002- 01254). High-throughput analysis of the ERF data was supported by a joint grant from the Netherlands Organization for Scientific Research and the Russian Foundation for Basic Research (NWO-RFBR 047.017.043). The Rotterdam Study is funded by Erasmus Medical Center and Erasmus University, Rotterdam, the Netherlands Organization for Health Research and Development (ZonMw), the Research Institute for Diseases in the Elderly (RIDE), the Ministry of

Education, Culture and Science, the Ministry for Health, Welfare and Sports, the European Commission (DG XII), and the municipality of Rotterdam. Genetic data sets are also supported by the Netherlands Organization of Scientific Research NWO Investments (175.010.2005.011, 911-03-012), the Genetic Laboratory of the Department of Internal Medicine, Erasmus MC, the Research Institute for Diseases in the Elderly (014-93-015; RIDE2), and the Netherlands Genomics Initiative (NGI)/Netherlands Organization for Scientific Research (NWO) Netherlands Consortium for Healthy Aging (NCHA), project 050-060-810. All studies are grateful to their participants, faculty and staff. The content of these manuscripts is solely the responsibility of the authors and does not necessarily represent the official views of the National Institutes of Health or the U.S. Department of Health and Human Services.

The FUS cohorts include: the Alzheimer's Disease Centers (ADC) ( P30 AG019610, P30 AG013846, P50 AG008702, P50 AG025688, P50 AG047266, P30 AG010133, P50 AG005146, P50 AG005134, P50 AG016574, P50 AG005138, P30 AG008051, P30 AG013854, P30 AG008017, P30 AG010161, P50 AG047366, P30 AG010129, P50 AG016573, P50 AG016570, P50 AG005131, P50 AG023501, P30 AG035982, P30 AG028383, P30 AG010124, P50 AG005133, P50 AG005142, P30 AG012300, P50 AG005136, P50 AG033514, P50 AG005681, and P50 AG047270), Alzheimer's Disease Neuroimaging Initiative (ADNI) (U19AG024904), Amish Protective Variant Study (RF1AG058066), Cache County Study (R01AG11380, R01AG031272, R01AG21136, RF1AG054052), Case Western Reserve University Brain Bank (CWRUBB) (P50AG008012), Case Western Reserve University Rapid Decline (CWRURD) (RF1AG058267, NU38CK000480), CubanAmerican Alzheimer's Disease Initiative (CuAADI) (3U01AG052410), Estudio Familiar de Influencia Genetica en Alzheimer (EFIGA) (5R37AG015473, RF1AG015473, R56AG051876), Genetic and Environmental Risk Factors for Alzheimer Disease Among African Americans Study (GenerAAtions) (2R01AG09029, R01AG025259, 2R01AG048927), Gwangju Alzheimer and Related Dementias Study (GARD) (U01AG062602), Hussman Institute for Human Genomics Brain Bank (HIHGBB) (R01AG027944, Alzheimer's Association "Identification of Rare Variants in Alzheimer Disease"), Ibadan Study of Aging (IBADAN) (5R01AG009956), Mexican Health and Aging Study (MHAS) (R01AG018016), Multi-Institutional Research in Alzheimer's Genetic Epidemiology (MIRAGE) (2R01AG09029, R01AG025259, 2R01AG048927), Northern Manhattan Study (NOMAS) (R01NS29993), Peru Alzheimer's Disease Initiative (PeADI) (RF1AG054074), Puerto Rican 1066 (PR1066) (Wellcome Trust (GR066133/GR080002), European Research Council (340755)), Puerto Rican Alzheimer Disease Initiative (PRADI) (RF1AG054074), Reasons for Geographic and Racial Differences in Stroke (REGARDS) (U01NS041588), Research in African American Alzheimer Disease Initiative (REAAADI) (U01AG052410), Rush Alzheimer's Disease Center (ROSMAP) (P30AG10161, R01AG15819, R01AG17919), University of Miami Brain Endowment Bank (MBB), and University of Miami/Case Western/North Carolina A&T African American (UM/CASE/NCAT) (U01AG052410, R01AG028786).

The four LSACs are: the Human Genome Sequencing Center at the Baylor College of Medicine (U54 HG003273), the Broad Institute Genome Center (U54HG003067), The American Genome Center at the Uniformed Services University of the Health Sciences (U01AG057659), and the Washington University Genome Institute (U54HG003079).

Biological samples and associated phenotypic data used in primary data analyses were stored at Study Investigators institutions, and at the National Cell Repository for Alzheimer's Disease (NCRAD, U24AG021886) at Indiana University funded by NIA. Associated Phenotypic Data used in primary and secondary data analyses were provided by Study Investigators, the NIA funded Alzheimer's Disease

Centers (ADCs), and the National Alzheimer's Coordinating Center (NACC, U01AG016976) and the National Institute on Aging Genetics of Alzheimer's Disease Data Storage Site (NIAGADS, U24AG041689) at the University of Pennsylvania, funded by NIA This research was supported in part by the Intramural Research Program of the National Institutes of health, National Library of Medicine. Contributors to the Genetic Analysis Data included Study Investigators on projects that were individually funded by NIA, and other NIH institutes, and by private U.S. organizations, or foreign governmental or nongovernmental organizations.

An up to date acknowledgment statement can be found on the ADSP site: https://www.niagads.org/adsp/content/acknowledgement-statement.

Data collection and sharing for this project was funded by the Alzheimer's Disease Neuroimaging Initiative (ADNI) (National Institutes of Health Grant U01 AG024904) and DOD ADNI (Department of Defense award number W81XWH-12-2-0012). ADNI is funded by the National Institute on Aging, the National Institute of Biomedical Imaging and Bioengineering, and through generous contributions from the following: AbbVie, Alzheimer's Association; Alzheimer's Drug Discovery Foundation; Araclon Biotech; BioClinica, Inc.; Biogen; Bristol-Myers Squibb Company; CereSpir, Inc.; Cogstate; Eisai Inc.; Elan Pharmaceuticals, Inc.; Eli Lilly and Company; EuroImmun; F. Hoffmann-La Roche Ltd and its affiliated company Genentech, Inc.; Fujirebio; GE Healthcare; IXICO Ltd.; Janssen Alzheimer Immunotherapy Research & Development, LLC.; Johnson & Johnson Pharmaceutical Research & Development LLC.; Lumosity; Lundbeck; Merck & Co., Inc.; Meso Scale Diagnostics, LLC.; NeuroRx Research; Neurotrack Technologies; Novartis Pharmaceuticals Corporation; Pfizer Inc.; Piramal Imaging; Servier; Takeda Pharmaceutical Company; and Transition Therapeutics. The Canadian Institutes of Health Research is providing funds to support ADNI clinical sites in Canada. Private sector contributions are facilitated by the Foundation for the National Institutes of Health (www.fnih.org). The grantee organization is the Northern California Institute for Research and Education, and the study is coordinated by the Alzheimer's Therapeutic Research Institute at the University of Southern California. ADNI data are disseminated by the Laboratory for Neuro Imaging at the University of Southern California.

Additional information to include in an acknowledgment statement can be found on the LONI site: https://adni.loni.usc.edu/wp-content/uploads/how_to_apply/ADNI_Data_Use_Agreement.pdf.

The Alzheimer's Disease Genetics Consortium (ADGC) supported sample preparation, whole exome sequencing and data processing through NIA grant U01AG032984. Sequencing data generation and harmonization is supported by the Genome Center for Alzheimer's Disease, U54AG052427, and data sharing is supported by NIAGADS, U24AG041689. Samples from the National Centralized Repository for Alzheimer's Disease and Related Dementias (NCRAD), which receives government support under a cooperative agreement grant (U24 AG021886) awarded by the National Institute on Aging (NIA), were used in this study. We thank contributors who collected samples used in this study, as well as patients and their families, whose help and participation made this work possible. NIH grants supported enrollment and data collection for the individual studies including: GenerAAtions R01AG20688 (PI M. Daniele Fallin, PhD); Miami/Duke R01 AG027944, R01 AG028786 (PI Margaret A. Pericak-Vance, PhD); NC A&T P20 MD000546, R01 AG28786-01A1 (PI Goldie S. Byrd, PhD); Case Western (PI Jonathan L. Haines, PhD); MIRAGE R01 AG009029 (PI Lindsay A. Farrer, PhD); ROS P30AG10161, R01AG15819, R01AG30146, TGen (PI David A. Bennett, MD); MAP R01AG17917, R01AG15819, TGen (PI David A. Bennett, MD). The NACC database is funded by NIA/NIH Grant U01 AG016976. NACC data are contributed by the NIA-funded

ADCs: P30 AG019610 (PI Eric Reiman, MD), P30 AG013846 (PI Neil Kowall, MD), P30 AG062428-01 (PI James Leverenz, MD) P50 AG008702 (PI Scott Small, MD), P50 AG025688 (PI Allan Levey, MD, PhD), P50 AG047266 (PI Todd Golde, MD, PhD), P30 AG010133 (PI Andrew Saykin, PsyD), P50 AG005146 (PI Marilyn Albert, PhD), P30 AG062421-01 (PI Bradley Hyman, MD, PhD), P30 AG062422-01 (PI Ronald Petersen, MD, PhD), P50 AG005138 (PI Mary Sano, PhD), P30 AG008051 (PI Thomas Wisniewski, MD), P30 AG013854 (PI Robert Vassar, PhD), P30 AG008017 (PI Jeffrey Kaye, MD), P30 AG010161 (PI David Bennett, MD), P50 AG047366 (PI Victor Henderson, MD, MS), P30 AG010129 (PI Charles DeCarli, MD), P50 AG016573 (PI Frank LaFerla, PhD), P30 AG062429-01(PI James Brewer, MD, PhD), P50 AG023501 (PI Bruce Miller, MD), P30 AG035982 (PI Russell Swerdlow, MD), P30 AG028383 (PI Linda Van Eldik, PhD), P30 AG053760 (PI Henry Paulson, MD, PhD), P30 AG010124 (PI John Trojanowski, MD, PhD), P50 AG005133 (PI Oscar Lopez, MD), P50 AG005142 (PI Helena Chui, MD), P30 AG012300 (PI Roger Rosenberg, MD), P30 AG049638 (PI Suzanne Craft, PhD), P50 AG005136 (PI Thomas Grabowski, MD), P30 AG062715-01 (PI Sanjay Asthana, MD, FRCP), P50 AG005681 (PI John Morris, MD), P50 AG047270 (PI Stephen Strittmatter, MD, PhD).
This work was supported by grants from the National Institutes of Health (R01AG044546, P01AG003991, RF1AG053303, R01AG058501, U01AG058922, RF1AG058501 and R01AG057777). The recruitment and clinical characterization of research participants at Washington University were supported by NIH P50 AG05681, P01 AG03991, and P01 AG026276. This work was supported by access to equipment made possible by the Hope Center for Neurological Disorders, and the Departments of Neurology and Psychiatry at Washington University School of Medicine.
We thank the contributors who collected samples used in this study, as well as patients and their families, whose help and participation made this work possible. Members of the National Institute on Aging Late-Onset Alzheimer Disease/National Cell Repository for Alzheimer Disease (NIA-LOAD NCRAD) Family Study Group include the following: Richard Mayeux, MD, MSc; Martin Farlow, MD; Tatiana Foroud, PhD; Kelley Faber, MS; Bradley F. Boeve, MD; Neill R. Graff-Radford, MD; David A. Bennett, MD; Robert A. Sweet, MD; Roger Rosenberg, MD; Thomas D. Bird, MD; Carlos Cruchaga, PhD; and Jeremy M. Silverman, PhD.
This work was partially supported by grant funding from NIH R01 AG039700 and NIH P50 AG005136. Subjects and samples used here were originally collected with grant funding from NIH U24 AG026395, U24 AG021886, P50 AG008702, P01 AG007232, R37 AG015473, P30 AG028377, P50 AG05128, P50 AG16574, P30 AG010133, P50 AG005681, P01 AG003991, U01MH046281, U01 MH046290 and U01 MH046373. The funders had no role in study design, analysis or preparation of the manuscript. The authors declare no competing interests.
This work was supported by the National Institutes of Health (R01 AG027944, R01 AG028786 to MAPV, R01 AG019085 to JLH, P20 MD000546); a joint grant from the Alzheimer's Association (SG-14-312644) and the Fidelity Biosciences Research Initiative to MAPV; the BrightFocus Foundation (A2011048 to MAPV). NIA-LOAD Family-Based Study supported the collection of samples used in this study through NIH grants U24 AG026395 and R01 AG041797 and the MIRAGE cohort was supported through the NIH grants R01 AG025259 and R01 AG048927. We thank contributors, including the Alzheimer's disease Centers who collected samples used in this study, as well as patients and their families, whose help and participation made this work possible. Study design: HNC, BWK, JLH, MAPV; Sample collection: MLC, JMV, RMC, LAF, JLH, MAPV; Whole exome sequencing and Sanger sequencing: SR, PLW; Sequencing data analysis: HNC, BWK, KLHN, SR, MAK, JRG, ERM, GWB, MAPV; Statistical analysis: BWK, KLHN, JMJ, MAPV; Preparation of

manuscript: HNC, BWK. The authors jointly discussed the experimental results throughout the duration of the study. All authors read and approved the final manuscript.

Data collection and sharing for this project was supported by the Washington Heights-Inwood Columbia Aging Project (WHICAP, PO1AG07232, R01AG037212, RF1AG054023) funded by the National Institute on Aging (NIA) and by the National Center for Advancing Translational Sciences, National Institutes of Health, through Grant Number UL1TR001873. This manuscript has been reviewed by WHICAP investigators for scientific content and consistency of data interpretation with previous WHICAP Study publications. We acknowledge the WHICAP study participants and the WHICAP research and support staff for their contributions to this study.

This work was supported by grants from the National Institutes of Health (R01AG044546, P01AG003991, RF1AG053303, R01AG058501, U01AG058922, RF1AG058501 and R01AG057777). The recruitment and clinical characterization of research participants at Washington University were supported by NIH P50 AG05681, P01 AG03991, and P01 AG026276. This work was supported by access to equipment made possible by the Hope Center for Neurological Disorders, and the Departments of Neurology and Psychiatry at Washington University School of Medicine.

We thank the contributors who collected samples used in this study, as well as patients and their families, whose help and participation made this work possible. Members of the National Institute on Aging Late-Onset Alzheimer Disease/National Cell Repository for Alzheimer Disease (NIA-LOAD NCRAD) Family Study Group include the following: Richard Mayeux, MD, MSc; Martin Farlow, MD; Tatiana Foroud, PhD; Kelley Faber, MS; Bradley F. Boeve, MD; Neill R. Graff-Radford, MD; David A. Bennett, MD; Robert A. Sweet, MD; Roger Rosenberg, MD; Thomas D. Bird, MD; Carlos Cruchaga, PhD; and Jeremy M. Silverman, PhD.

This work was supported by grants from the National Institutes of Health (R01AG044546, P01AG003991, RF1AG053303, R01AG058501, U01AG058922, RF1AG058501 and R01AG057777). The recruitment and clinical characterization of research participants at Washington University were supported by NIH P50 AG05681, P01 AG03991, and P01 AG026276. This work was supported by access to equipment made possible by the Hope Center for Neurological Disorders, and the Departments of Neurology and Psychiatry at Washington University School of Medicine.

We thank the contributors who collected samples used in this study, as well as patients and their families, whose help and participation made this work possible. Members of the National Institute on Aging Late-Onset Alzheimer Disease/National Cell Repository for Alzheimer Disease (NIA-LOAD NCRAD) Family Study Group include the following: Richard Mayeux, MD, MSc; Martin Farlow, MD; Tatiana Foroud, PhD; Kelley Faber, MS; Bradley F. Boeve, MD; Neill R. Graff-Radford, MD; David A. Bennett, MD; Robert A. Sweet, MD; Roger Rosenberg, MD; Thomas D. Bird, MD; Carlos Cruchaga, PhD; and Jeremy M. Silverman, PhD.

Mayo RNAseq Study- Study data were provided by the following sources: The Mayo Clinic Alzheimer's Disease Genetic Studies, led by Dr. Nilufer Ertekin-Taner and Dr. Steven G. Younkin, Mayo Clinic, Jacksonville, FL using samples from the Mayo Clinic Study of Aging, the Mayo Clinic Alzheimer's Disease Research Center, and the Mayo Clinic Brain Bank. Data collection was supported through funding by NIA grants P50 AG016574, R01 AG032990, U01 AG046139, R01 AG018023, U01 AG006576, U01 AG006786, R01 AG025711, R01 AG017216, R01 AG003949, NINDS grant R01 NS080820, CurePSP Foundation, and support from Mayo Foundation. Study data includes samples collected through the Sun Health Research Institute Brain and Body Donation Program of Sun City, Arizona. The Brain and Body Donation Program is supported by the National Institute of Neurological Disorders and Stroke (U24 NS072026 National Brain and Tissue Resource for Parkinson's Disease and Related Disorders), the National Institute on Aging (P30 AG19610 Arizona Alzheimer's Disease Core Center), the Arizona Department of Health Services (contract 211002, Arizona Alzheimer's Research Center), the Arizona Biomedical Research Commission (contracts 4001, 0011, 05-901 and 1001 to the Arizona Parkinson's Disease Consortium) and the Michael J. Fox Foundation for Parkinson's Research

ROSMAP- We are grateful to the participants in the Religious Order Study, the Memory and Aging Project. This work is supported by the US National Institutes of Health [U01 AG046152, R01 AG043617, R01 AG042210, R01 AG036042, R01 AG036836, R01 AG032990, R01 AG18023, RC2 AG036547, P50 AG016574, U01 ES017155, KL2 RR024151, K25 AG041906-01, R01 AG30146, P30 AG10161, R01 AG17917, R01 AG15819, K08 AG034290, P30 AG10161 and R01 AG11101.

Mount Sinai Brain Bank (MSBB)- This work was supported by the grants R01AG046170, RF1AG054014, RF1AG057440 and R01AG057907 from the NIH/National Institute on Aging (NIA). R01AG046170 is a component of the AMP-AD Target Discovery and Preclinical Validation Project. Brain tissue collection and characterization was supported by NIH HHSN271201300031C.

This study was supported by the National Institute on Aging (NIA) grants AG030653, AG041718, AG064877 and P30-AG066468.

We would like to thank study participants, their families, and the sample collectors for their invaluable contributions. This research was supported in part by the National Institute on Aging grant U01AG049508 (PI Alison M. Goate). This research was supported in part by Genentech, Inc. (PI Alison M. Goate, Robert R. Graham).

The NACC database is funded by NIA/NIH Grant U01 AG016976. NACC data are contributed by these NIA-funded ADCs: P30 AG013846 (PI Neil Kowall, MD), P50 AG008702 (PI Scott Small, MD), P50 AG025688 (PI Allan Levey, MD, PhD), P30 AG010133 (PI Andrew Saykin, PsyD), P50 AG005146 (PI Marilyn Albert, PhD), P50 AG005134 (PI Bradley Hyman, MD, PhD), P50 AG016574 (PI Ronald Petersen, MD, PhD), P30 AG013854 (PI M. Marsel Mesulam, MD), P30 AG008017 (PI Jeffrey Kaye, MD), P30 AG010161 (PI David Bennett, MD), P30 AG010129 (PI Charles DeCarli, MD), P50 AG016573 (PI Frank LaFerla, PhD), P50 AG005131 (PI Douglas Galasko, MD), P30 AG028383 (PI Linda Van Eldik, PhD), P30 AG010124 (PI John Trojanowski, MD, PhD), P50 AG005142 (PI Helena Chui, MD), P30 AG012300 (PI Roger Rosenberg, MD), P50 AG005136 (PI Thomas Grabowski, MD), P50 AG005681 (PI John Morris, MD), P30 AG028377 (Kathleen Welsh-Bohmer, PhD), and P50 AG008671 (PI Henry Paulson, MD, PhD).

Samples from the National Cell Repository for Alzheimer’s Disease (NCRAD), which receives government support under a cooperative agreement grant (U24 AG21886) awarded by the National Institute on Aging (NIA), were used in this study. We thank contributors who collected samples used in this study, as well as patients and their families, whose help and participation made this work possible.

The Alzheimer's Disease Genetics Consortium supported the collection of samples used in this study through National Institute on Aging (NIA) grants U01AG032984 and RC2AG036528.

We acknowledge the generous contributions of the Cache County Memory Study participants. Sequencing for this study was funded by RF1AG054052 (PI: John S.K. Kauwe).

#### Acknowledgements for the use of GWAS data distributed by NIAGADS

The NIA Genetics of Alzheimer's Disease Data Storage Site (NIAGADS) is supported by a collaborative agreement from the National Institute on Aging, U24AG041689.

NG00047: The NIA supported this work through grants U01-AG032984, RC2-AG036528, U01-AG016976 (Dr Kukull); U24 AG026395, U24 AG026390, R01AG037212, R37 AG015473 (Dr Mayeux); K23AG034550 (Dr Reitz); U24-AG021886 (Dr Foroud); R01AG009956, RC2 AG036650 (Dr Hall); UO1 AG06781, UO1 HG004610 (Dr Larson); R01 AG009029 (Dr Farrer); 5R01AG20688 (Dr Fallin); P50 AG005133, AG030653 (Dr Kamboh); R01 AG019085 (Dr Haines); R01 AG1101, R01 AG030146, RC2 AG036650 (Dr Evans); P30AG10161, R01AG15819, R01AG30146, R01AG17917, R01AG15819 (Dr Bennett); R01AG028786 (Dr Manly); R01AG22018, P30AG10161 (Dr Barnes); P50AG16574 (Dr Ertekin-Taner, Dr Graff-Radford), R01 AG032990 (Dr Ertekin-Taner), KL2 RR024151 (Dr Ertekin-Taner); R01 AG027944, R01 AG028786 (Dr Pericak-Vance); P20 MD000546, R01 AG28786-01A1 (Dr Byrd); AG005138 (Dr Buxbaum); P50 AG05681, P01 AG03991, P01 AG026276 (Dr Goate); and P30AG019610, P30AG13846, U01-AG10483, R01CA129769, R01MH080295, R01AG017173, R01AG025259, R01AG33193, P50AG008702, P30AG028377, AG05128, AG025688, P30AG10133, P50AG005146, P50AG005134, P01AG002219, P30AG08051, MO1RR00096, UL1RR029893, P30AG013854, P30AG008017, R01AG026916, R01AG019085, P50AG016582, UL1RR02777, R01AG031581, P30AG010129, P50AG016573, P50AG016575, P50AG016576, P50AG016577, P50AG016570, P50AG005131, P50AG023501, P50AG019724, P30AG028383, P50AG008671, P30AG010124, P50AG005142, P30AG012300, AG010491, AG027944, AG021547, AG019757, P50AG005136 (Alzheimer Disease GeneticsConsortium [ADGC]). We thank Creighton Phelps, Stephen Synder, and Marilyn Miller from the NIA, who are ex-officio members of the ADGC. Support was also provided by the Alzheimer's Association (IIRG-08-89720 [Dr Farrer] and IIRG-05-14147 [Dr Pericak-Vance]), National Institute of Neurological Disorders and Stroke grant NS39764, National Institute of Mental Health grant MH60451, GlaxoSmithKline, and the Office of Research and Development, Biomedical Laboratory Research Program, US Department of Veterans Affairs Administration. For the ADGC, biological samples and associated phenotypic data used in primary data analyses were stored at principal investigators' institutions and at the National Cell Repository for Alzheimer's Disease (NCRAD) at Indiana University, funded by the NIA. Associated phenotypic data used in secondary data analyses were stored at the National Alzheimer's Coordinating Center and at the NIA Alzheimer's Disease Data Storage Site at the University of Pennsylvania, funded by the NIA. Contributors to the genetic analysis data included principal investigators on projects ndividually funded by the NIA, other NIH institutes, or private entities.

#### Acknowledgements for other GWAS and phenotype data

##### *NACC*

The NACC database is funded by NIA/NIH Grant U01 AG016976. NACC data are contributed by the NIA-funded ADCs: P30 AG019610 (PI Eric Reiman, MD), P30 AG013846 (PI Neil Kowall, MD), P30 AG062428-01 (PI James Leverenz, MD) P50 AG008702 (PI Scott Small, MD), P50 AG025688 (PI Allan Levey, MD, PhD), P50 AG047266 (PI Todd Golde, MD, PhD), P30 AG010133 (PI Andrew Saykin, PsyD), P50 AG005146 (PI Marilyn Albert, PhD), P30 AG062421-01 (PI Bradley Hyman, MD, PhD), P30 AG062422-01 (PI Ronald Petersen, MD, PhD), P50 AG005138 (PI Mary Sano, PhD), P30 AG008051 (PI Thomas Wisniewski, MD), P30 AG013854 (PI Robert Vassar, PhD), P30 AG008017 (PI Jeffrey Kaye, MD), P30 AG010161 (PI David Bennett, MD), P50 AG047366 (PI Victor Henderson, MD, MS), P30 AG010129 (PI Charles DeCarli, MD), P50 AG016573 (PI Frank LaFerla, PhD), P30 AG062429-01(PI James Brewer, MD, PhD), P50 AG023501 (PI Bruce

Miller, MD), P30 AG035982 (PI Russell Swerdlow, MD), P30 AG028383 (PI Linda Van Eldik, PhD), P30 AG053760 (PI Henry Paulson, MD, PhD), P30 AG010124 (PI John Trojanowski, MD, PhD), P50 AG005133 (PI Oscar Lopez, MD), P50 AG005142 (PI Helena Chui, MD), P30 AG012300 (PI Roger Rosenberg, MD), P30 AG049638 (PI Suzanne Craft, PhD), P50 AG005136 (PI Thomas Grabowski, MD), P30 AG062715-01 (PI Sanjay Asthana, MD, FRCP), P50 AG005681 (PI John Morris, MD), P50 AG047270 (PI Stephen Strittmatter, MD, PhD).

##### *MARS & LATC*

We thank all Minority Aging Research Study and Latino Core participants and the Rush Alzheimer's Disease Center staff. This database was funded by the NIH/NIA grants R01AG22018 (MARS) and P30AG 072975 (ADC).

##### *GenADA*

The genotypic and associated phenotypic data used in the study "Multi-Site Collaborative Study for Genotype-Phenotype Associations in Alzheimer's Disease (GenADA)" were provided by the GlaxoSmithKline, R&D Limited.

##### *ROSMAP*

ROSMAP study data were provided by the Rush Alzheimer's Disease Center, Rush University Medical Center, Chicago. Data collection was supported through funding by NIA grants P30AG10161, R01AG15819, R01AG17917, R01AG30146, R01AG36836, U01AG32984, U01AG46152, the Illinois Department of Public Health, and the Translational Genomics Research Institute.

##### *AddNeuroMed*

The AddNeuroMed data are from a public-private partnership supported by EFPIA companies and SMEs as part of InnoMed (Innovative Medicines in Europe), an Integrated Project funded by the European Union of the Sixth Framework program priority FP6-2004-LIFESCIHEALTH-5. Clinical leads responsible for data collection are Iwona Kłoszewska (Lodz), Simon Lovestone (London), Patrizia Mecocci (Perugia), Hilkka Soininen (Kuopio), Magda Tsolaki (Thessaloniki), and Bruno Vellas (Toulouse), imaging leads are Andy Simmons (London), Lars-Olad Wahlund (Stockholm) and Christian Spenger (Zurich) and bioinformatics leads are Richard Dobson (London) and Stephen Newhouse (London).

##### *ADNI*

Data collection and sharing for this project was funded by the Alzheimer's Disease Neuroimaging Initiative (ADNI) (National Institutes of Health Grant U01 AG024904) and DOD ADNI (Department of Defense award number W81XWH-12-2-0012). ADNI is funded by the National Institute on Aging, the National Institute of Biomedical Imaging and Bioengineering and through generous contributions from the following: AbbVie. Alzheimer's Association; Alzheimer's Drug Discovery Foundation; Araclon Biotech; BioClinica. Inc.; Biogen; Bristol-Myers Squibb Company; CereSpir. Inc.; Cogstate; Eisai Inc.; Elan Pharmaceuticals. Inc.; Eli Lilly and Company; EuroImmun; F. Hoffmann-La Roche Ltd and its affiliated company Genentech. Inc.; Fujirebio; GE HealtControlsare; IXICO Ltd.; Janssen Alzheimer Immunotherapy Research & Development. LLC.; Johnson & Johnson Pharmaceutical Research & Development LLC.; Lumosity; Lundbeck; Merck & Co. Inc.; Meso Scale Diagnostics. LLC.; NeuroRx Research; Neurotrack Technologies; Novartis Pharmaceuticals Corporation; Pfizer Inc.; Piramal Imaging; Servier; Takeda Pharmaceutical Company; and Transition

Therapeutics. The Canadian Institutes of Health Research is providing funds to support ADNI clinical sites in Canada. Private sector contributions are facilitated by the Foundation for the National Institutes of Health. The grantee organization is the Northern California Institute for Research and Education, and the study is coordinated by the Alzheimer's Therapeutic Research Institute at the University of Southern California. ADNI data are disseminated by the Laboratory for Neuro Imaging at the University of Southern California.

##### *NCRAD*

Biological samples used in this study were stored at study investigators' institutions and at the National Cell Repository for Alzheimer's Disease (NCRAD) at Indiana University, which receives government support under a cooperative agreement grant (U24 AG21886) awarded by the National Institute on Aging (NIA). We thank contributors who collected samples used in this study, as well as patients and their families, whose help and participation made this work possible.

##### *UK Biobank*

UK Biobank data were analyzed under Application Number 45420.

##### *FinnGen Study*

We want to acknowledge the participants and investigators of the FinnGen study. The FinnGen project is funded by two grants from Business Finland (HUS 4685/31/2016 and UH 4386/31/2016) and the following industry partners: AbbVie Inc., Alnylam Pharmaceuticals, Inc., AstraZeneca UK Ltd, Bayer AG, Biogen MA Inc., Boehringer Ingelheim International GmbH, Bristol Myers Squibb Inc. (and Celgene Corporation & Celgene International II Sàrl), Genentech Inc., GlaxoSmithKline Intellectual Property Development Ltd., Johnson&Johnson Innovative Medicine Inc., Maze Therapeutics Inc., Merck Sharp & Dohme LCC, Novartis AG, Pfizer Inc. and Sanofi US Services Inc. Following biobanks are acknowledged for delivering biobank samples to FinnGen: Auria Biobank (www.auria.fi/biopankki), THL Biobank (www.thl.fi/biobank), Helsinki Biobank (www.helsinginbiopankki.fi), Biobank Borealis of Northern Finland (https://www.ppshp.fi/Tutkimus-ja-opetus/Biopankki/Pages/Biobank-Borealis-briefly-in-English.aspx), Finnish Clinical Biobank Tampere (www.tays.fi/en-US/Research_and_development/Finnish_Clinical_Biobank_Tampere), Biobank of Eastern Finland (www.ita-suomenbiopankki.fi/en), Central Finland Biobank (www.ksshp.fi/fi-FI/Potilaalle/Biopankki), Finnish Red Cross Blood Service Biobank (www.veripalvelu.fi/verenluovutus/biopankkitoiminta), Terveystalo Biobank (www.terveystalo.com/fi/Yritystietoa/Terveystalo-Biopankki/Biopankki/) and Arctic Biobank (https://www.oulu.fi/en/university/faculties-and-units/faculty-medicine/northern-finland-birth-cohorts-and-arctic-biobank). All Finnish Biobanks are members of BBMRI.fi infrastructure (https://www.bbmri-eric.eu/national-nodes/finland/). Finnish Biobank Cooperative -FINBB (https://finbb.fi/) is the coordinator of BBMRI-ERIC operations in Finland. The Finnish biobank data can be accessed through the Fingenious® services (https://site.fingenious.fi/en/) managed by FINBB.

##### *FinnGen ethics statement*

Study subjects in FinnGen provided informed consent for biobank research, based on the Finnish Biobank Act. Alternatively, separate research cohorts, collected prior the Finnish Biobank Act came into effect (in September 2013) and start of FinnGen (August 2017), were collected based on study-specific consents

and later transferred to the Finnish biobanks after approval by Fimea (Finnish Medicines Agency), the National Supervisory Authority for Welfare and Health. Recruitment protocols followed the biobank protocols approved by Fimea. The Coordinating Ethics Committee of the Hospital District of Helsinki and Uusimaa (HUS) statement number for the FinnGen study is Nr HUS/990/2017.
The FinnGen study is approved by Finnish Institute for Health and Welfare (permit numbers: THL/2031/6.02.00/2017, THL/1101/5.05.00/2017, THL/341/6.02.00/2018, THL/2222/6.02.00/2018, THL/283/6.02.00/2019, THL/1721/5.05.00/2019 and THL/1524/5.05.00/2020), Digital and population data service agency (permit numbers: VRK43431/2017-3, VRK/6909/2018-3, VRK/4415/2019-3), the Social Insurance Institution (permit numbers: KELA 58/522/2017, KELA 131/522/2018, KELA 70/522/2019, KELA 98/522/2019, KELA 134/522/2019, KELA 138/522/2019, KELA 2/522/2020, KELA 16/522/2020), Findata permit numbers THL/2364/14.02/2020, THL/4055/14.06.00/2020, THL/3433/14.06.00/2020, THL/4432/14.06/2020, THL/5189/14.06/2020, THL/5894/14.06.00/2020, THL/6619/14.06.00/2020, THL/209/14.06.00/2021, THL/688/14.06.00/2021, THL/1284/14.06.00/2021, THL/1965/14.06.00/2021, THL/5546/14.02.00/2020, THL/2658/14.06.00/2021, THL/4235/14.06.00/2021, Statistics Finland (permit numbers: TK-53-1041-17 and TK/143/07.03.00/2020 (earlier TK-53-90-20) TK/1735/07.03.00/2021, TK/3112/07.03.00/2021) and Finnish Registry for Kidney Diseases permission/extract from the meeting minutes on 4th July 2019.
The Biobank Access Decisions for FinnGen samples and data utilized in FinnGen Data Freeze 12 include: THL Biobank BB2017_55, BB2017_111, BB2018_19, BB_2018_34, BB_2018_67, BB2018_71, BB2019_7, BB2019_8, BB2019_26, BB2020_1, BB2021_65, Finnish Red Cross Blood Service Biobank 7.12.2017, Helsinki Biobank HUS/359/2017, HUS/248/2020, HUS/430/2021 §28, §29, HUS/150/2022 §12, §13, §14, §15, §16, §17, §18, §23, §58, §59, HUS/128/2023 §18, Auria Biobank AB17-5154 and amendment #1 (August 17 2020) and amendments BB_2021-0140, BB_2021-0156 (August 26 2021, Feb 2 2022), BB_2021-0169, BB_2021-0179, BB_2021-0161, AB20-5926 and amendment #1 (April 23 2020) and it´s modifications (Sep 22 2021), BB_2022-0262, BB_2022-0256, Biobank Borealis of Northern Finland_2017_1013, 2021_5010, 2021_5010 Amendment, 2021_5018, 2021_5018 Amendment, 2021_5015, 2021_5015 Amendment, 2021_5015 Amendment_2, 2021_5023, 2021_5023 Amendment, 2021_5023 Amendment_2, 2021_5017, 2021_5017 Amendment, 2022_6001, 2022_6001 Amendment, 2022_6006 Amendment, 2022_6006 Amendment, 2022_6006 Amendment_2, BB22-0067, 2022_0262, 2022_0262 Amendment, Biobank of Eastern Finland 1186/2018 and amendment 22§/2020, 53§/2021, 13§/2022, 14§/2022, 15§/2022, 27§/2022, 28§/2022, 29§/2022, 33§/2022, 35§/2022, 36§/2022, 37§/2022, 39§/2022, 7§/2023, 32§/2023, 33§/2023, 34§/2023, 35§/2023, 36§/2023, 37§/2023, 38§/2023, 39§/2023, 40§/2023, 41§/2023, Finnish Clinical Biobank Tampere MH0004 and amendments (21.02.2020 & 06.10.2020), BB2021-0140 8§/2021, 9§/2021, §9/2022, §10/2022, §12/2022, 13§/2022, §20/2022, §21/2022, §22/2022, §23/2022, 28§/2022, 29§/2022, 30§/2022, 31§/2022, 32§/2022, 38§/2022, 40§/2022, 42§/2022, 1§/2023, Central Finland Biobank 1-2017, BB_2021-0161, BB_2021-0169, BB_2021-0179, BB_2021-0170, BB_2022-0256, BB_2022-0262, BB22-0067, Decision allowing to continue data processing until 31st Aug 2024 for projects: BB_2021-0179, BB22-0067,BB_2022-0262, BB_2021-0170, BB_2021-0164, BB_2021-0161, and BB_2021-0169, and Terveystalo Biobank STB 2018001 and amendment 25th Aug 2020, Finnish Hematological Registry and Clinical Biobank decision 18th June 2021, Arctic biobank P0844: ARC_2021_1001.

#### *VA Million Veteran Program*

**MVP Program Office**

- Sumitra Muralidhar, Ph.D., Program Director
US Department of Veterans Affairs, 810 Vermont Avenue NW, Washington, DC 20420
- Jennifer Moser, Ph.D., Associate Director, Scientific Programs
US Department of Veterans Affairs, 810 Vermont Avenue NW, Washington, DC 20420
- Jennifer E. Deen, B.S., Associate Director, Cohort & Public Relations
US Department of Veterans Affairs, 810 Vermont Avenue NW, Washington, DC 20420

**MVP Steering Committee**

- Co-Chair: Philip S. Tsao, Ph.D.
VA Palo Alto Health Care System, 3801 Miranda Avenue, Palo Alto, CA 94304
- Co-Chair: Sumitra Muralidhar, Ph.D.
US Department of Veterans Affairs, 810 Vermont Avenue NW, Washington, DC 20420
- J. Michael Gaziano, M.D., M.P.H.
VA Boston Healthcare System, 150 S. Huntington Avenue, Boston, MA 02130
- Adriana Hung, M.D., M.P.H.,
VA Tennessee Valley Healthcare System, 1310 24th Avenue, South Nashville, TN 37212
- Dave Oslin, M.D.
Philadelphia VA Medical Center, 3900 Woodland Avenue, Philadelphia, PA 19104
- Deepak Voora, M.D.
Durham VA Medical Center, 508 Fulton Street, Durham, NC 27705

**MVP Co-Principal Investigators**

- J. Michael Gaziano, M.D., M.P.H.
VA Boston Healthcare System, 150 S. Huntington Avenue, Boston, MA 02130
- Philip S. Tsao, Ph.D.
VA Palo Alto Health Care System, 3801 Miranda Avenue, Palo Alto, CA 94304

**MVP Core Operations**

- Jessica V. Brewer, M.P.H., Director, MVP Cohort Operations
VA Boston Healthcare System, 150 S. Huntington Avenue, Boston, MA 02130
- Mary T. Brophy M.D., M.P.H., Director, VA Central Biorepository
VA Boston Healthcare System, 150 S. Huntington Avenue, Boston, MA 02130
- Kelly Cho, M.P.H, Ph.D., Director, MVP Phenomics
VA Boston Healthcare System, 150 S. Huntington Avenue, Boston, MA 02130
- Lori Churby, B.S., Director, MVP Regulatory Affairs
VA Palo Alto Health Care System, 3801 Miranda Avenue, Palo Alto, CA 94304
- Jacob T. Kean, Ph.D., Acting Director, VA Informatics and Computing Infrastructure (VINCI)
VA Salt Lake City Health Care System, 500 Foothill Drive, Salt Lake City, UT 84148
- Saiju Pyarajan Ph.D., Director, Data and Computational Sciences
VA Boston Healthcare System, 150 S. Huntington Avenue, Boston, MA 02130
- Robert Ringer, Pharm.D., Director, VA Albuquerque Central Biorepository

New Mexico VA Health Care System, 1501 San Pedro Drive SE, Albuquerque, NM 87108
- Luis E. Selva, Ph.D., Director, MVP Biorepository Coordination
VA Boston Healthcare System, 150 S. Huntington Avenue, Boston, MA 02130
- Shahpoor (Alex) Shayan, M.S., Director, MVP PRE Informatics
VA Boston Healthcare System, 150 S. Huntington Avenue, Boston, MA 02130
- Brady Stephens, M.S., Principal Investigator, MVP Information Center
Canandaigua VA Medical Center, 400 Fort Hill Avenue, Canandaigua, NY 14424
- Stacey B. Whitbourne, Ph.D., Director, MVP Cohort Development and Management
VA Boston Healthcare System, 150 S. Huntington Avenue, Boston, MA 02130

### Supplementary Figures

#### XWAS

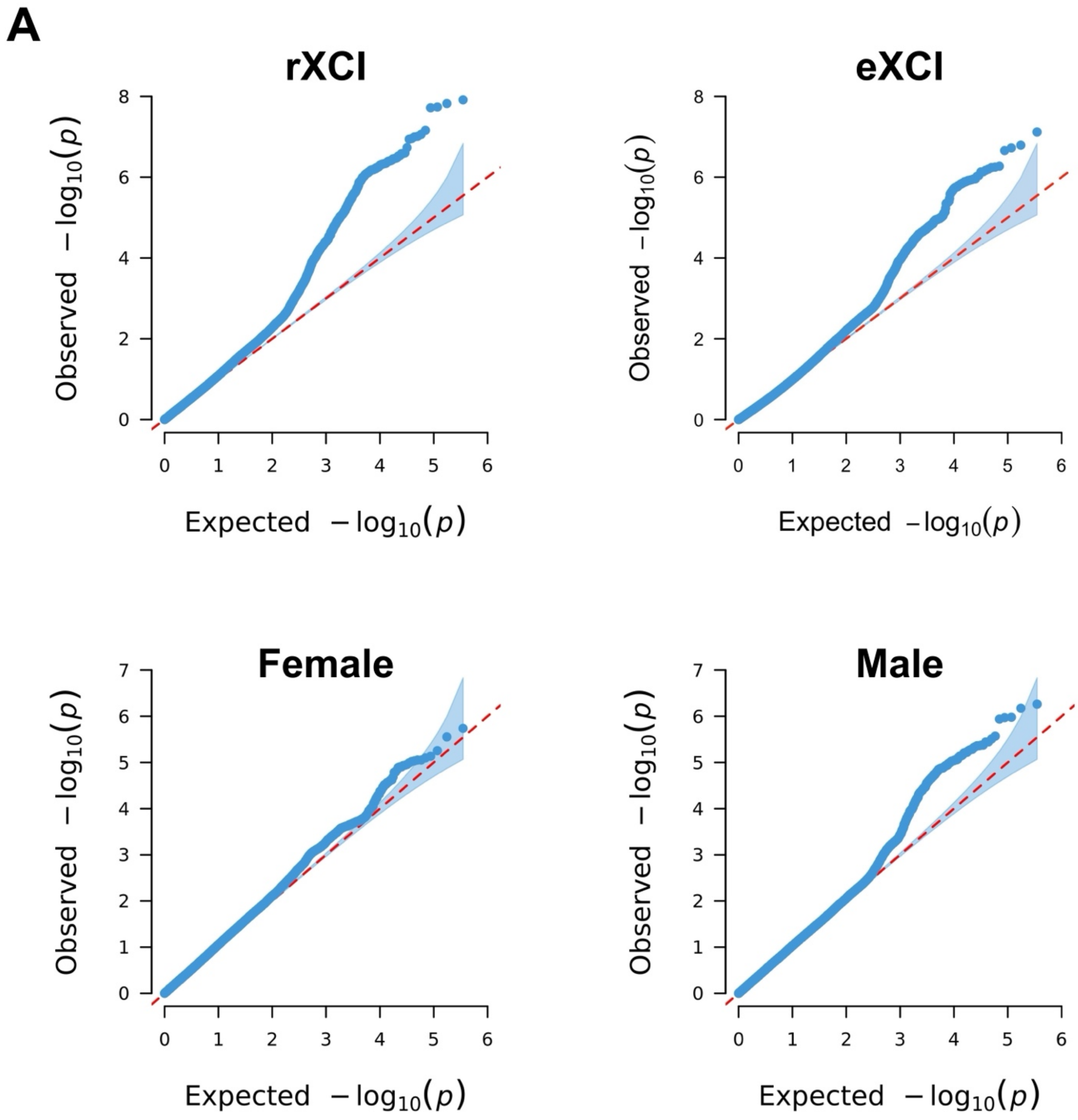

**B**

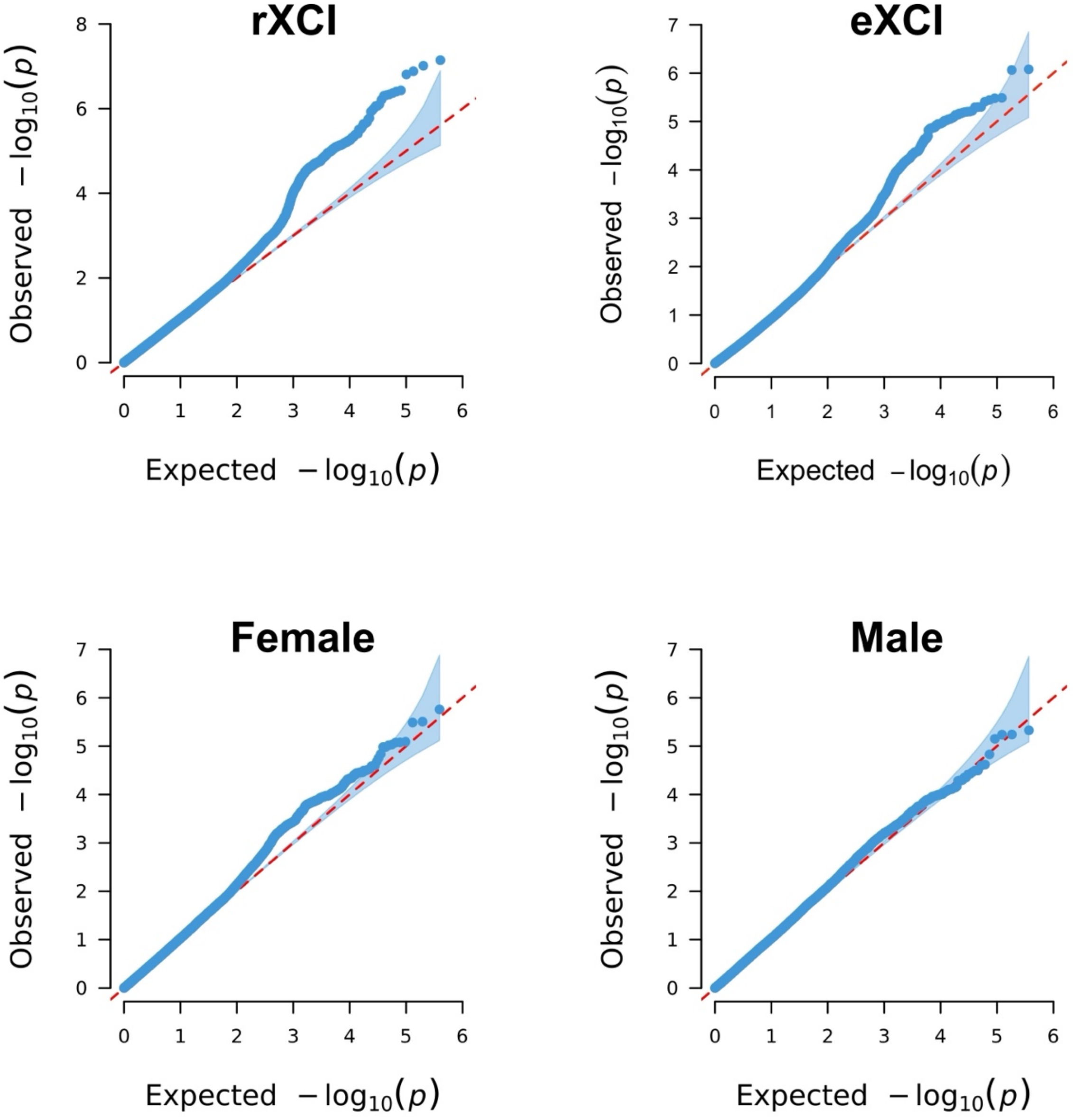
rXCI
eXCI
Female
Male
Observed $-\log_{10}(p)$
Expected $-\log_{10}(p)$
0
1
2
3
4
5
6
7
8

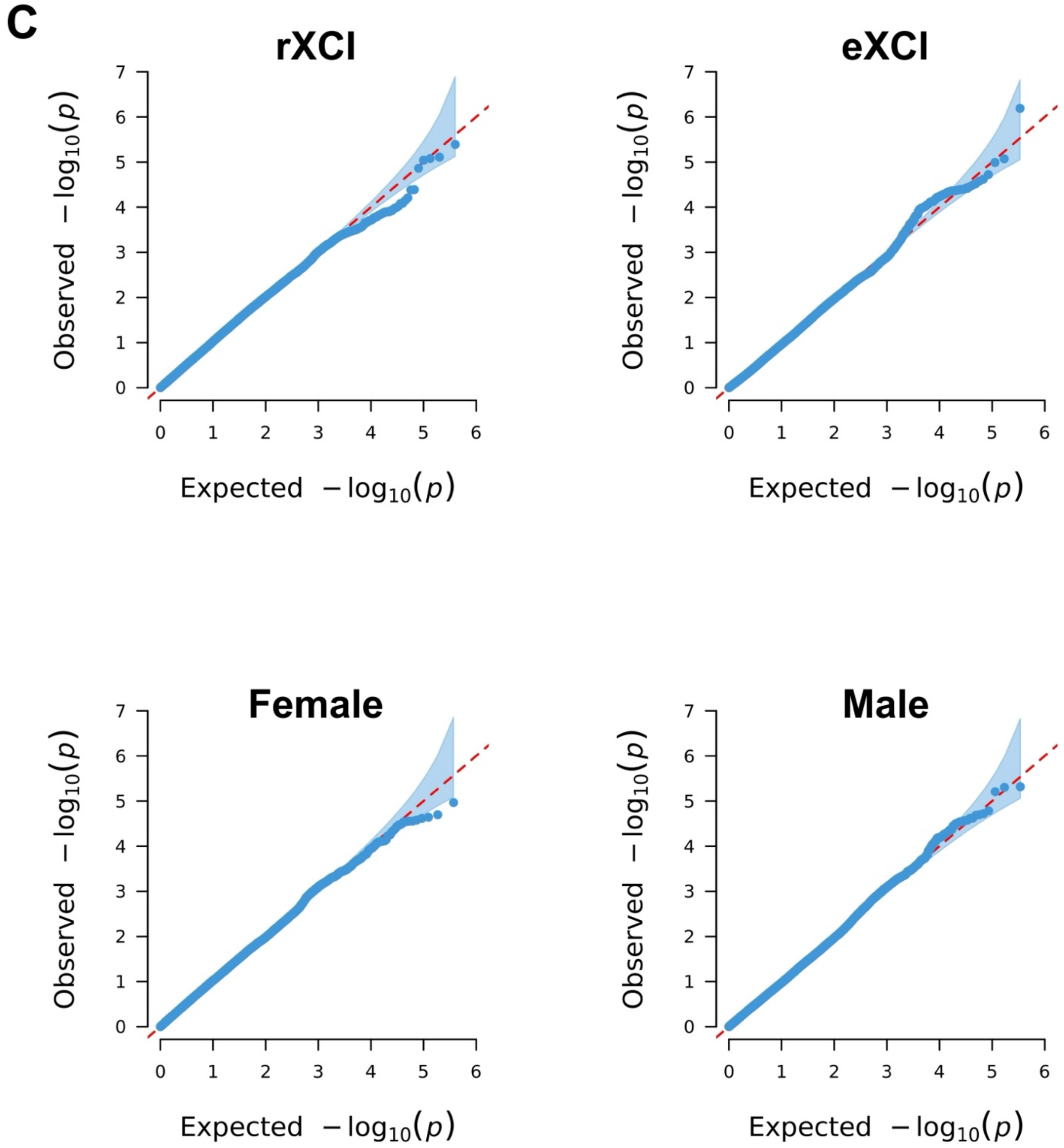

**Supplementary Figure 1. Quantile-quantile (QQ) plots of X-chromosome association analyses across genetic models and *APOE**4 strata.** Observed versus expected -$\log_{10}$(P) values from XWAS under four modeling frameworks: random X-chromosome inactivation (rXCI), escape from X-chromosome inactivation (eXCI), female-only, and male-only models. **A)** Non-*APOE**4-stratified, **B)** *APOE**4 negative, **C)** *APOE**4 positive. Across all 12 XWAS models, genomic inflation was minimal ($\lambda_{1000}$ range: 0.9994–1.0004).

## S2.1

### *NLGN4X* - Female

##### Index SNP: rs191558397

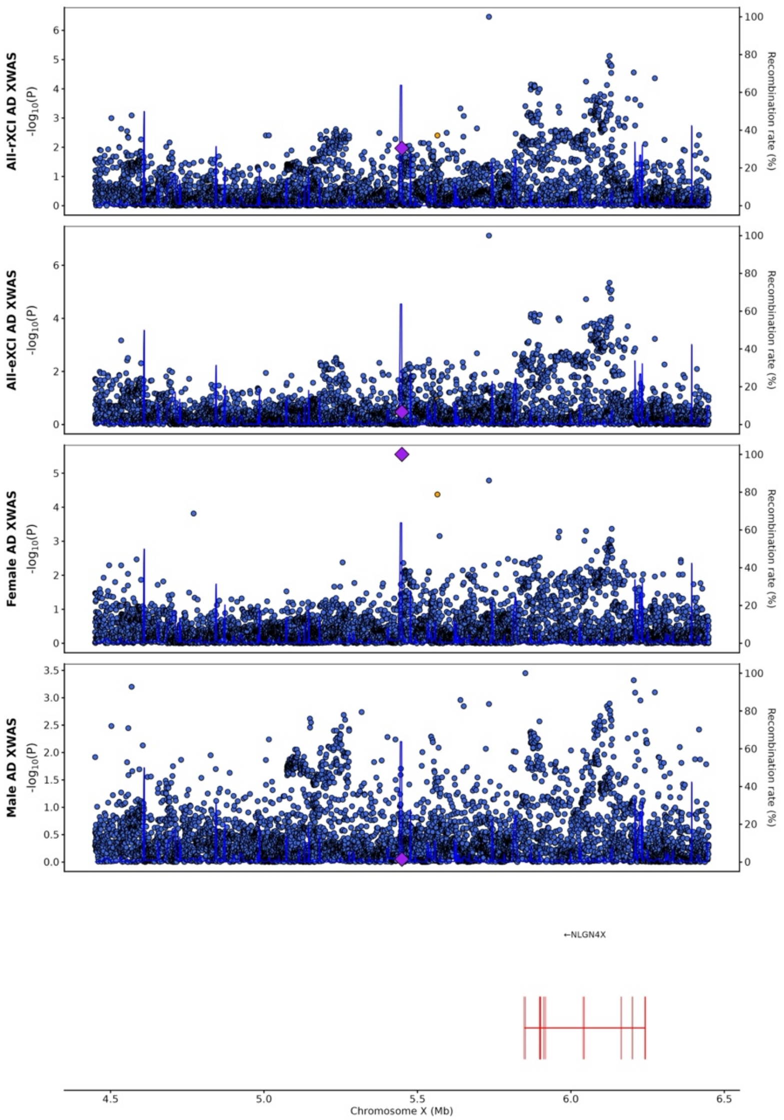

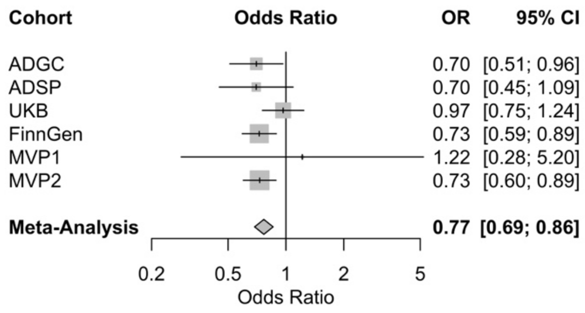

Heterogeneity: $I^2 = 0.0\%$, $\tau^2 = 0.0027$, $p = 0.4484$

**S2.2**

### *NLGN4X* - eXCI

#### Index SNP: rs150798997

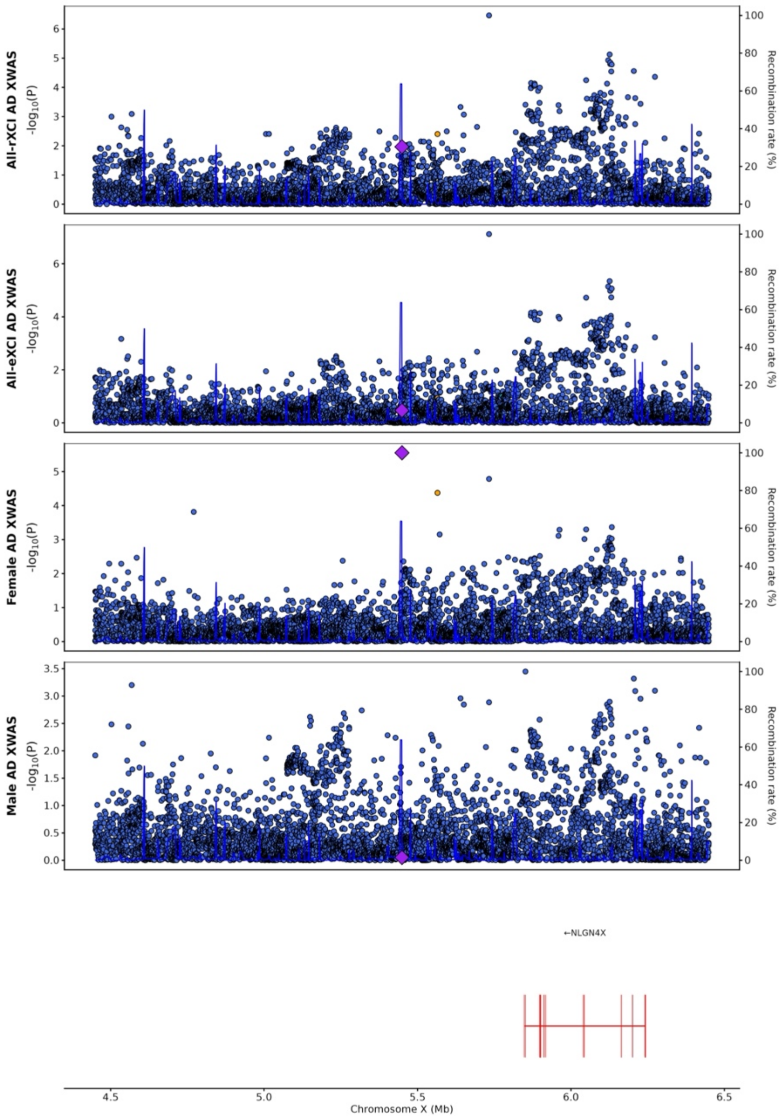

| Cohort | Odds Ratio | OR | 95% CI |
|---|---|---|---|
| ADGC | | 0.54 | [0.40; 0.74] |
| UKB | | 1.00 | [0.72; 1.39] |
| FinnGen | | 0.65 | [0.49; 0.86] |
| MVP | | 0.72 | [0.59; 0.87] |
| **Meta-Analysis** | | **0.70** | **[0.62; 0.80]** |

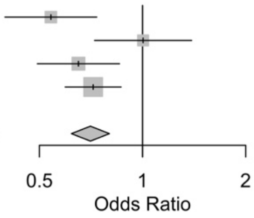

Heterogeneity: $I^2$ = 61.3%, $\tau^2$ = 0.0340, $p$ = 0.0516

## S2.3

### *NLGN4X* - eXCI

##### Index SNP: rs35566460

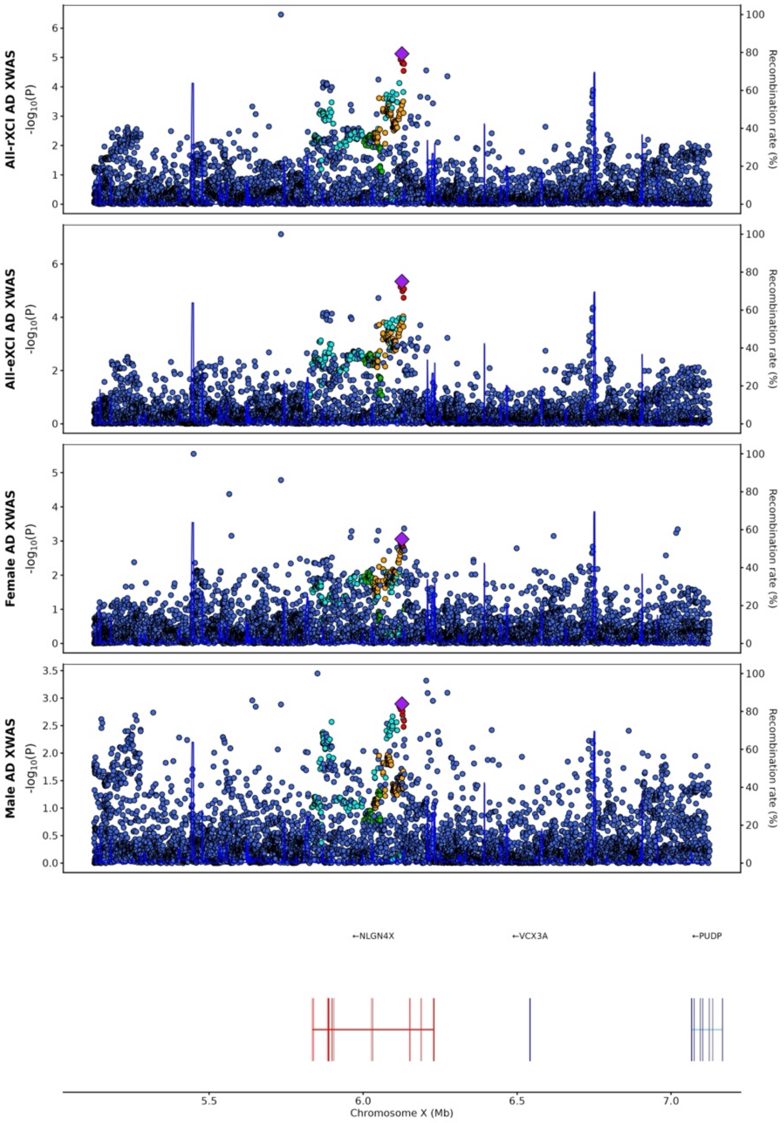

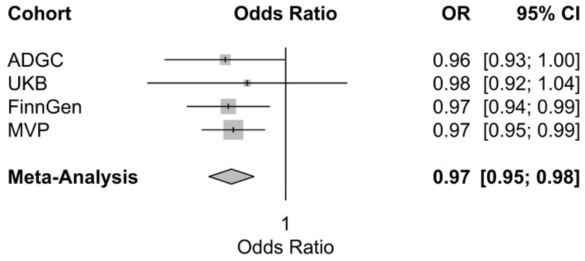

Heterogeneity: $I^2 = 0.0\%$, $\tau^2 = 0$, $p = 0.9819$

**S2.4**

### *MID1* - Female

#### Index SNP: rs12852495

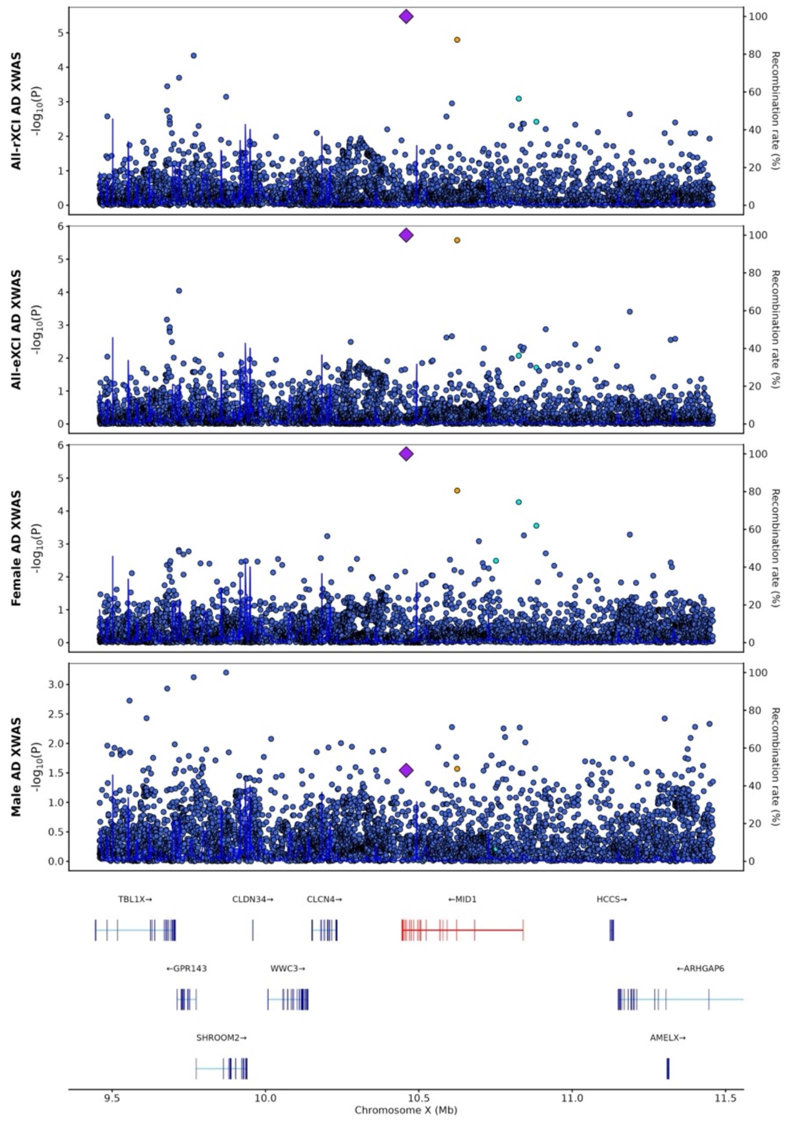

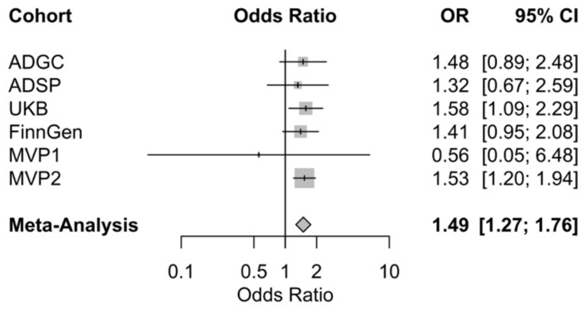

Heterogeneity: $I^2$ = 0.0%, $\tau^2$ = 0, $p$ = 0.9659

**S2.5**

#### *DMD* - eXCI

##### Index SNP: rs12014100

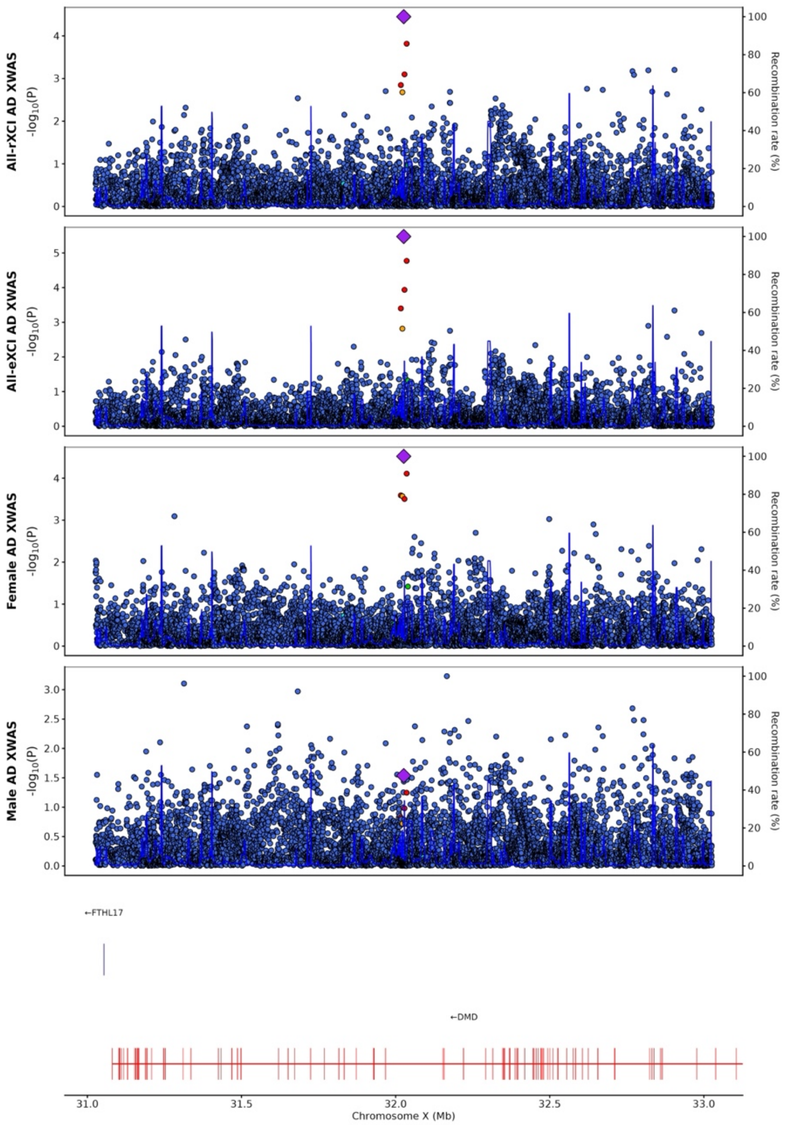

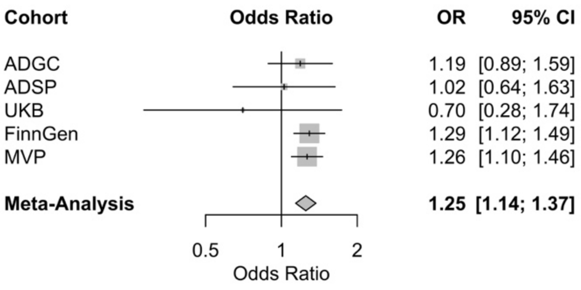

Heterogeneity: $I^2$ = 0.0%, $\tau^2$ < 0.0001, $p$ = 0.6277

**S2.6**

#### *SLC9A7* - rXCI

##### Index SNP: rs2142791

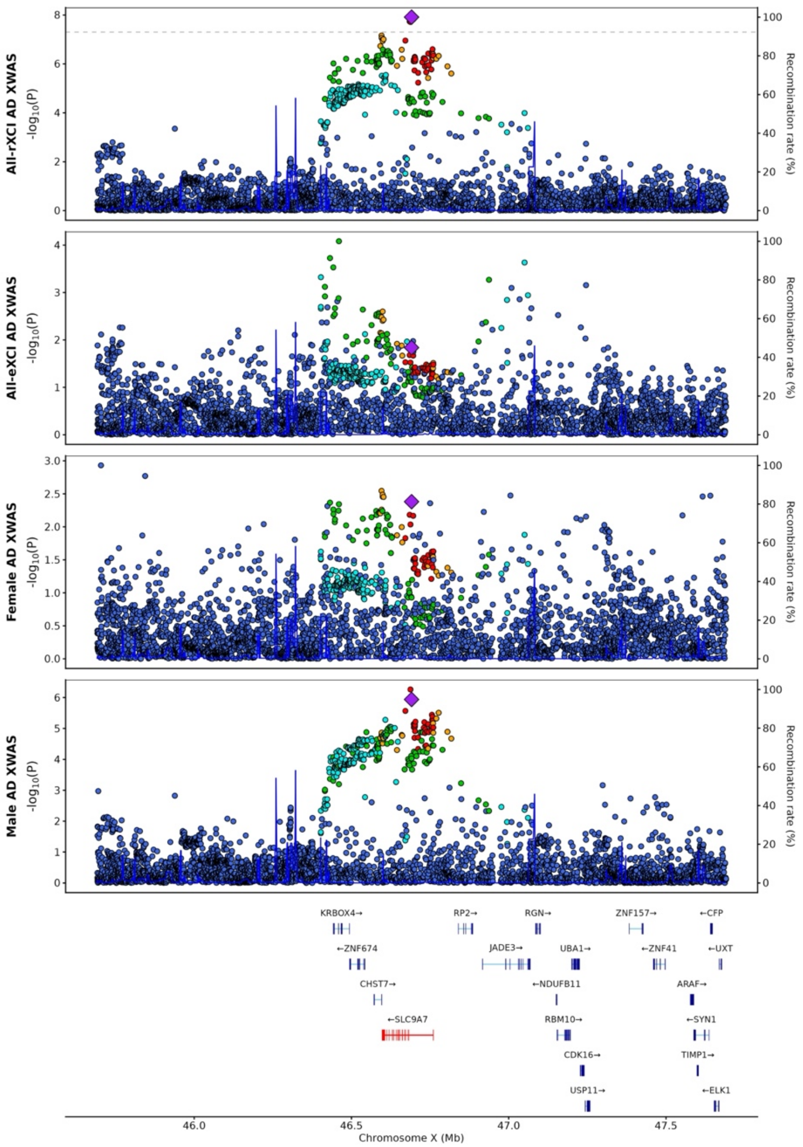

| Cohort | OR | 95% CI |
|---|---|---|
| ADGC | 1.02 | [0.99; 1.05] |
| ADSP | 1.01 | [0.95; 1.07] |
| UKB | 1.02 | [1.00; 1.05] |
| FinnGen | 1.01 | [0.99; 1.03] |
| MVP1 | 1.04 | [1.02; 1.05] |
| MVP2 | 1.04 | [1.01; 1.07] |
| **Meta-Analysis** | **1.03** | **[1.02; 1.04]** |

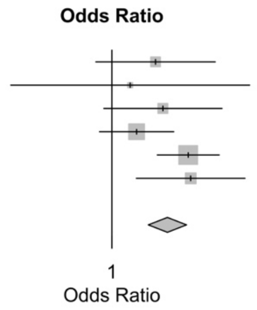

Heterogeneity: $I^2 = 11.3\%$, $\tau^2 < 0.0001$, $p = 0.3430$

**S2.7**

#### *DACH2* - Female

##### Index SNP: rs5923518

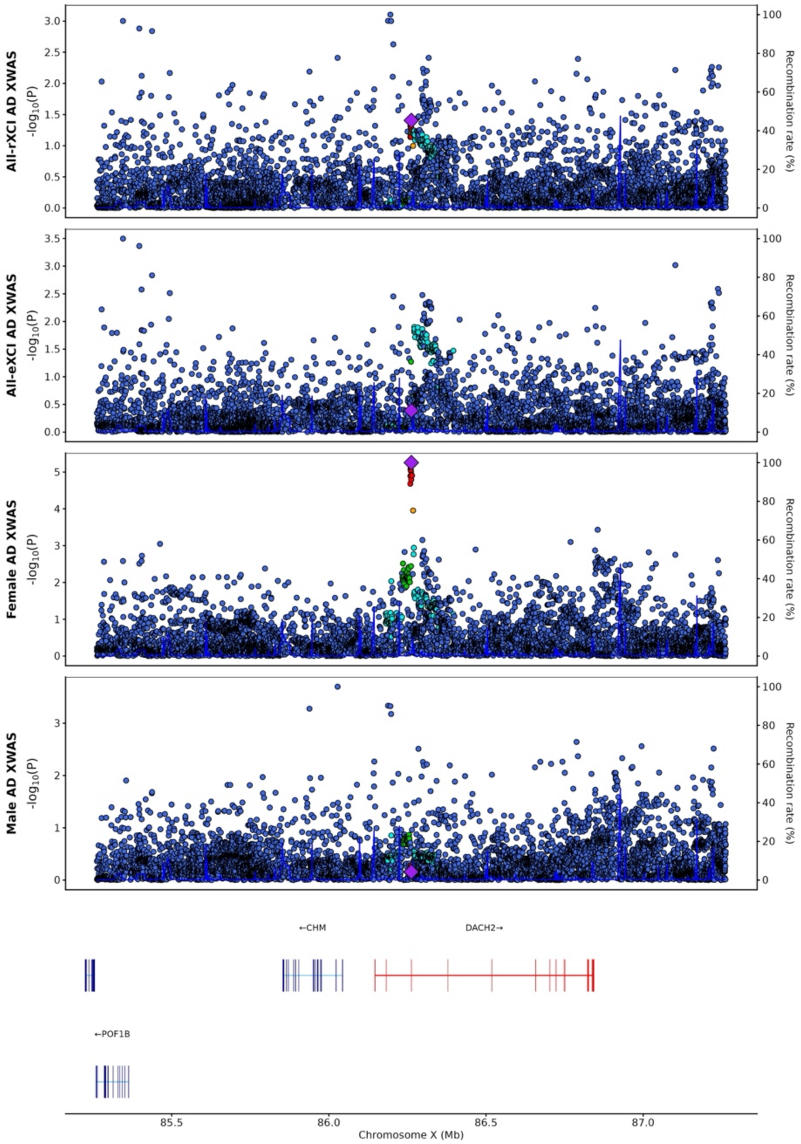

| Cohort | OR | 95% CI |
|---|---|---|
| ADGC | 0.93 | [0.89; 0.98] |
| ADSP | 0.95 | [0.86; 1.06] |
| UKB | 0.96 | [0.91; 1.00] |
| FinnGen | 0.97 | [0.93; 1.00] |
| MVP1 | 0.99 | [0.85; 1.15] |
| MVP2 | 0.96 | [0.93; 0.99] |
| **Meta-Analysis** | **0.96** | **[0.94; 0.98]** |

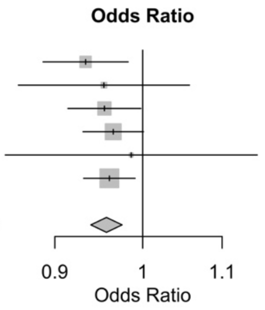

Heterogeneity: $I^2 = 0.0\%$, $\tau^2 = 0$, $p = 0.9323$

**S2.8**

#### *GLUD2* - Male

##### Index SNP: rs182269003

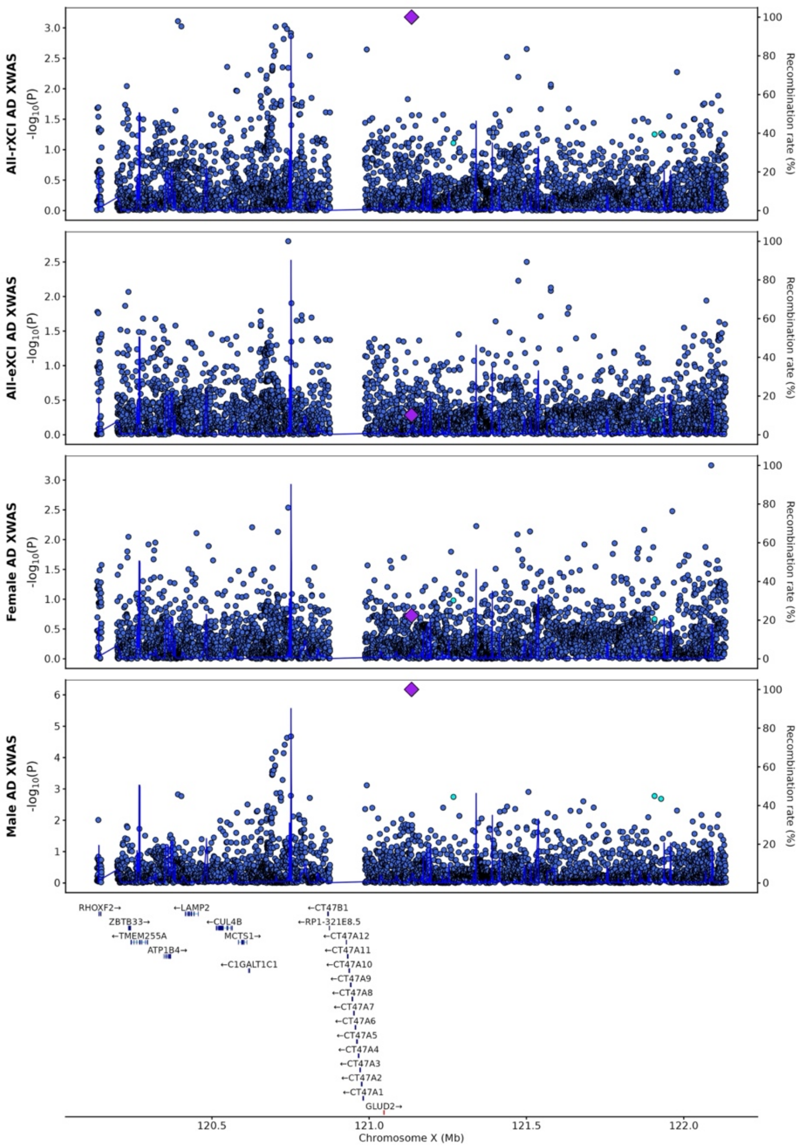

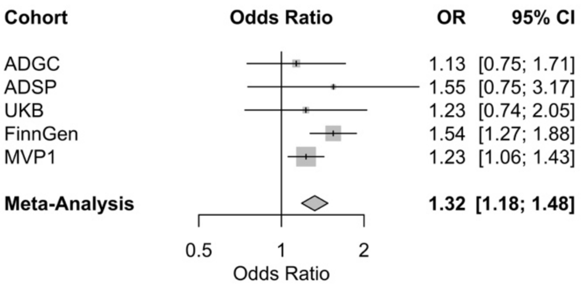

Heterogeneity: $I^2$ = 4.0%, $\tau^2$ = 0.0080, $p$ = 0.3837

**S2.9**

### *ZNF280C* - eXCI

#### Index SNP: rs34923017

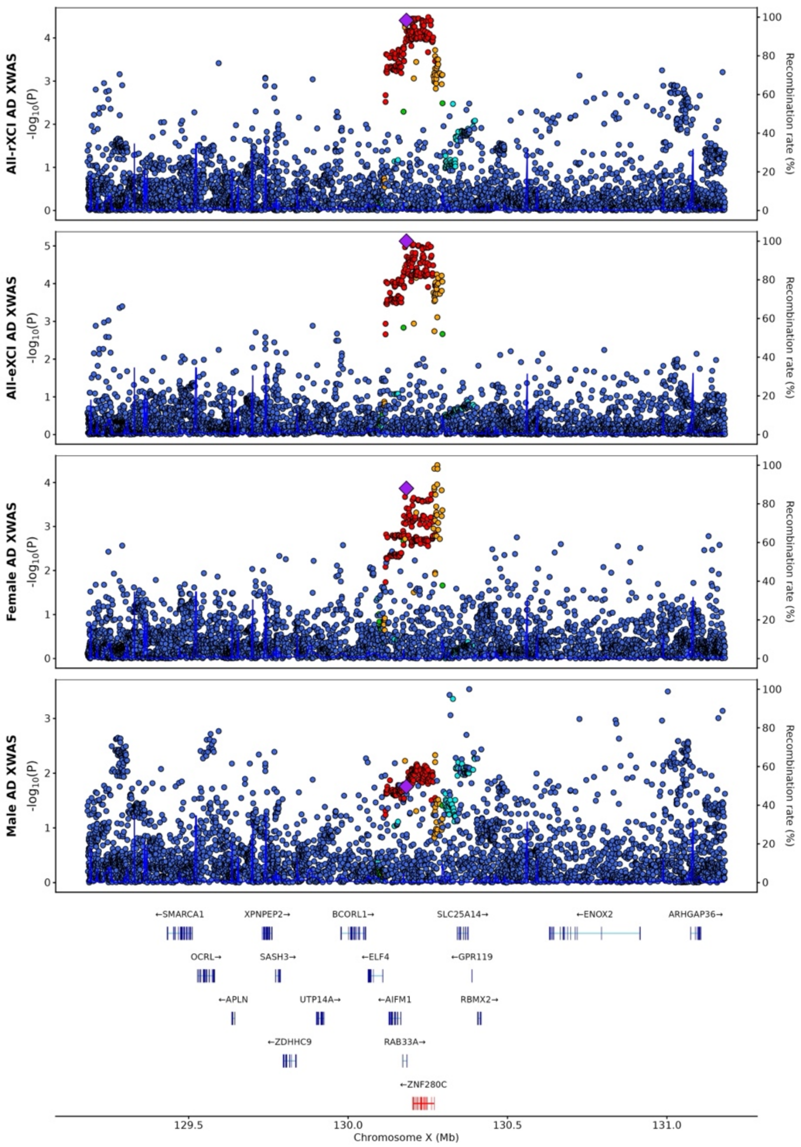

| Cohort | Odds Ratio | OR | 95% CI |
|---|---|---|---|
| ADGC | | 1.04 | [1.00; 1.08] |
| UKB | | 1.03 | [0.99; 1.06] |
| FinnGen | | 1.02 | [1.00; 1.05] |
| MVP | | 1.03 | [1.01; 1.05] |
| **Meta-Analysis** | | **1.03** | **[1.02; 1.05]** |

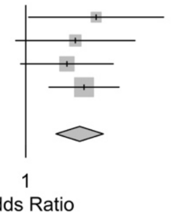

Heterogeneity: $I^2$ = 0.0%, $\tau^2$ = 0, $p$ = 0.8961

**S2.10**

### *MAP7D3* - eXCI

#### Index SNP: rs5975709

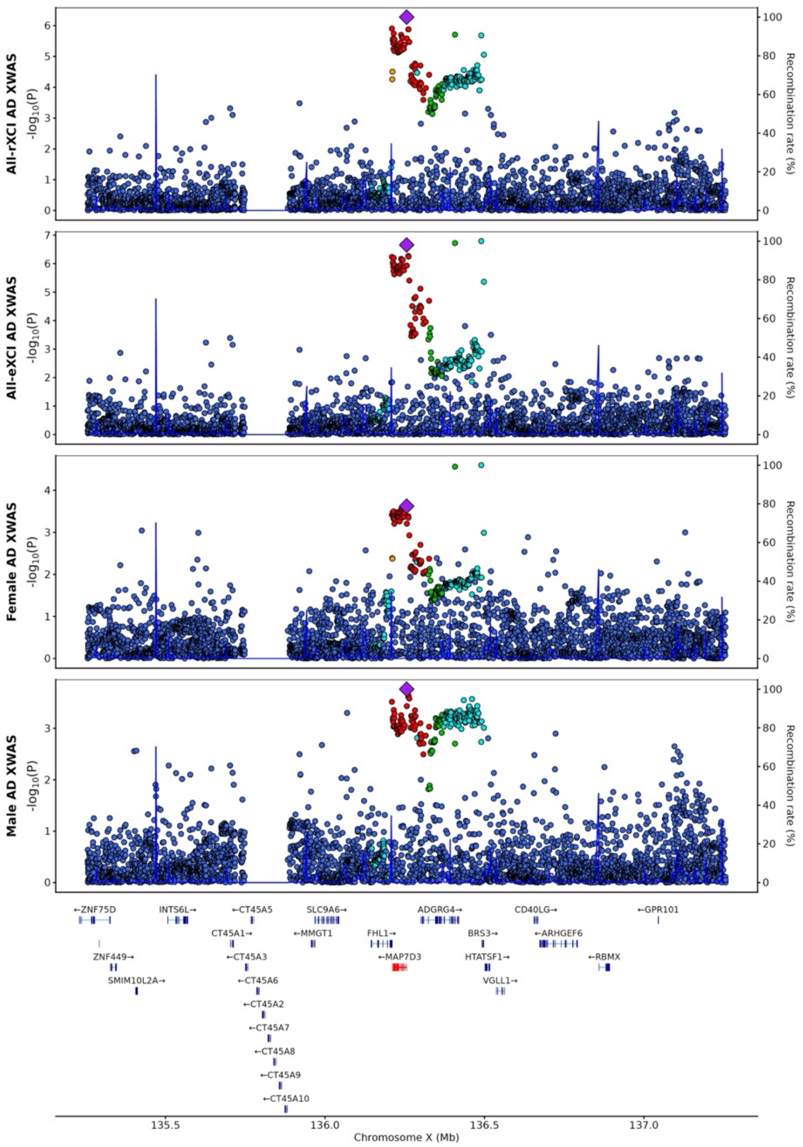

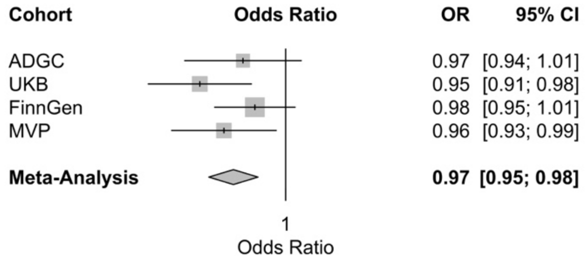

Heterogeneity: $I^2$ = 0.0%, $\tau^2$ < 0.0001, $p$ = 0.3991

**S2.11** ***CD99L2*** **- rXCI**

**Index SNP: rs146964414**

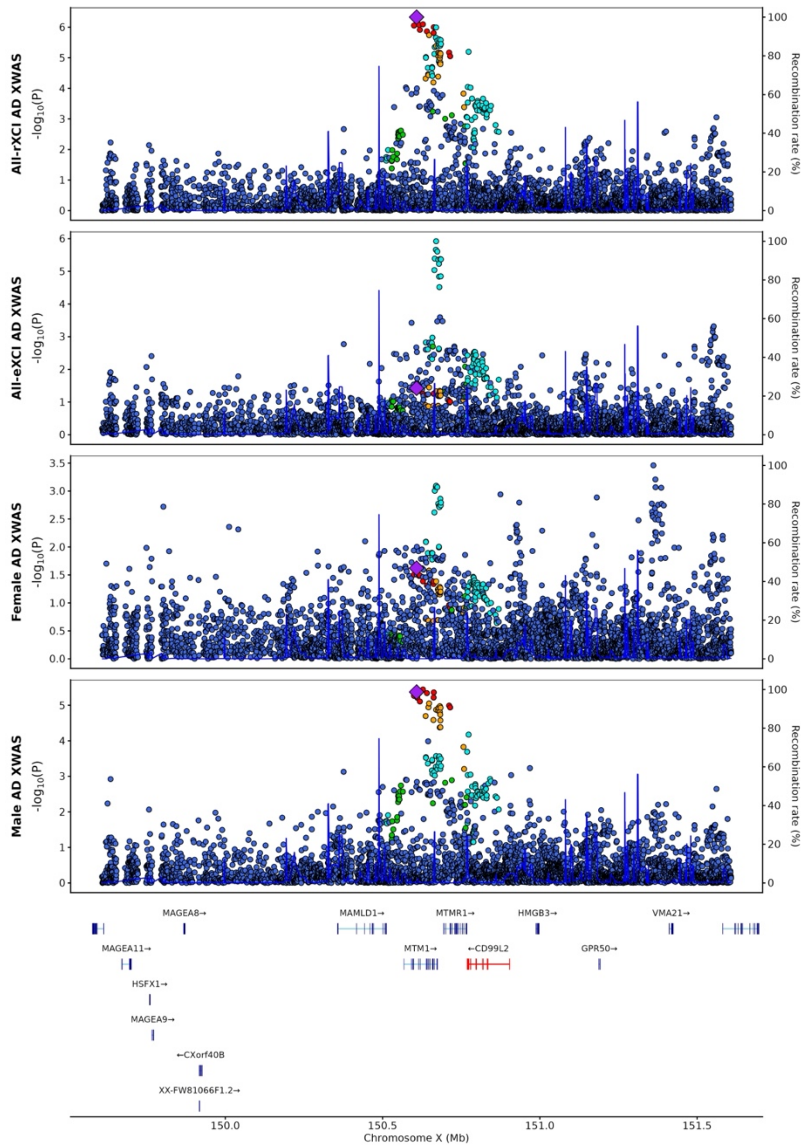

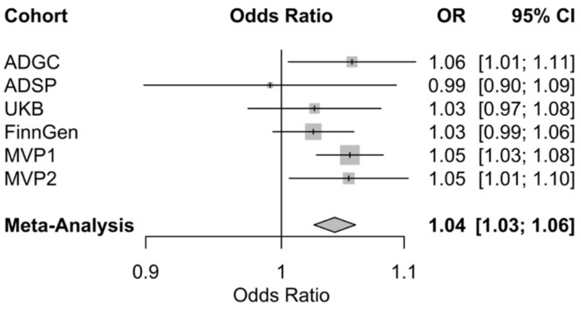

Heterogeneity: $I^2$ = 0.0%, $\tau^2$ = 0, $p$ = 0.5895

#### S2.12 *TGIF2LX* - eXCI

##### Index SNP: rs2752037

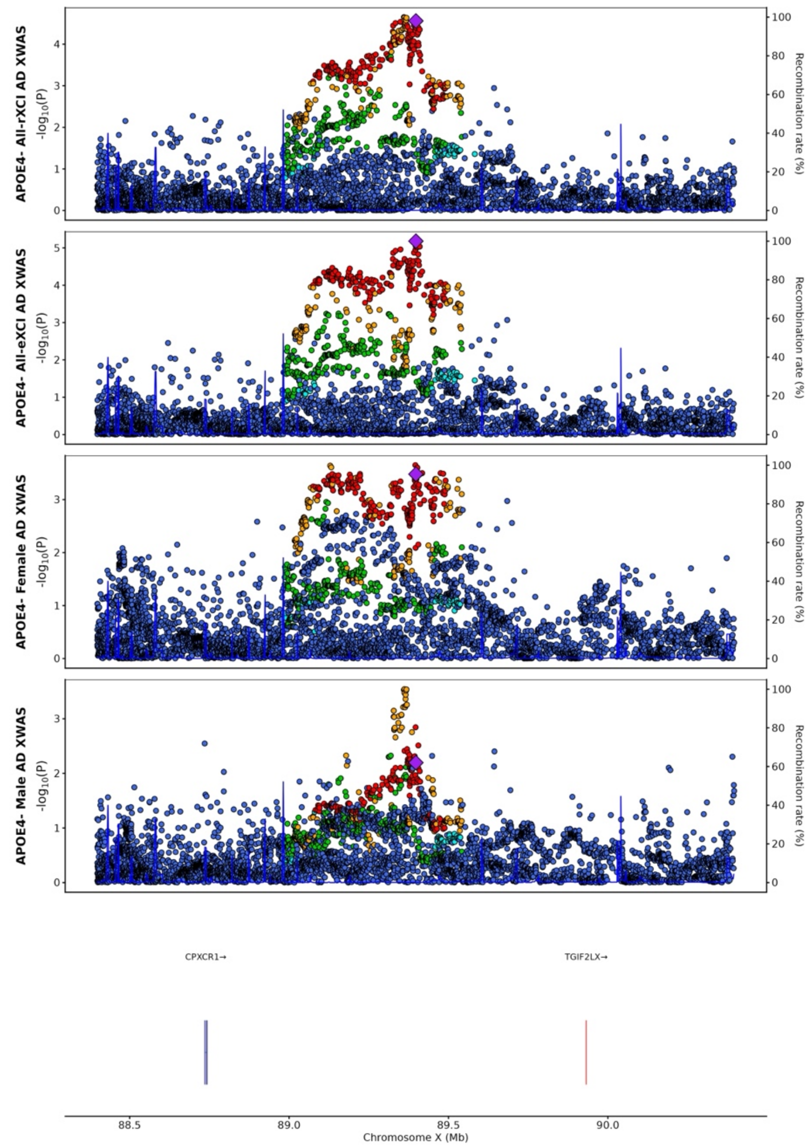

| Cohort | OR | 95% CI |
|---|---|---|
| ADGC | 0.95 | [0.89; 1.00] |
| ADSP | 0.93 | [0.84; 1.04] |
| UKB | 0.95 | [0.91; 0.99] |
| MVP | 0.96 | [0.94; 0.99] |
| **Meta-Analysis** | **0.96** | **[0.94; 0.97]** |

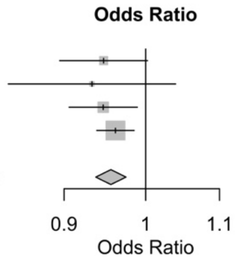

Heterogeneity: $I^2 = 0.0\%$, $\tau^2 = 0$, $p = 0.8673$

**S2.13**

#### *PLS3* - eXCI

##### Index SNP: rs146144791

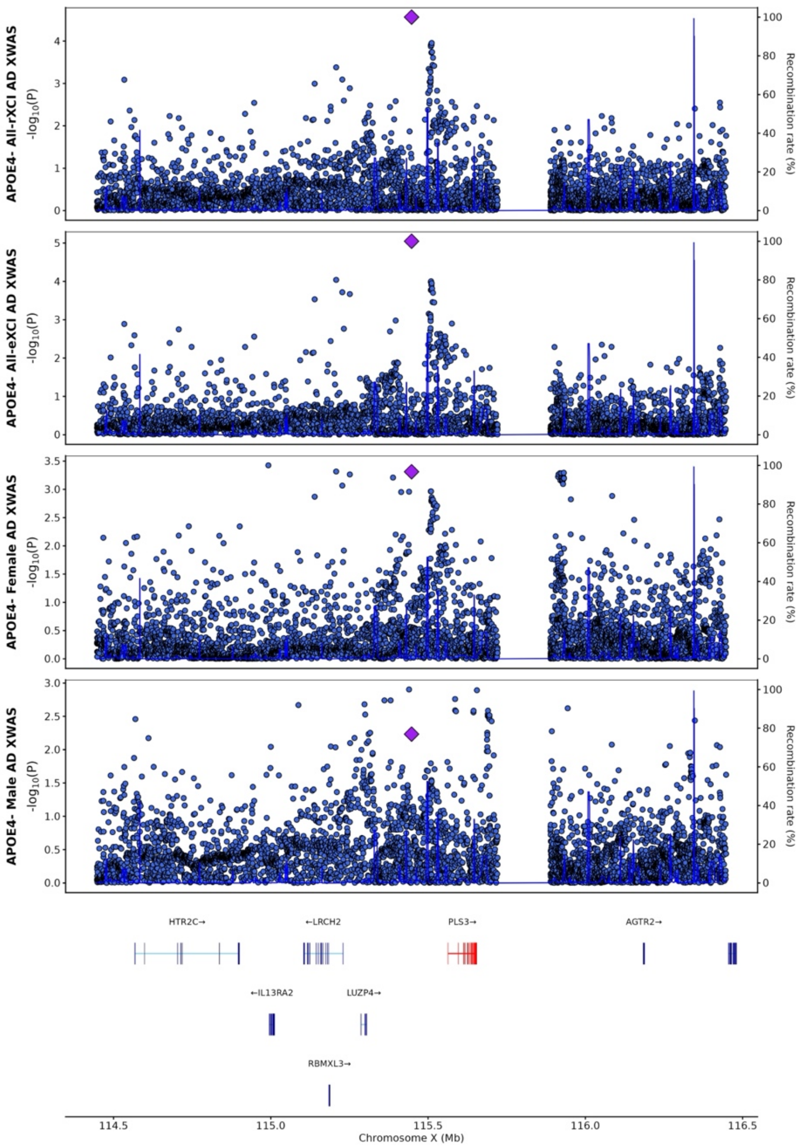

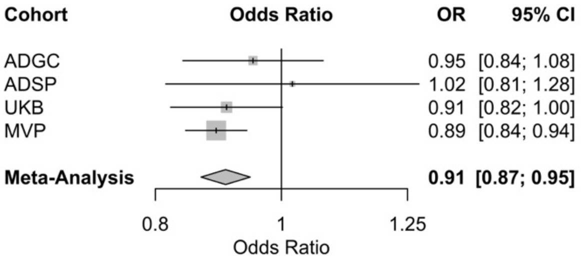

Heterogeneity: $I^2$ = 0.0%, $\tau^2$ = 0, $p$ = 0.5690

**S2.14**

#### *SMIM10L2B* - Female

##### Index SNP: rs12689435

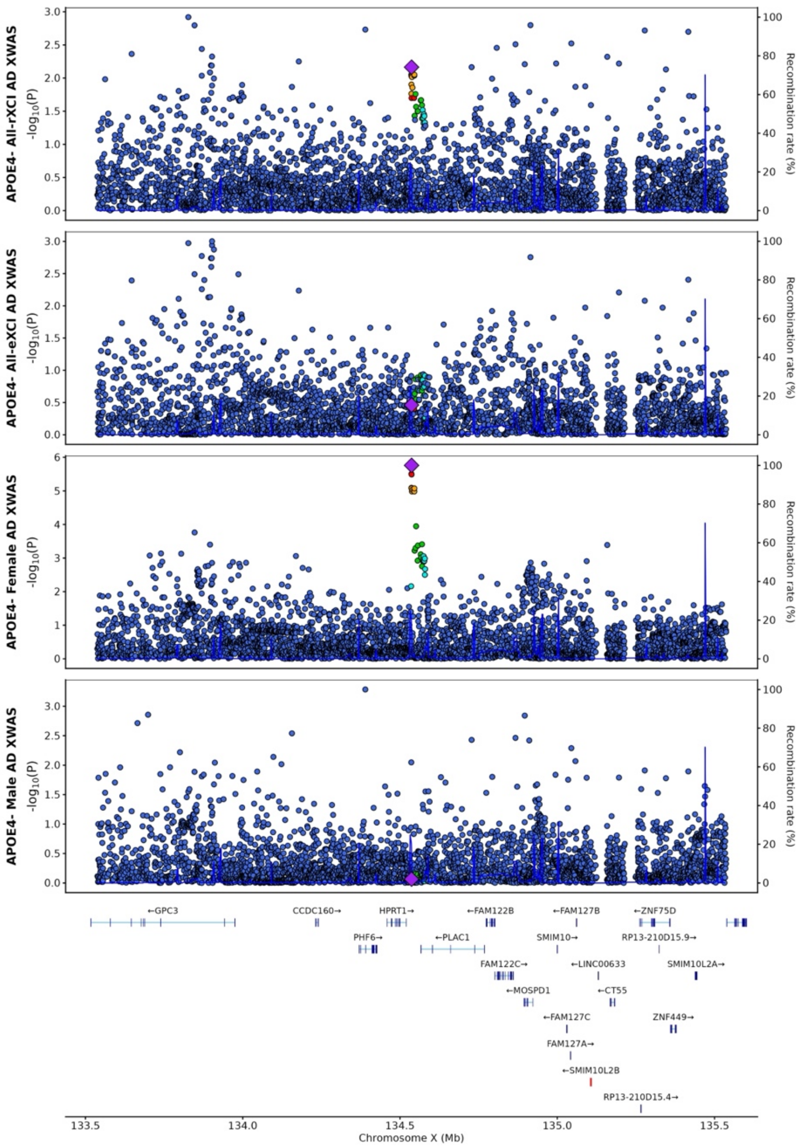

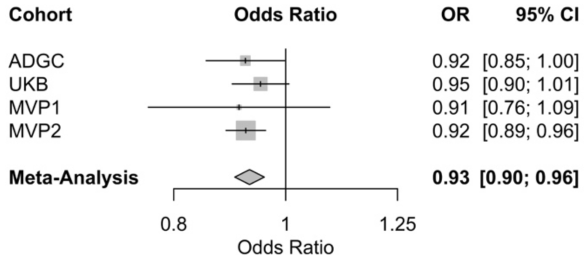

Heterogeneity: $I^2 = 0.0\%$, $\tau^2 = 0$, $p = 0.8468$

**S2.15** ***RBBP7* - eXCI**

**Index SNP: rs5924530**

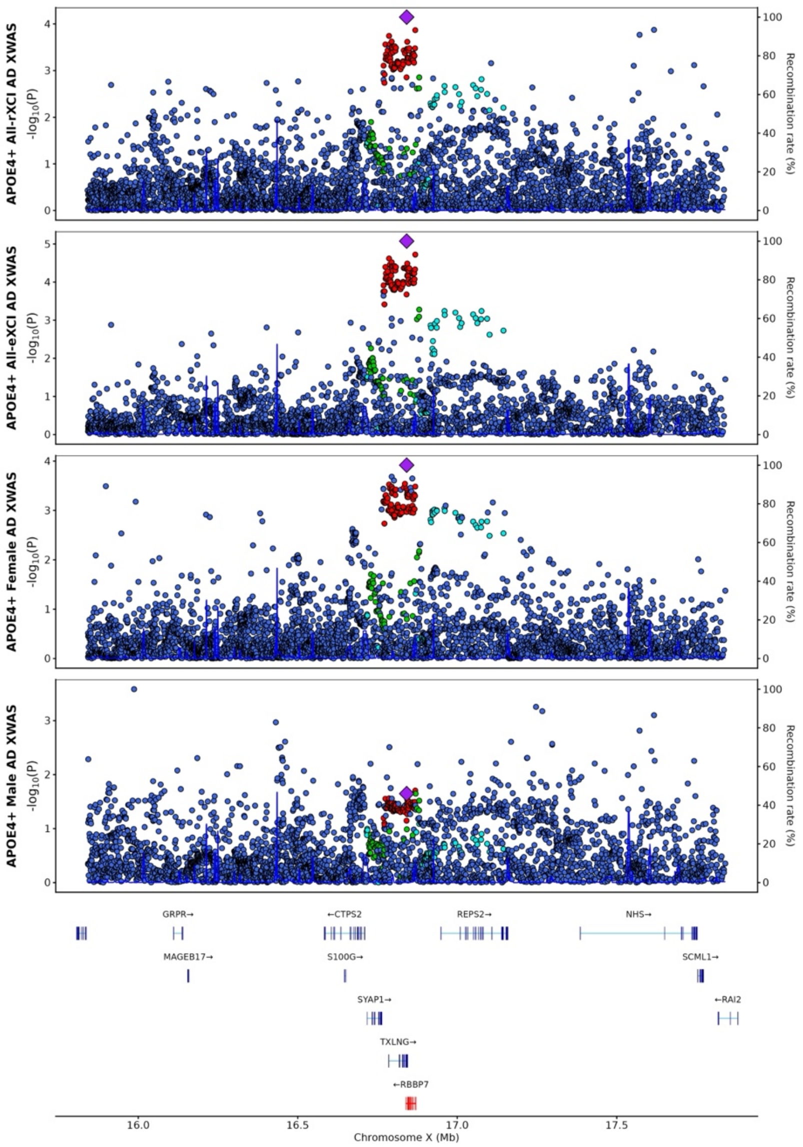

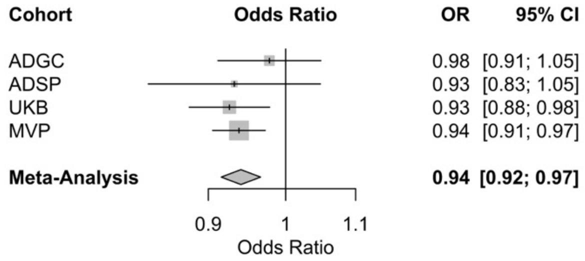

Heterogeneity: $I^2$ = 0.0%, $\tau^2$ = 0, $p$ = 0.6746

**S2.16**

#### *HTR2C* - eXCI

##### Index SNP: rs139542115

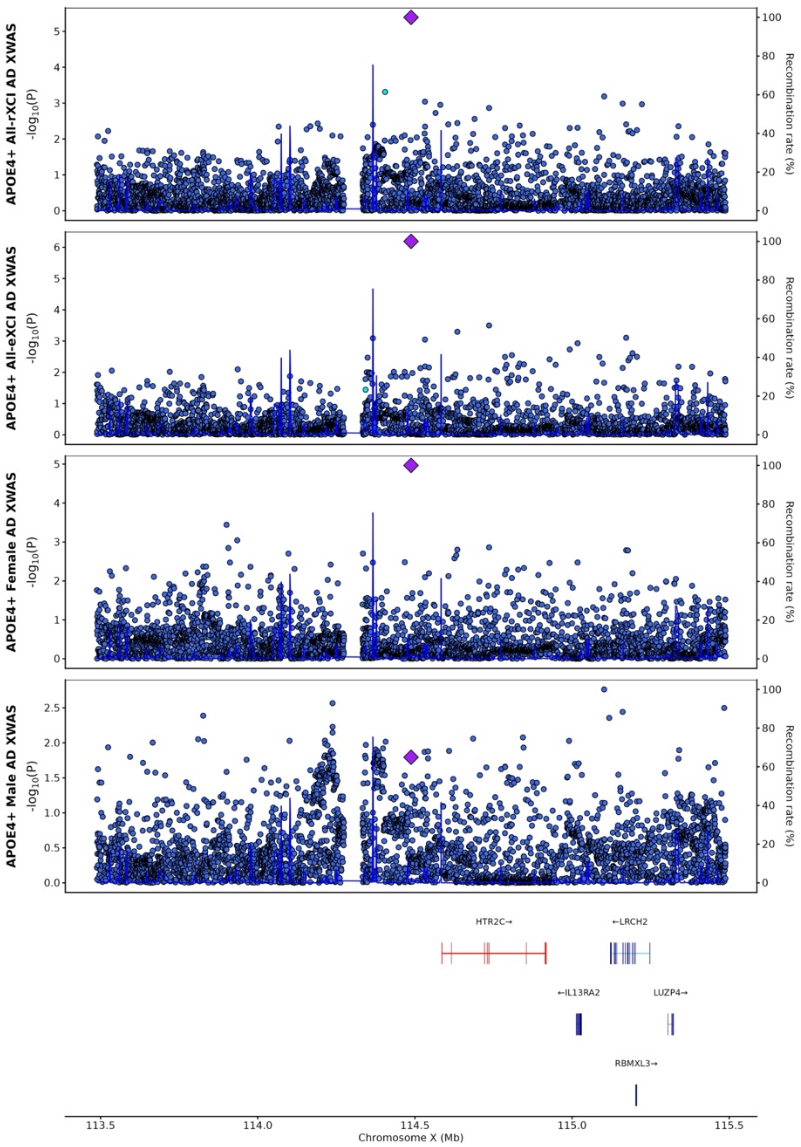

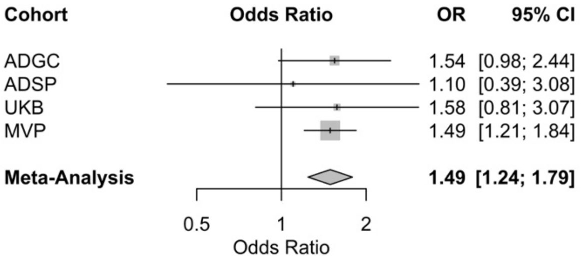

Heterogeneity: $I^2$ = 0.0%, $\tau^2$ = 0, $p$ = 0.9437

**S2.17**

#### *GPR101* - Male

##### Index SNP: rs146938321

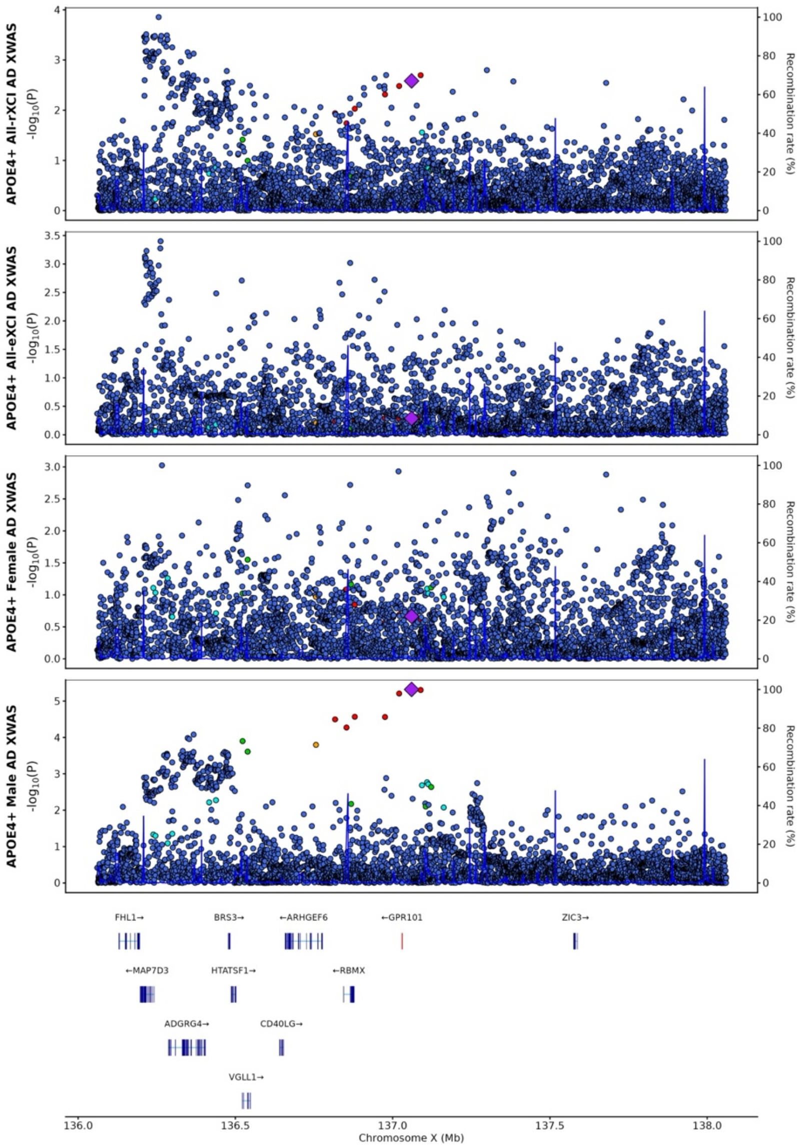

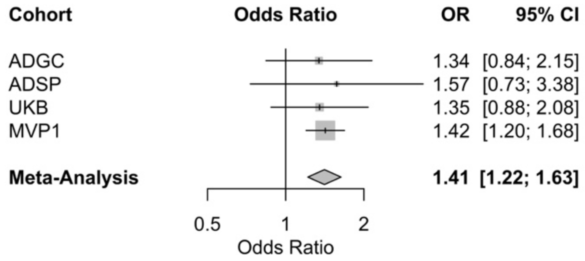

Heterogeneity: $I^2$ = 0.0%, $\tau^2$ = 0, $p$ = 0.9830

**S2.18**

### *SLITRK2* - rXCI

#### Index SNP: rs554300003

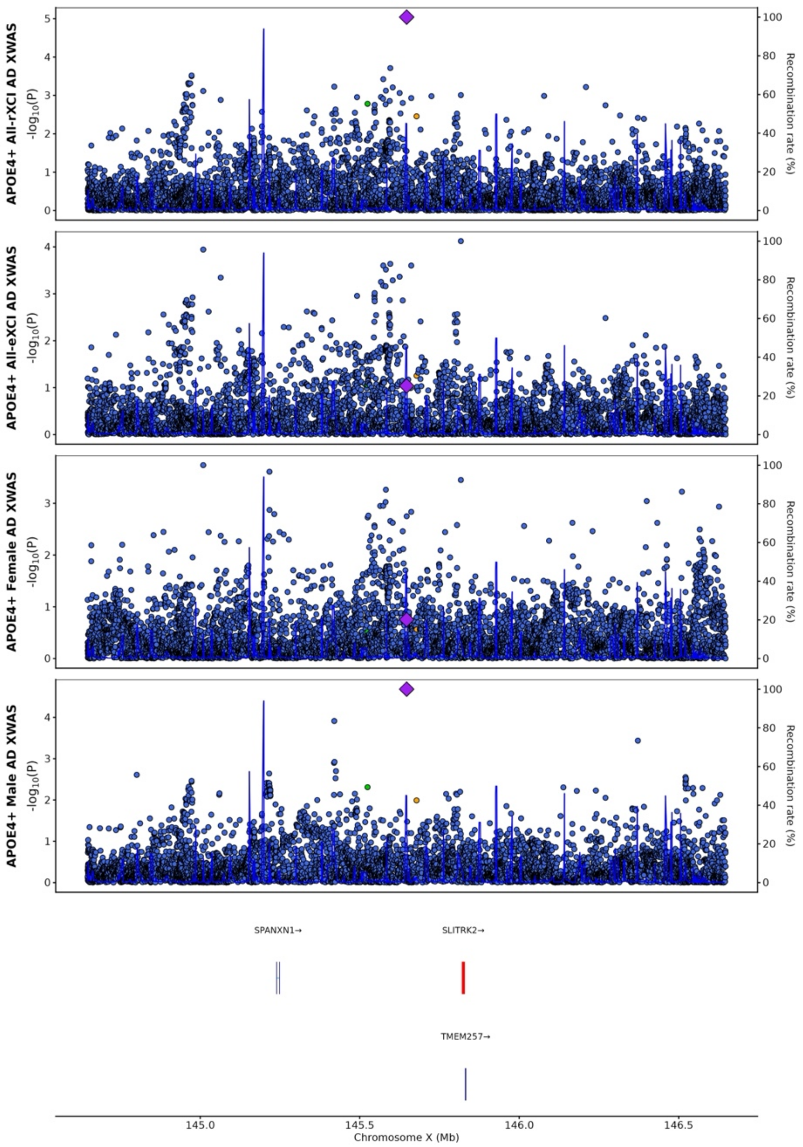

| Cohort | OR | 95% CI |
|---|---|---|
| ADGC | 1.55 | [1.04; 2.32] |
| UKB | 1.25 | [0.85; 1.83] |
| MVP1 | 1.38 | [1.17; 1.62] |
| MVP2 | 1.15 | [0.83; 1.60] |
| **Meta-Analysis** | **1.34** | **[1.18; 1.53]** |

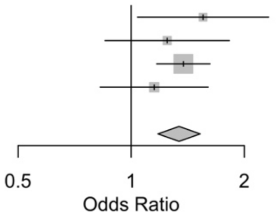

Heterogeneity: $I^2$ = 0.0%, $\tau^2$ = 0, $p$ = 0.6663

**Supplementary Figure 2. Locus zoom plots and forest plots for X-wide significant AD index variants. (1-11)** Non-*APOE**4-stratified, **(12-14)** *APOE**4 negative, **(15-18)** *APOE**4 positive. For each signal, the upper panels display stacked locus zoom plots (top to bottom): random X-chromosome inactivation model (rXCI), escape from X-chromosome inactivation model (eXCI), female-only, and male-only. Points represent $-\log_{10}(P)$ values for tested variants across the locus. The forest plot at the bottom of each panel displays per-cohort effect estimates (odds ratios and 95% confidence intervals) from the best XWAS model or sex along with the fixed-effects meta-analysis estimates and heterogeneity statistics. Dot color coding represents LD with tagging variant (purple diamond): red - R2: 0.8-1.0, orange - R2:0.6-0.8, green - R2:0.4-0.6, cyan - R2:0.2-0.4, blue - R2:0.0-0.2.

*Note that for the MAP7D3 locus (S2.10), the reported variant corresponds to the third most significant XWAS signal at the locus, as it demonstrated colocalization with external traits and xQTL signals. The two more significant variants at this locus (LD $R^2 \approx 0.4$) did not show evidence of colocalization with external traits or molecular phenotypes.*

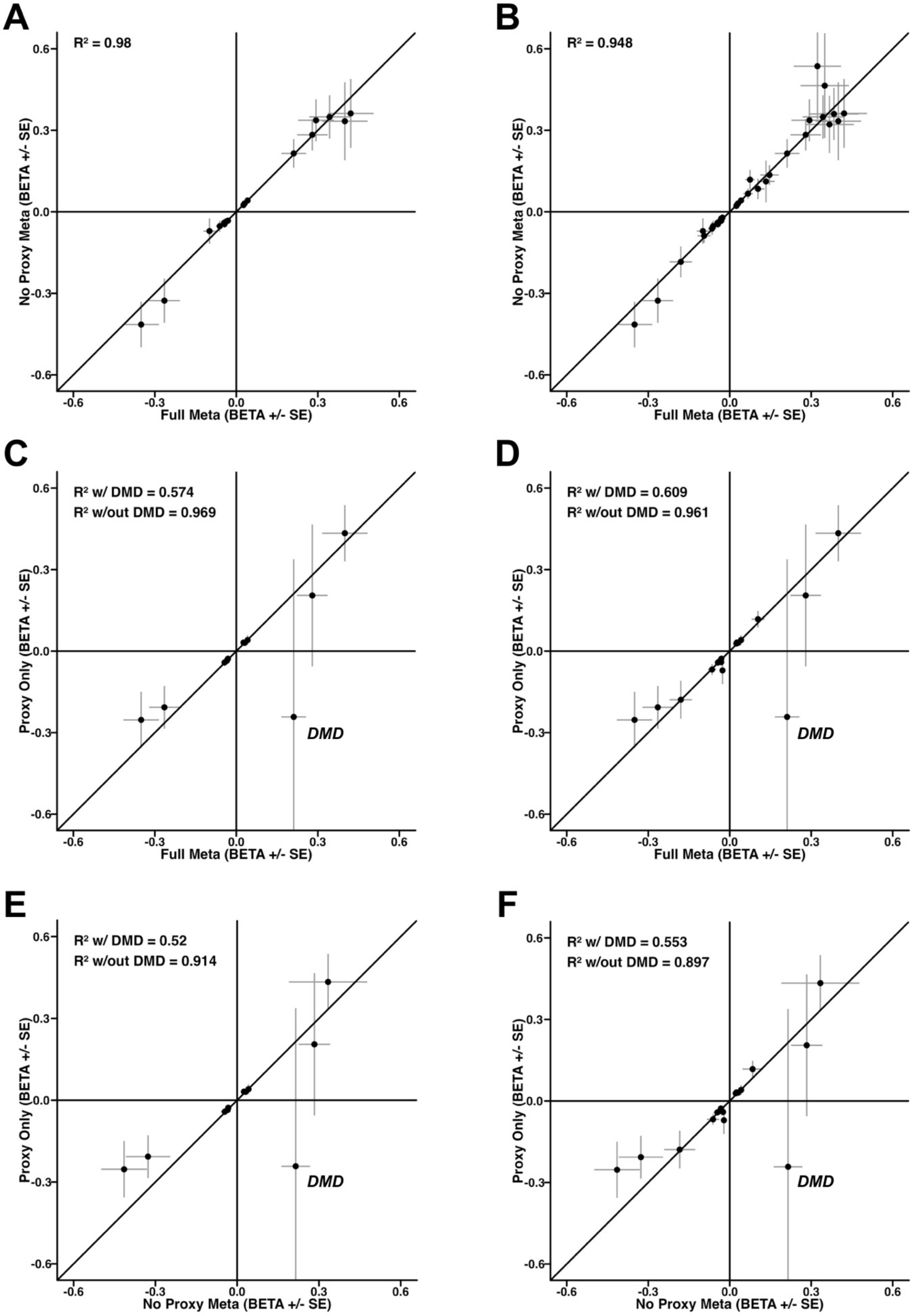
A
R² = 0.98
No Proxy Meta (BETA +/- SE)
Full Meta (BETA +/- SE)
B
R² = 0.948
No Proxy Meta (BETA +/- SE)
Full Meta (BETA +/- SE)
C
R² w/ DMD = 0.574
R² w/out DMD = 0.969
Proxy Only (BETA +/- SE)
Full Meta (BETA +/- SE)
DMD
D
R² w/ DMD = 0.609
R² w/out DMD = 0.961
Proxy Only (BETA +/- SE)
Full Meta (BETA +/- SE)
DMD
E
R² w/ DMD = 0.52
R² w/out DMD = 0.914
Proxy Only (BETA +/- SE)
No Proxy Meta (BETA +/- SE)
DMD
F
R² w/ DMD = 0.553
R² w/out DMD = 0.897
Proxy Only (BETA +/- SE)
No Proxy Meta (BETA +/- SE)
DMD

**Supplementary Figure 3. Concordance of effect sizes between full, no-proxy, and proxy-only XWAS meta-analyses.** Given potential concerns with the use of Alzheimer's disease proxy (family history) or health registry phenotypes, we compared effect size estimates (BETA ± SE) for AD X-wide significant and pleiotropy lead variants across meta-analysis designs. The diagonal line indicates y=x to provide a visual reference of equal effects. $R^2$ values denote the squared Pearson correlation of β estimates. **A)** Full meta-analysis (x-axis) versus no-proxy meta-analysis (y-axis) for the 18 independent XWAS index SNPs. **B)** Same comparison as in (A), additionally including prioritized variants from pleiotropy analyses that were not X-wide significant in the AD XWAS. **C)** Full meta-analysis (x-axis) versus proxy-only meta-analysis (y-axis) for the 11 index SNPs identified in the non-*APOE**4-stratified XWAS. **D)** Same comparison as in (C), additionally including prioritized non-*APOE**4-stratified pleiotropic variants not detected at X-wide significance. **E)** No-proxy meta-analysis (x-axis) versus proxy-only meta-analysis (y-axis) for the 11 non-*APOE**4-stratified index SNPs. **F)** Same comparison as in (E), additionally including prioritized non-*APOE**4-stratified pleiotropic variants not detected at X-wide significance.

*Note panels C-F are restricted to non-APOE*4-stratified signals due to limited power to estimate proxy-only effects for APOE*4-stratified loci. The variant at the DMD locus shows an outlier effect in proxy-only analyses; therefore,* $R^2$ *is reported with and without this variant where indicated.*

#### Gene Prioritization

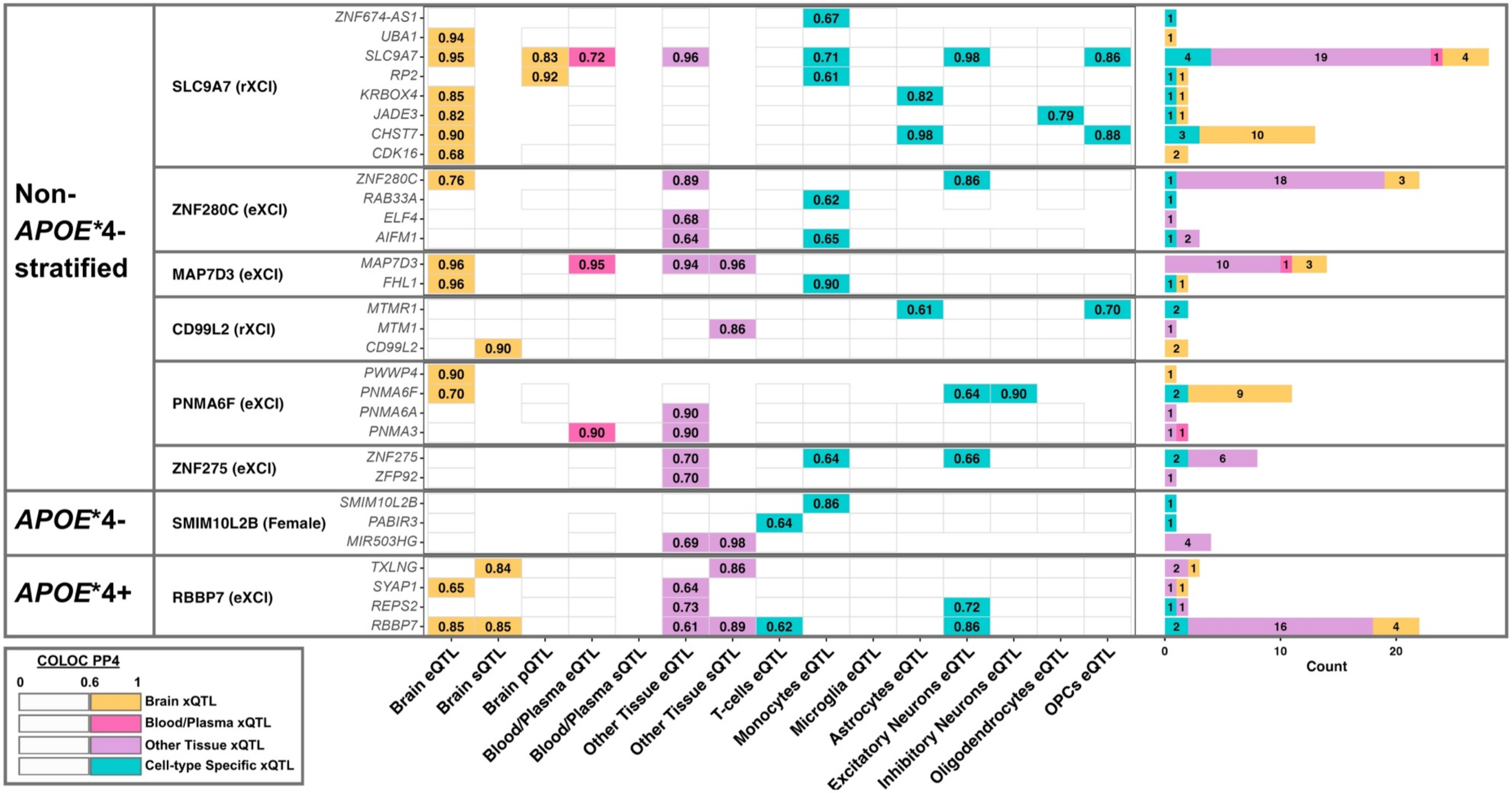

**Supplementary Figure 4. Summary of xQTL colocalization candidate genes at X-linked AD loci.** Shown are the nine loci for which at least one gene demonstrated evidence of colocalization (PP4 > 0.6) in ≥1 dataset. The heatmap displays PP4 values exceeding 0.6 across QTL modality and tissue/cell-type categories. Only PP4 > 0.6 are shown; blank cells indicate PP4 ≤ 0.6. Columns are organized by QTL type, and for aggregated categories (e.g., "other tissue eQTL"), the displayed value represents the maximum PP4 observed across all datasets within that category (for example, across 35 GTEx v8 tissues). The bar plot on the right summarizes the total number of unique xQTL colocalizations (PP4 > 0.6) per gene across all evaluated datasets. Because some columns represent aggregated categories, the number of colocalizations summarized in the bar plot may exceed the number of visible heatmap cells for a given gene.

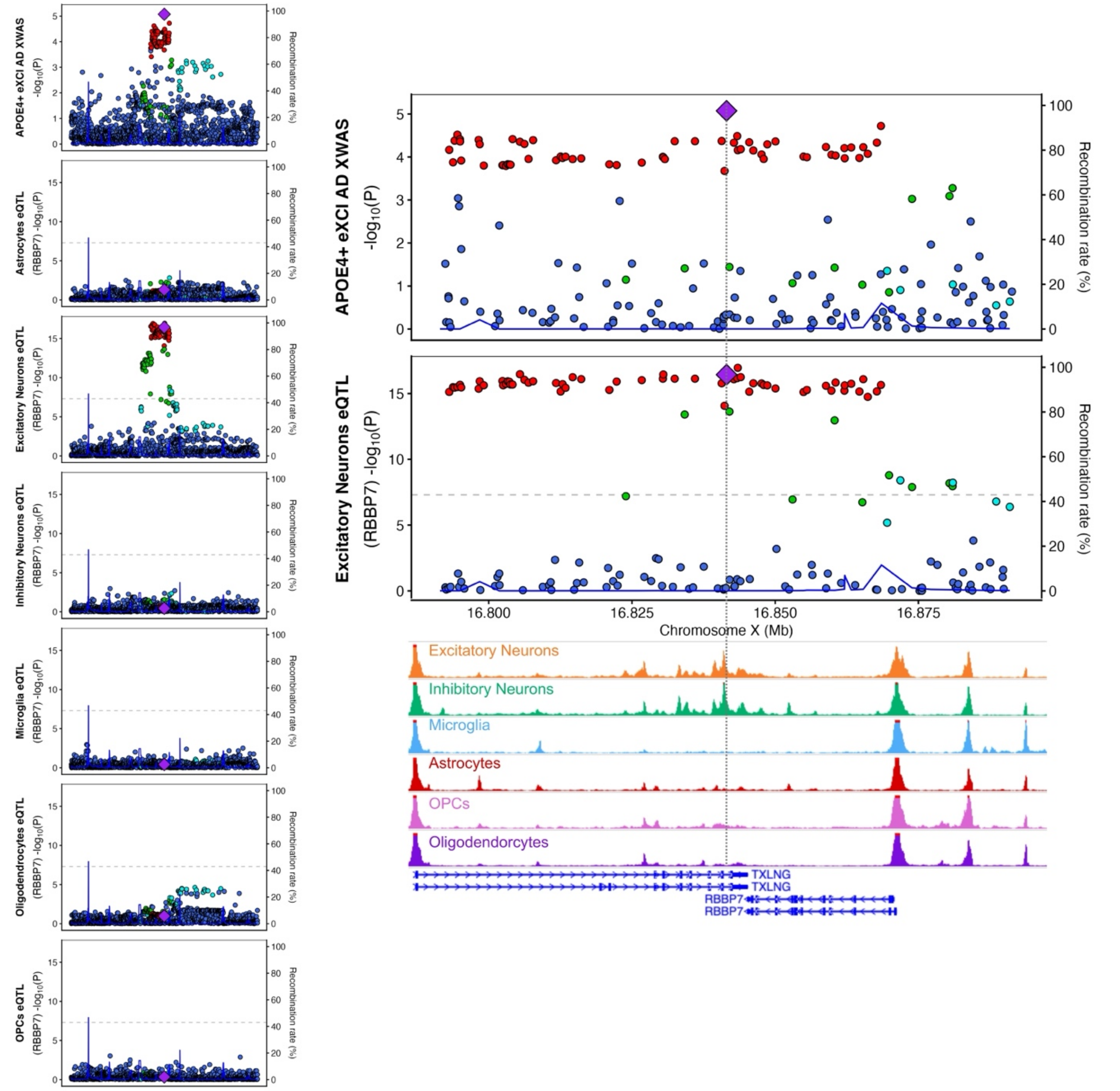

**Supplementary Figure 5. Cell-type-specific regulatory context of the *APOE**4 positive *RBBP7* AD signal.** Left panels show regional association plots for *APOE**4 positive eXCI AD XWAS and eQTLs for *RBBP7* across brain cell types (astrocytes, excitatory neurons, inhibitory neurons, microglia, oligodendrocytes, and OPCs). The AD association signal shows strong colocalization with the excitatory neuron eQTL for *RBBP7*, whereas other cell-types did not display meaningful eQTL signals. Right panels display zoomed-in locus plots for the *RBBP7* AD locus association signal, excitatory neuron *RBBP7* eQTL data, and their overlap with chromatin accessibility (ATAC-seq) tracks in 6 brain cell-types. This shows the AD index variant overlaps an open chromatin peak in excitatory neurons (confirming through direct peak coordinate data) and inhibitory neurons but not in other brain cell-types. Together, these data support a model in which the *APOE**4 positive AD association at this locus operates through excitatory neuron-specific regulatory effects on *RBBP7* expression. Dot color coding represents LD with tagging variant (purple diamond): red - R2: 0.8-1.0, orange - R2:0.6-0.8, green - R2:0.4-0.6, cyan - R2:0.2-0.4, blue - R2:0.0-0.2.

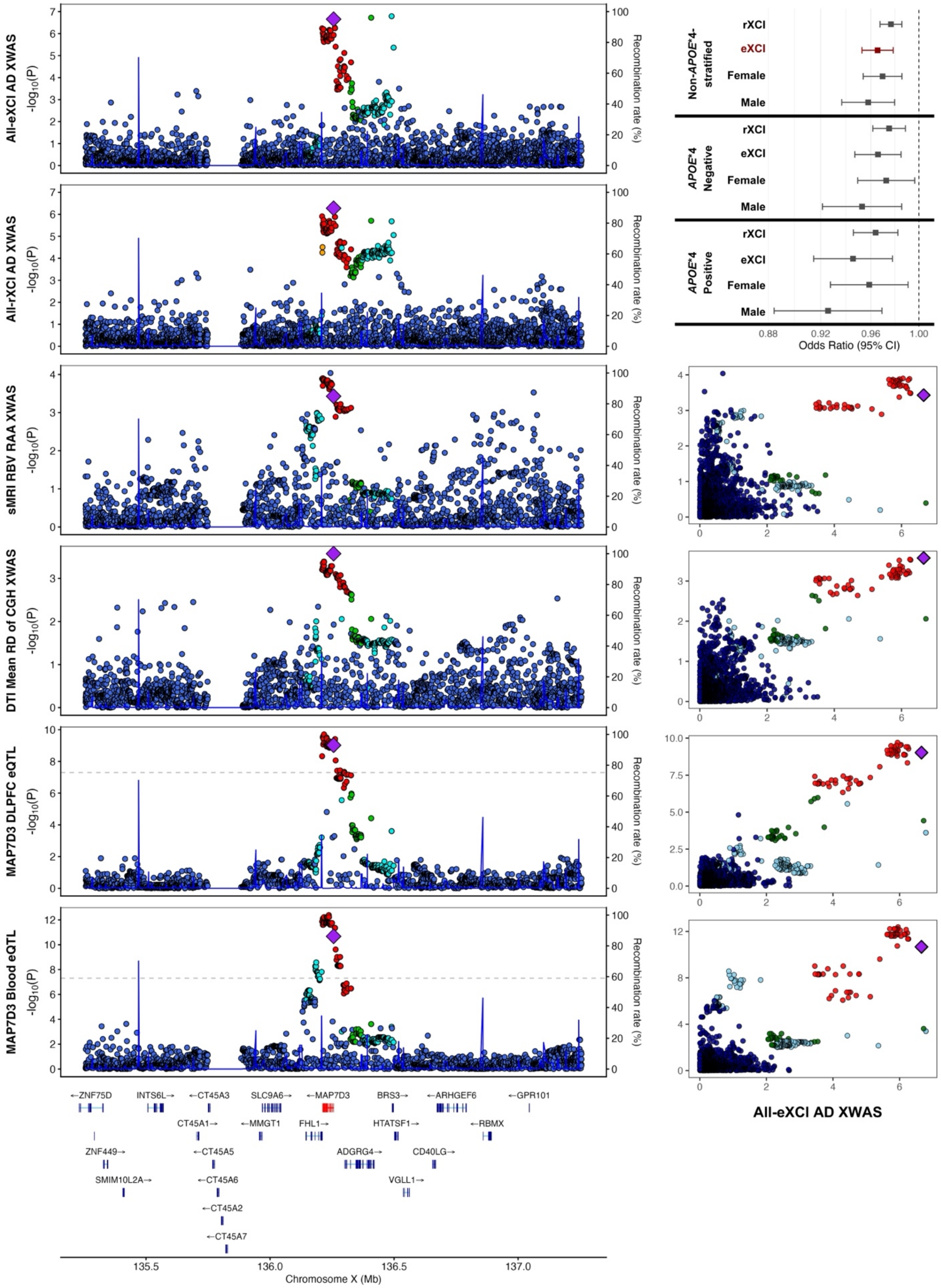

All-eXCI AD XWAS
All-rXCI AD XWAS
sMRI RBV RAA XWAS
DTI Mean RD of CGH XWAS
MAP7D3 DLPFC eQTL
MAP7D3 Blood eQTL
-log10(P)
Recombination rate (%)
Chromosome X (Mb)
Non-APOE*4-stratified
APOE*4 Negative
APOE*4 Positive
rXCI
eXCI
Female
Male
Odds Ratio (95% CI)
All-eXCI AD XWAS

**Supplementary Figure 6. Summary of cross-trait findings at the *MAP7D3* locus.** The stacked locus zoom panels (top to bottom) display: Non-*APOE**4–stratified eXCI AD XWAS, Non-*APOE**4–stratified rXCI AD XWAS, structural-MRI regional brain volume in the right accumbens area, Diffusion tensor imaging mean radial diffusivity in the Cingulum (hippocampus), Dorsolateral prefrontal cortex eQTL for *MAP7D3*, Blood eQTL for *MAP7D3*. The upper-right forest plot summarizes effect size estimates (odds ratios and 95% confidence intervals) for the tagged variant across all 12 model, sex, and *APOE**4 stratum combinations, with the prioritized model and stratum highlighted in red. The lower-right panels show locus-compare scatter plots corroborating evidence of colocalization between the AD XWAS association statistics and the corresponding trait in each row of the stacked regional plots (y-axis). Dot color coding represents LD with tagging variant (purple diamond): red - R2: 0.8-1.0, orange - R2:0.6-0.8, green - R2:0.4-0.6, cyan - R2:0.2-0.4, blue - R2:0.0-0.2.

*Abbreviations: CGH, cingulum (hippocampus); CI, confidence interval; DLPFC, dorsolateral prefrontal cortex; DTI, diffusion tensor imaging; RAA, right accumbens area; RBV, regional brain volume; RD, radial diffusivity; sMRI, structural magnetic resonance imaging*

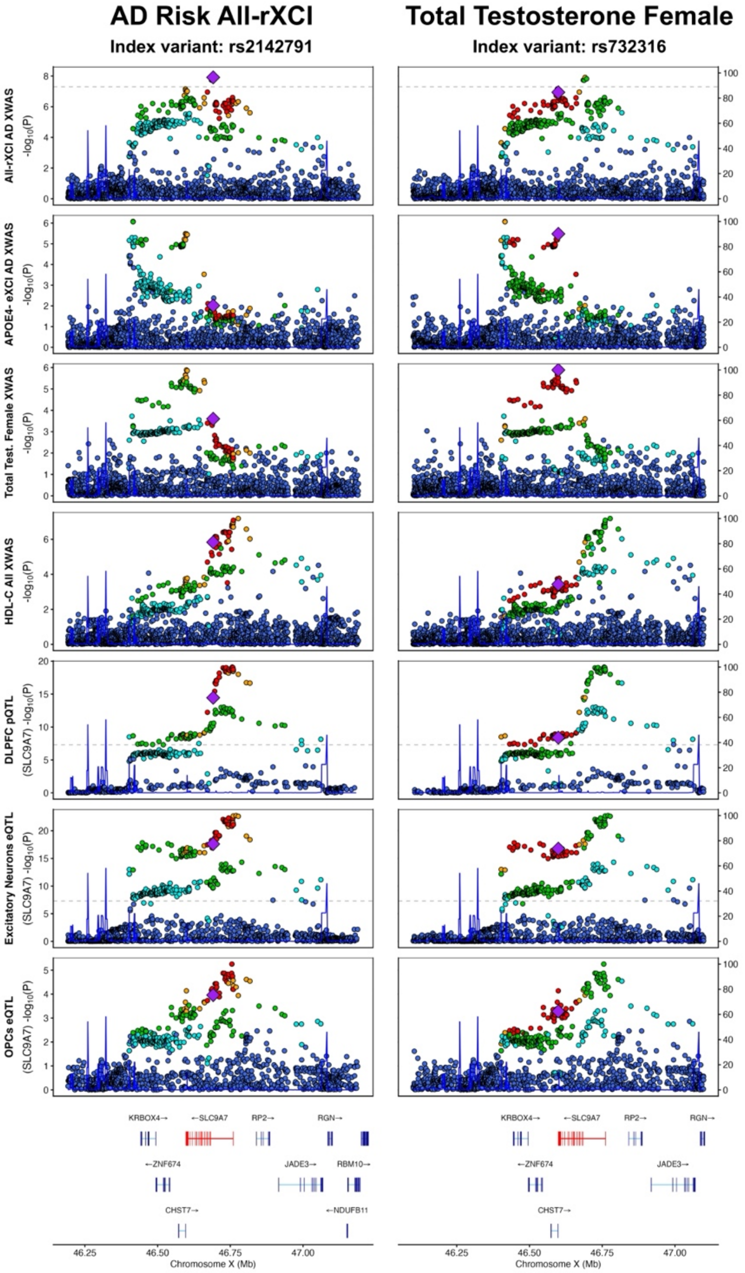

**Supplementary Figure 7. Summary of cross-trait findings at the *SLC9A7* locus.** Regional association plots at the *SLC9A7* locus highlighting two index variants that suggest there appear to be at least 2 independent signals in partial LD (rs2142791-rXCI versus rs732316-eXCI). Each column is centered on a different index variant: Left, AD XWAS lead variant (non-*APOE**4-stratified rXCI model); Right, female total testosterone lead variant. For each tagged variant, seven traits are shown (top to bottom): Non-*APOE**4-stratified AD XWAS, *APOE**4 negative eXCI AD XWAS, Female total testosterone XWAS, HDL-cholesterol XWAS, DLPFC pQTL for SLC9A7, Excitatory neuron eQTL for *SLC9A7*, Oligodendrocyte precursor cell eQTL for *SLC9A7*. Dot color coding represents LD with tagging variant (purple diamond): red - R2: 0.8-1.0, orange - R2:0.6-0.8, green - R2:0.4-0.6, cyan - R2:0.2-0.4, blue - R2:0.0-0.2.

*Abbreviations: DLPFC, dorsolateral prefrontal cortex; OPC, oligodendrocyte precursor cells, Test., testosterone.*

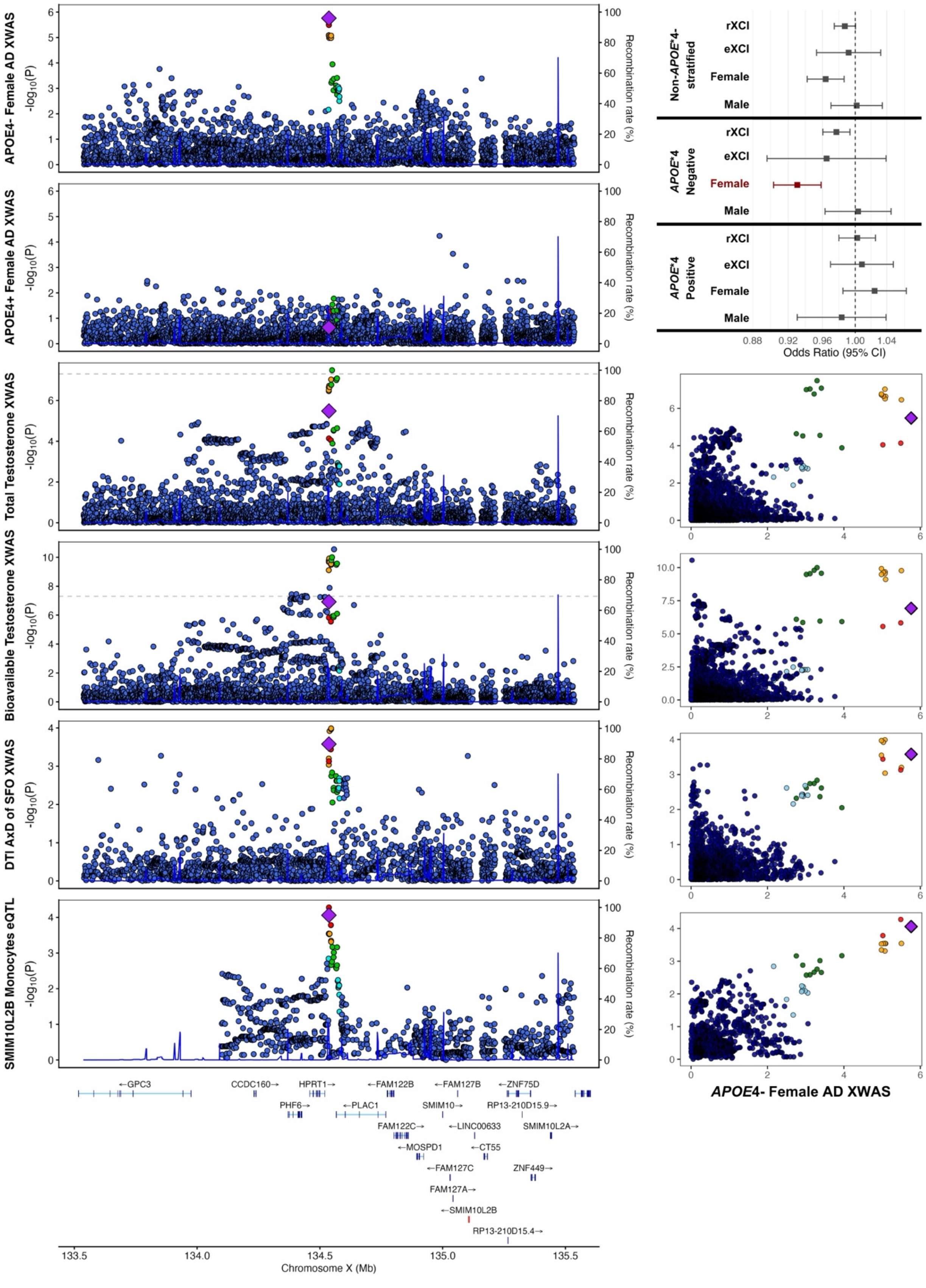

APOE4- Female AD XWAS
APOE4+ Female AD XWAS
Total Testosterone XWAS
Bioavailable Testosterone XWAS
DTI AxD of SFO XWAS
SMIM10L2B Monocytes eQTL
-log10(P)
Recombination rate (%)
Chromosome X (Mb)
GPC3
CCDC160
HPRT1
FAM122B
FAM127B
ZNF75D
PHF6
PLAC1
SMIM10
RP13-210D15.9
FAM122C
LINC00633
SMIM10L2A
MOSPD1
CT55
FAM127C
ZNF449
FAM127A
SMIM10L2B
RP13-210D15.4
Non-APOE*4-stratified
APOE*4 Negative
APOE*4 Positive
rXCI
eXCI
Female
Male
Odds Ratio (95% CI)
APOE4- Female AD XWAS

**Supplementary Figure 8. Summary of cross-trait findings at the *SMIM10L2B* locus.** The stacked locus zoom panels (top to bottom) display: *APOE**4 negative female AD XWAS, *APOE**4 positive female AD XWAS, Total testosterone, Bioavailable testosterone, Diffusion tensor imaging axial diffusivity in the Superior fronto-occipital fasciculus, Monocyte eQTL for *SMIM10L2B*. The upper-right forest plot summarizes effect size estimates (odds ratios and 95% confidence intervals) for the tagged variant across all 12 model, sex, and *APOE**4 stratum combinations, with the prioritized model and stratum highlighted in red. The lower-right panels show locus-compare scatter plots corroborating evidence of colocalization between the AD XWAS association statistics and the corresponding trait in each row of the stacked regional plots (y-axis). Dot color coding represents LD with tagging variant (purple diamond): red - R2: 0.8-1.0, orange - R2:0.6-0.8, green - R2:0.4-0.6, cyan - R2:0.2-0.4, blue - R2:0.0-0.2.

*Abbreviations: AxD, axial diffusivity; CI, confidence interval; DTI, diffusion tensor imaging; RAA, right accumbens area; SFO, superior fronto-occipital fasciculus*

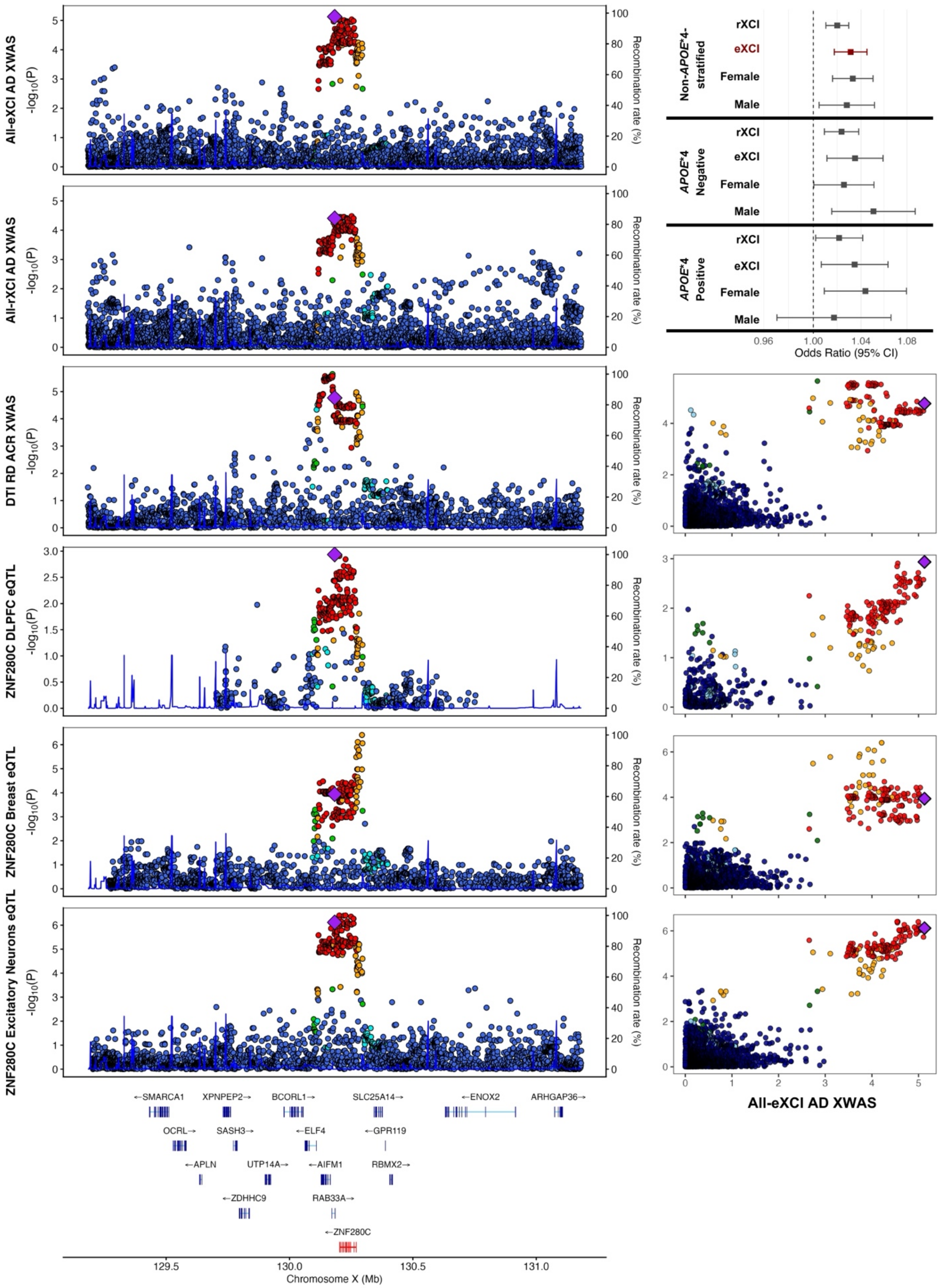

All-eXCI AD XWAS
All-rXCI AD XWAS
DTI RD ACR XWAS
ZNF280C DLPFC eQTL
ZNF280C Breast eQTL
ZNF280C Excitatory Neurons eQTL
-log10(P)
Recombination rate (%)
←SMARCA1
XPNPEP2→
BCORL1→
SLC25A14→
←ENOX2
ARHGAP36→
OCRL→
SASH3→
←ELF4
←GPR119
←APLN
UTP14A→
←AIFM1
RBMX2→
←ZDHHC9
RAB33A→
←ZNF280C
129.5
130.0
130.5
131.0
Chromosome X (Mb)
Non-APOE*4-stratified
APOE*4 Negative
APOE*4 Positive
rXCI
eXCI
Female
Male
0.96
1.00
1.04
1.08
Odds Ratio (95% CI)
All-eXCI AD XWAS

**Supplementary Figure 9. Summary of cross-trait findings at the *ZNF280C* locus.** The stacked locus zoom panels (top to bottom) display: Non-*APOE**4–stratified eXCI AD XWAS, Non-*APOE**4–stratified rXCI AD XWAS, Diffusion tensor imaging radial diffusivity in the anterior corona radiata, Dorsolateral prefrontal cortex eQTL for *ZNF280C*, Breast tissue eQTL for *ZNF280C*, Excitatory neuron eQTL for *ZNF280C*. The upper-right forest plot summarizes effect size estimates (odds ratios and 95% confidence intervals) for the tagged variant across all 12 model, sex, and *APOE**4 stratum combinations, with the prioritized model and stratum highlighted in red. The lower-right panels show locus-compare scatter plots corroborating evidence of colocalization between the AD XWAS association statistics and the corresponding trait in each row of the stacked regional plots (y-axis). Dot color coding represents LD with tagging variant (purple diamond): red - R2: 0.8-1.0, orange - R2:0.6-0.8, green - R2:0.4-0.6, cyan - R2:0.2-0.4, blue - R2:0.0-0.2.

*Abbreviations: ACR, anterior corona radiata; CI, confidence interval; DLPFC, dorsolateral prefrontal cortex; DTI, diffusion tensor imaging; RD, radial diffusivity*

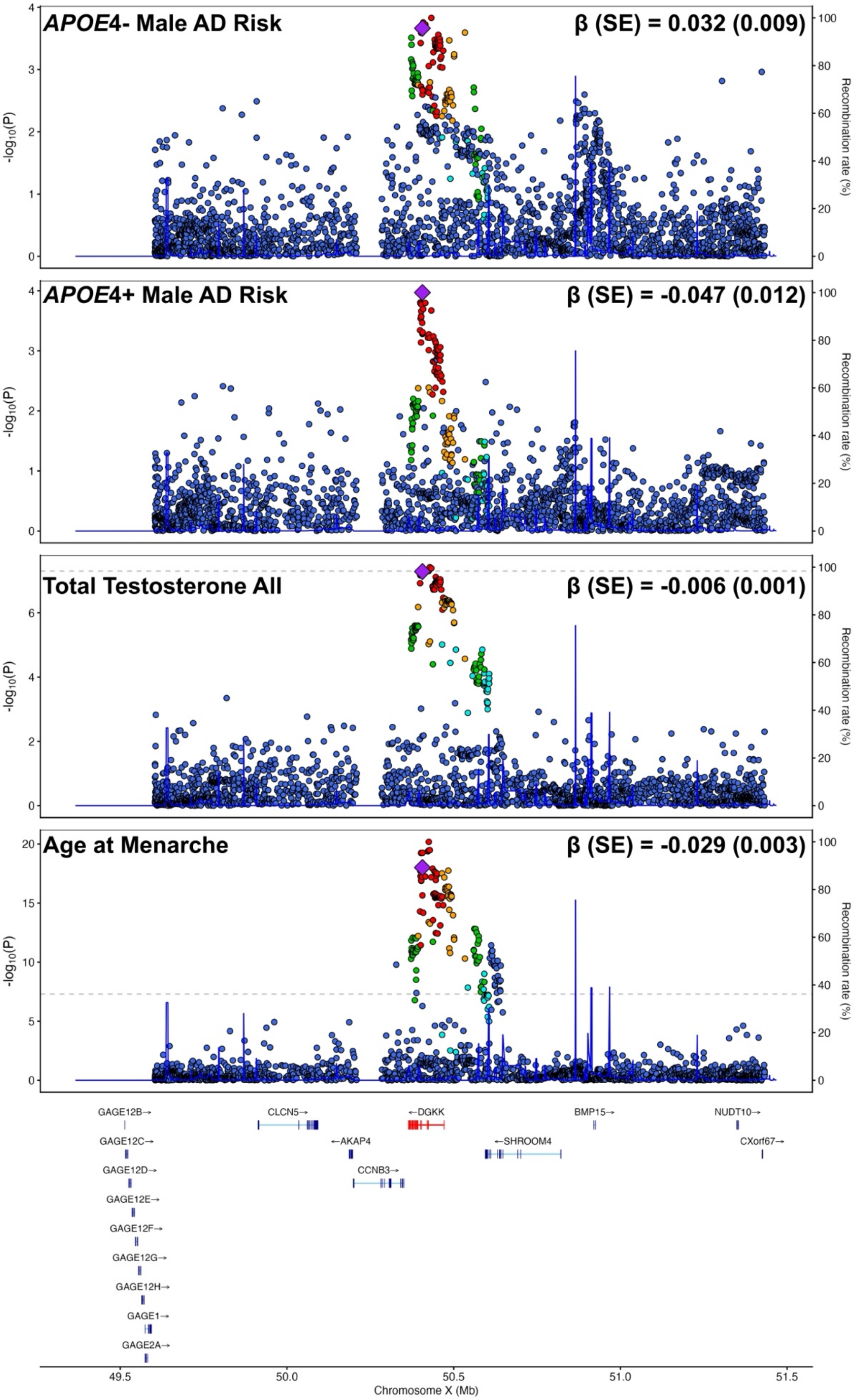

APOE4- Male AD Risk
β (SE) = 0.032 (0.009)
APOE4+ Male AD Risk
β (SE) = -0.047 (0.012)
Total Testosterone All
β (SE) = -0.006 (0.001)
Age at Menarche
β (SE) = -0.029 (0.003)
-log10(P)
Recombination rate (%)
GAGE12B→
GAGE12C→
GAGE12D→
GAGE12E→
GAGE12F→
GAGE12G→
GAGE12H→
GAGE1→
GAGE2A→
CLCN5→
←AKAP4
CCNB3→
←DGKK
←SHROOM4
BMP15→
NUDT10→
CXorf67→
49.5
50.0
50.5
51.0
51.5
Chromosome X (Mb)

**Supplementary Figure 10. Male-specific *DGKK* locus shows *APOE**4-stratified effect heterogeneity.** Regional association plots highlighting a male-specific AD risk signal at the *DGKK* locus. The lead variant demonstrates opposite directions of effect between *APOE**4 negative and *APOE**4 positive males, indicating *APOE**4-dependent modulation of the genetic association at this locus. Notably, the region shows overlapping association patterns with circulating testosterone levels and age at menarche. Dot color coding represents LD with tagging variant (purple diamond): red - R2: 0.8-1.0, orange - R2:0.6-0.8, green - R2:0.4-0.6, cyan - R2:0.2-0.4, blue - R2:0.0-0.2.
